# The Effect of Adolescent Pregnancy on Child Mortality in 46 Low- and Middle-Income Countries

**DOI:** 10.1101/2021.06.10.21258227

**Authors:** Navideh Noori, Joshua L. Proctor, Yvette Efevbera, Assaf P. Oron

## Abstract

**Introduction:** Adolescent pregnancy is a known health risk to mother and child. Statements and reports of health outcomes typically group mothers under 20 years old together. Few studies examined this risk at a finer age resolution, none of them comprehensively, and with differing results.

**Methods:** We analyzed Demographic and Health Surveys (DHS) data from 2004-2018 in Sub-Saharan Africa (SSA) and South Asia, on firstborn children of mothers 25 years old or younger. We examined the association between maternal age and stillbirths, and rates of neonatal (NNMR), infant (IMR), and under-5 mortality (U5MR), using mixed-effects logistic regression adjusting for major demographic variables and exploring the impact of maternal health-seeking.

**Results:** In both regions and across all endpoints, mortality rates of children born to mothers aged <16 years, 16-17 years, and 18-19 years at first birth were about 2-4 times, 1.5-2 times, and 1.2-1.5 times higher, respectively, than among firstborn children of mothers aged 23-25. Absolute mortality rates declined over time, but the age gradient remained similar across time periods and regions. Adjusting for rural/urban residence and maternal education, in SSA in 2014-2018 having a <16 year old mother was associated with odds ratio (ORs) of 3.71 [95% CI 2.50–5.51] for stillbirth, 1.92 [1.60–2.30] for NNMR, 2.13 [1.85–2.46] IMR, and 2.39 [2.13–2.68] U5MR, compared with having a mother aged 23-25. In South Asia in 2014-2018 ORs were 5.12 [2.85–9.20] stillbirth, 2.46 [2.03–2.97] NNMR, 2.62 [2.22–3.08] IMR, and 2.59 [2.22–3.03] U5MR. Part of the effect on NNMR and IMR may be mediated by a lower maternal health-seeking rate.

**Conclusions:** Adolescent pregnancy is associated with dramatically worse child survival and mitigated by health-seeking behavior, likely reflecting a combination of biological and social factors. Refining maternal age reporting will avoid masking the increased risk to children born to very young adolescent mothers. Collection of additional biological and social data may better reveal mediators of this relationship. Targeted intervention strategies to reduce unintended pregnancy at earlier ages may also improve child survival.

**What is already known?:**

- Most previous studies treat under-20 mothers as a single group when looking at risk of child health outcomes.
- Few studies have assessed the risk gradient versus age within this group, focusing only on neonatal and infant mortality rather than broader child survival outcomes.
- These studies found a higher risk of neonatal and infant mortality among younger adolescent mothers, even after adjusting for socio-economic, demographic and health service accessibility variables.
- The risk gradients for stillbirths and under-5 mortality outcomes of children born to adolescent mothers remain unexplored.

**What are the new findings?:**

- This is the most comprehensive, multi-regional study to-date that investigated the potential impacts of adolescent pregnancy, examining multiple child survival endpoints from stillbirths to under-5 mortality, and quantifying the risk gradient as a function of maternal age from adolescence through young adulthood.
- Children of mothers younger than 16 faced 2-4 times higher risk of death at all child mortality stages (stillbirths, neonatal, infant, and under-5) in both sub-Saharan Africa and South Asia regions.
- The association extends across socio-economic status (SES) groups in both urban and rural settings and stays consistent when controlling for maternal education and health seeking risk factors.

**What do the new findings imply?:**

- We recommend revision of maternal-age-group reporting conventions to make the increased child survival risk with adolescent pregnancy more visible.
- To improve child survival outcomes, improving health-seeking behavior and quality of maternal care, as well as targeted interventions to reduce unintended pregnancy among adolescents and mitigate its harmful consequences are needed.
- Collecting additional data on the social and biological aspects of adolescent pregnancy could help understand the impact size of these mediators on child health outcomes.

## Introduction

Every year, nearly 12 million adolescent girls and young women aged 15–19 years and nearly a million under 15 years give birth.^1^ The majority of these births are in low- and middle-income countries (LMICs).^2^ The adolescent fertility rate (birth rate per 1,000 girls and young women aged 15-19 years) over the period 2015-2020 was the highest in the Sub-Saharan Africa (SSA) region at 102.8 births per 1,000 person-years, far higher than the global average (44 per 1,000), followed by South Asia with 26 births per 1,000 girls aged 15-19.^3^

Adolescence is a unique stage of human development and an important time for building the foundation of good health; consequently, pregnancy during this lifestage can have impacts on both a young woman and her children. Early pregnancy can lead to devastating health consequences for the mother, since adolescent girls may not yet be physically and biologically ready for pregnancy or childbirth.^3^ Many adolescents experience complications during pregnancy and childbirth, which has become the leading global cause of death among 15-19 years old females.^4^ Pregnant adolescents are at a higher risk of receiving inadequate antenatal care in some settings.^5^ A significant proportion of adolescents in SSA do not access nor utilize maternal services during pregnancy, which is a consequence of several individual, interpersonal, institutional, and systemic factors.^6^ Early pregnancy and motherhood for an adolescent girl in some contexts can also have adverse social consequences such as stigma and dropping out of school.^1,7^ They may not have the opportunity to return to school which jeopardizes their economic and employment opportunities due to their double burden of household maintenance and child-rearing,^7,8^ resulting in sustained poverty and increased vulnerability.

Reduction of adolescent pregnancy has long been the focus of several organizations and is of current policy interest. In fact, with only eight years left to achieve the 2030 Agenda for Sustainable Development, agreed to by more than 190 countries, there remains a timely commitment and need to ensure access to sexual and reproductive healthcare services, particularly for adolescent girls and young women (Target 3.7), and eliminate child, early, and forced marriage (Target 5.3), given their strong associations with adolescent pregnancy and its outcomes.^9^ Despite these efforts and the recent decline in overall adolescent mortality^10^ and global adolescent fertility rate, prevalence of adolescent pregnancies remains high and a major public health concern, especially in LMICs.

In standard surveys, reports, and World Health Organization (WHO) statements, mothers under 20 are usually treated as a single group.^11^ However, adolescence represents a time of developmental transition, including physically, cognitively, and psychologically, and there are substantial differences across the 10 to 19 years age range.^12^ Few studies have looked at the risk gradient versus age among young mothers. Several studies have associated early maternal age with neonatal and infant mortality,^2,9,13,14^ infant stunting, and preterm birth even after adjustment for socio-demographic factors.^15^ In contrast, two recent multi-country studies did not find a consistent significant association between adolescent motherhood and stillbirth.^16,17^ Current findings and studies leave unanswered questions about the true nature of these relationships.

A meta-analysis of Demographic Health Surveys (DHS) showed higher risk of mortality to neonates born to mothers aged < 16 and 16-17 years old than neonates born to mothers aged 20-29 years in SSA and South and Southeast Asia,^11^ even after adjusting for socio-economic, demographic and health service utilization variables. In LMICs, the infant mortality rate was higher among mothers with ages of 12-14 and 15-17 years than among older mothers.^13^ Finlay et al^14^ showed in a separate analysis that the risk of infant mortality in SSA is highest for high parity young mothers, and short birth intervals negatively affect infant mortality and stunting outcomes. A WHO multi-country study divided mother ages into <16, 16-17, 18-19, and 20-24 years old. They found stillbirth rates among adolescent mothers to be mildly higher than 20-24 years old mothers (odds ratios 1.0-1.3), with the difference significant only for the 16-17 years old group. ^17^ A more recent study examined the association between maternal age, both young and advanced, and risk of neonatal mortality in LMICs using DHS data, and found the risk of mortality of neonates born to mothers aged 12-15 and 45+ years was higher than neonates born to mothers aged 25-29 years.^18^ A systematic review and meta-analysis in SSA found that most evidence about the effects of early childbearing was for mothers 15-19 years old as a single group, with very few studies providing data on adolescents aged <18, and concluded that there is a lack of high quality observational studies that adjust for sociodemographic factors.^19^ Overall, there are limited number of studies focusing on risk gradient versus maternal age among young mothers, and majorities of these studies focused on neonatal and infant mortality rather than broader child survival outcomes.

In our study, the most comprehensive of its kind to date, we have investigated the potential impacts of adolescent pregnancy on a substantially broader scope than previous studies, examining child mortality endpoints from stillbirths to under-5 mortality, and quantifying the risk gradient as a function of age from adolescence through young adulthood. In contrast to prior studies which focused mostly on survival endpoints around birth, we hypothesized that since adolescent mothers face greater physical, emotional, and social challenges, the impact on their offspring’s survival might be felt throughout early childhood. In addition, to examine whether observed associations between maternal age and child survival may be caused by confounding variables that affect both, we explored adjustment for key demographic variables such as urban vs. rural residence. We also investigated whether the association between mother’s age and child mortality endpoints might be mediated by maternal health-seeking. We focused on SSA, the region with the highest adolescent pregnancy and child mortality burdens, as well as South Asia, the second-highest region in child mortality burden where adolescent pregnancy rates fell rapidly in recent years.The comprehensiveness and multi-regionality of our analysis helps frame the inconsistent findings from previous studies^9,13-17^ on the relationship between maternal age and child health outcomes. Disaggregation of the adolescent age group helps highlight the increased risk of younger adolescents and the potential benefits of providing health services for these girls.

## Methods

### Data Source and Study Population

We analyzed DHS data collected between 2004 and 2018 from countries in SSA and South Asia. DHS are cross-sectional nationally representative large-scale household surveys that collect and analyze demographic, health, and nutrition data, in a manner that enables comparisons across countries and over time. ^20^ The women’s questionnaire, used in this study, invites all women aged 15-49 in a surveyed household to respond. In a few surveys, the target group was women and girls aged 10-49 years old. We defined three time periods: 2004-2008, 2009-2013, and 2014-2018, to assess the variation in each outcome over time. We estimated the risk gradient versus age at first birth among adolescent and young adult mothers. We considered only first births in order to avoid the various confounders associated with parity, and also because most adolescent births are first births. Since the vast majority of first births in the two regions take place by women’s mid-20s, we restricted the analysis to mothers 25 years old or younger. In total, 35 countries with 80 surveys in SSA and 11 countries with 27 surveys in South Asia were included in the analysis (Supplementary Material).

### Endpoints and Risk Factors

Maternal age at first birth within the ten years preceding the survey was divided into five groups: <16, 16-17, 18-19, 20-22, and 23-25 years old. For stillbirth, DHS only collects data from the five years preceding the survey. The age group <16 includes mothers aged 10-15 years old, or 15 years old in surveys restricted to women aged 15-49. Risk factors accounted for in this analysis are socio-demographic factors: urban/rural residency and maternal education status (dichotomized as any education vs. none); economic factors: wealth quintile (a country-specific measure of the household wealth compared to other households in each survey and grouped as poorest, poorer/middle/richer, and richest in our analysis); and health-seeking factors: place of delivery (at home vs. health facility) and antenatal care (ANC) utilization (no ANC visit vs. any ANC visit). When we used literacy instead of education, we found similar results and thus we did not include it in our analysis. All risk factors were coded as categorical variables in our analysis.

We examined the following outcomes in our study: stillbirth (pregnancies that lasted seven or more months and terminated in fetal death), neonatal mortality (death after a live birth within the first 28 days of life), infant mortality (death within the first year of life), child mortality (death after the first year and before reaching the age of five years), 1-59 month mortality (death after the first month and before reaching the age of five years), and under-5 mortality (death before reaching the age of five years). Of these, stillbirths, neonatal, infant and under-5 mortality are reported in the main article, and the remaining endpoints in Supplementary Material.

### Statistical Analysis

Descriptive analyses included calculating the neonatal, infant, child, and under-5 and 1-59 month mortality rates per 1,000 live births and their sampling errors based on the DHS mortality rates estimation methodology, a synthetic cohort life table.^21^ We calculated the mortality rates for three time periods in each region and for each maternal age group combined with selected risk factors. Among multiple births (twins, triplets, etc.), only those with the birth assigned order number of 1 by DHS were considered in the analysis. While excluding the remaining multiple-birth siblings reduces the sample size by about 1%, it helps simplify and stabilize the analysis by avoiding the need to account for another level of dependence.

A mixed effect logistic regression model was applied to each time period and each region separately, to examine the association between each of the outcome variables and the risk factors. The sample size for the maternal age group and sociodemographic factors are the same and therefore, these models are layered stepwise and comparable. A model including only age group and the random effects was developed, called here Model 0. The model was further adjusted for different combinations of risk factors, based on prior knowledge of common risk factors for LMIC child mortality, as well as to assess the impact of leading healthcare access and socioeconomic indicators on the risk gradient. In Model 1, maternal age at first birth, urban/rural residency, and maternal education status were included. Questions about place of delivery and antenatal care utilization were available for the last births in the three/five years preceding the survey. A model adjusting for maternal age and healthcare accessibility factors was developed, called here Model 2. All the first births that happened within the three/five years preceding the survey were included in Model 2. Given that the Primary sampling units (PSUs), year of survey, and country name were used as nested random effects in the model, the wealth index represents the deviation of household wealth from its own country’s mean wealth at the time of interview ^11^ and Model 1 was further adjusted for the wealth index, called here Model 3. We also combined the two health care variables into one and grouped the outcomes as having either ANC visit or facility birth; both ANC visit and facility birth; and no ANC visit and home delivery, and further adjusted Model 1 for this variable, called here Model 4. Note that despite including Model 1’s variables, Model 4 is not nested within Model 1 as reported here, because the health-seeking variables were only available for each mother’s last birth. Together with the first-birth constraint of all models, this reduces the sample size by 5-7 times, as well as weights the sample more towards recent first births. Models 0-2 are reported in the main article, and Models 3 and 4 in the supplementary material. Models 2 and 4 are layered stepwise and comparable, separately from models 0, 1, and 3. Each survey’s sample weights were used as the model prior weights in the fitting process. Mothers of age 23-25 years old were considered the reference age group. All the analyses were performed in R 4.0.^22^

### Patient and Public Involvement statement

Study participants or the public were not involved in the design, or conduct, or reporting, or dissemination plans of our research.

## Results

### Univariate Maternal-Age Risk Gradient

The numbers of first births and neonatal, infant, child, and under-5 deaths to women aged under 25 years old that occurred within ten years preceding the survey, grouped by region, survey period, and age, are given in online supplemental Tables 1 and 2 in the supplement. Within each time period, about 20-23% of first births to mothers 25 years old or younger in SSA were attributed to mothers under 18 years old. In South Asia, this proportion was lower and decreased from 15.7% in 2004-2008 to 8.1% in 2014-2018.

**Table 1.** Mortality rates of different child health outcomes and their sampling errors in Sub-Saharan Africa and South Asia

| Sub-Saharan Africa |  |  |  |  |  |  |  |  |  |
| --- | --- | --- | --- | --- | --- | --- | --- | --- | --- |
|  | 2004-2008 |  |  | 2009-2013 |  |  | 2014-2018 |  |  |
|  | NNMR<sup>a</sup> | IMR<sup>b</sup> | U5MR<sup>c</sup> | NNMR | IMR | U5MR | NNMR | IMR | U5MR |
| Maternal age at first birth (years) |  |  |  |  |  |  |  |  |  |
| < 16 | 85.1 ± 5 | 152.8±6.5 | 238.5±7.5 | 63.8±4.0 | 117.7±5.3 | 186.2±6.2 | 54.5±4.5 | 95.9 ± 5.5 | 156.5 ± 6.6 |
| 16-17 | 58.3 ± 2.3 | 115.8±3.1 | 188.3±3.9 | 49.8±1.9 | 91.3±2.6 | 145.2±3.3 | 44.4±2.0 | 72.5 ± 2.6 | 112.8 ± 3.2 |
| 18-19 | 46.9 ± 1.8 | 89±2.3 | 146±2.9 | 39.1±1.4 | 73.8±1.9 | 115.5±2.3 | 34.4±1.4 | 58 ± 1.8 | 88.2 ± 2.2 |
| 20-22 | 39.9 ± 1.4 | 79.3±2.1 | 127.3±2.6 | 35.8±1.2 | 62.4±1.6 | 98.6±2.0 | 29.8 ±1.2 | 50.3 ± 1.5 | 74.6 ± 1.8 |
| 23-25 | 40.2 ± 2.1 | 74.5±2.8 | 123.5±3.7 | 33.7±1.8 | 55.7±2.2 | 88.8±2.7 | 28±1.6 | 44.5 ± 1.9 | 64.8 ± 2.4 |
| Urban/Rural Residency |  |  |  |  |  |  |  |  |  |
| < 16 | 53.2 ± 2.9<br>92.6 ± 13.7 | 115.6 ± 6.1<br>161.6 ± 13.1 | 182.6 ± 10<br>251.9 ± 10.5 | 58.3±16.2<br>65.8±1.0 | 108.1±16.2<br>121.3±1.9 | 159.1±15.5<br>196.1±3.9 | 43.2 ± 3.1<br>58.6 ± 18.7 | 74.1 ± 9<br>103.6 ± 18.3 | 115.9 ± 10.6<br>170.9 ± 16.8 |
| 16-17 | 50.2 ± 2.4<br>60.7 ± 2.1 | 97.6 ± 2.3<br>121.2 ± 3.1 | 160.4 ± 2.3<br>196.8 ± 6.9 | 30.2±3.2<br>57.8±2.0 | 63.7±3.2<br>102.5±2.3 | 100.1±3.4<br>163.6±2.8 | 39 ± 13.2<br>46.3 ± 1 | 62.4 ± 13.9<br>76.2 ± 1.7 | 90.6 ± 13.6<br>121.2 ± 2.3 |
| 18-19 | 39.2 ± 2.4<br>49.8 ± 2.7 | 75.2 ± 2.5<br>94.1 ± 2.9 | 119.2 ± 2.9<br>156 ± 3.4 | 33.5±1.8<br>41.6±2.0 | 59.9±2.0<br>80.3±2.8 | 87.8±2.7<br>128.4±3.6 | 25.9 ± 3.2<br>37.9 ± 3.1 | 45.2 ± 4<br>63.5 ± 3 | 67.5 ± 4.1<br>97.1 ± 3 |
| 20-22 | 30 ± 1.8<br>44.2 ± 1.2 | 60.6 ± 1.8<br>87.4 ± 1.3 | 95.2 ± 1.9<br>141.4 ± 1.8 | 30.7±0.3<br>38.7±1.4 | 49.5±0.7<br>69.6±3.3 | 72.9±1.3<br>113.0±4.5 | 25.5 ± 0.9<br>32.2 ± 1.4 | 41.3 ± 1.5<br>55.4 ± 1.5 | 57.9 ± 4.5<br>84 ± 1.5 |
| 23-25 | 30.2 ± 0.5<br>46.4 ± 1.3 | 56.9 ± 2<br>85.4 ± 2.3 | 83.3 ± 2.1<br>147.3 ± 4.4 | 32.4±0.3<br>34.7±4.3 | 47.9±0.9<br>62.0±4.1 | 72.4±1.2<br>101.7±4.8 | 27.6 ± 0.4<br>28.4 ± 0.2 | 40.5 ± 0.5<br>48 ± 0.4 | 55.4 ± 0.8<br>72.8 ± 0.6 |
|  | 2004-2008 |  |  | 2009-2013 |  |  | 2014-2018 |  |  |
|  | NNMR | IMR | U5MR | NNMR | IMR | U5MR | NNMR | IMR | U5MR |
| Maternal Education Status: Any Education/No Education |  |  |  |  |  |  |  |  |  |
| < 16 | 69.5 ± 0.9<br>95.2 ± 15.7 | 124.7 ± 1.6<br>170.8 ± 15.7 | 185.3 ± 1.8<br>271.5 ± 13.8 | 59.5 ± 12.8<br>67.6 ± 1.3 | 112.7 ± 12.2<br>122.1 ± 3.4 | 169.8 ± 11.7<br>199.1 ± 5.3 | 55.4 ± 10.8<br>53.4 ± 16.8 | 93 ± 11.9<br>99.2 ± 17.3 | 140.4 ± 12.1<br>174.5 ± 15.8 |
| 16-17 | 51.6 ± 0.4<br>64.9 ± 2.8 | 98.6 ± 0.8<br>132.6 ± 5.1 | 150.2 ± 1.2<br>224.7 ± 9.4 | 42.1 ± 0.4<br>60.3 ± 3 | 77.4 ± 0.8<br>110 ± 3.3 | 121.6 ± 1.3<br>174.7 ± 3.6 | 40.8 ± 8.7<br>50.6 ± 0.7 | 67.3 ± 9.1<br>81.4 ± 1.1 | 97.7 ± 8.8<br>136.3 ± 1.7 |
| 18-19 | 37.7 ± 2.2<br>61.6 ± 4.5 | 74.3 ± 2.1<br>112.1 ± 4 | 117.8 ± 2.2<br>189 ± 4 | 35 ± 0.3<br>46.4 ± 3.8 | 65.9 ± 2.3<br>87.8 ± 5 | 98.5 ± 3.6<br>143.3 ± 5.4 | 31.5 ± 2<br>41.1 ± 6.7 | 52.3 ± 2<br>71.2 ± 6.4 | 76.4 ± 2.3<br>113.4 ± 6.4 |
| 20-22 | 31.8 ± 0.2<br>56.4 ± 2.4 | 64.3 ± 0.4<br>109.2 ± 2.4 | 100.8 ± 0.6<br>178.7 ± 3.6 | 32.5 ± 0.3<br>42.2 ± 2.6 | 56.2 ± 0.5<br>74.4 ± 5.8 | 84.3 ± 1.9<br>124.2 ± 7.6 | 27.8 ± 0.7<br>35.1 ± 3 | 45.1 ± 1.1<br>64.2 ± 2.8 | 65.1 ± 1.6<br>97.9 ± 2.6 |
| 23-25 | 35 ± 0.4<br>52.3 ± 2.3 | 65.1 ± 0.7<br>96.3 ± 4.2 | 102.1 ± 0.9<br>169.9 ± 7.5 | 32.1 ± 0.2<br>37.1 ± 6.7 | 51.1 ± 0.4<br>65.2 ± 6.5 | 78 ± 0.5<br>109.6 ± 7.6 | 28.1 ± 0.3<br>27.9 ± 0.1 | 43 ± 0.5<br>48.9 ± 0.1 | 58.7 ± 0.7<br>81.1 ± 0.2 |
| Antenatal Care Visit (ANC)<sup>d</sup>: Any Visit/No Visit |  |  |  |  |  |  |  |  |  |
| < 16 | 33.5 ± 1.6<br>38.5 ± 3.6 | 79.9 ± 4<br>53.2 ± 5 | 187.4 ± 15<br>141.3 ± 16.2 | 26.7 ± 18.6<br>81.9 ± 16.9 | 60.7 ± 22.3<br>122.1 ± 20.7 | 109.3 ± 30.8<br>209.1 ± 47 | 27.9 ± 3.6<br>26.1 ± 1.8 | 43.9 ± 5.7<br>85.2 ± 17.6 | - |
| 16-17 | 27 ± 1<br>57.4 ± 2 | 60.2 ± 2.2<br>93.6 ± 10.6 | 94.2 ± 11.1<br>173.1 ± 11.8 | 24.8 ± 0.5<br>28.8 ± 14.7 | 51.6 ± 1.1<br>64.3 ± 14.4 | 93 ± 2<br>95.8 ± 14.2 | 24.3 ± 0.7<br>41 ± 1.5 | 39.8 ± 1.1<br>79.2 ± 2.8 | 64.8 ± 1.3<br>41 ± 5.2 |
| 18-19 | 25.7 ± 0.7<br>0.8 ± 1.7 | 48.5 ± 1.4<br>77.3 ± 3.3 | 80.8 ± 2.4<br>132.6 ± 3.6 | 23.7 ± 0.4<br>36.6 ± 1.3 | 47.8 ± 3.8<br>60.3 ± 7.6 | 87.9 ± 3.7<br>176.2 ± 11.1 | 19.4 ± 3.7<br>38.3 ± 1.3 | 32.1 ± 3.5<br>62.8 ± 2.3 | 52.4 ± 3.3<br>103.9 ± 3.4 |
| 20-22 | 22.8 ± 0.4<br>53 ± 2 | 48.5 ± 0.7<br>101.7 ± 4 | 77.3 ± 1.1<br>206.9 ± 18.9 | 22.6 ± 0.4<br>6.2 ± 15.3 | 41.1 ± 0.6<br>74.5 ± 18.2 | 59.8 ± 0.8<br>150.1 ± 19.7 | 15.9 ± 0.2<br>42.8 ± 1.6 | 27.4 ± 0.3<br>82.1 ± 2.9 | 38.2 ± 0.4<br>125.4 ± 4.2 |
| 23-25 | 26.1 ± 0.7<br>24.7 ± 1.2 | 52.3 ± 1.4<br>43.1 ± 15.4 | 82.7 ± 1.9<br>132.2 ± 248.3 | 21.3 ± 0.3<br>20.6 ± 1.2 | 36.8 ± 0.4<br>44.9 ± 2.7 | 65.4 ± 0.5<br>63.1 ± 3.8 | 16.5 ± 0.9<br>15.2 ± 1.2 | 24.9 ± 1<br>52.3 ± 4.5 | 34.1 ± 1.4<br>96.1 ± 9.9 |
|  | 2004-2008 |  |  | 2009-2013 |  |  | 2014-2018 |  |  |
|  | NNMR | IMR | U5MR | NNMR | IMR | U5MR | NNMR | IMR | U5MR |
| Place of Delivery<sup>d</sup>: At Home/Health Facility |  |  |  |  |  |  |  |  |  |
| < 16 | 88 ± 8.8<br>59.8 ± 19 | 161 ± 9.5<br>94.2 ± 21.5 | 253.1 ± 11.6<br>188.8 ± 31.2 | 65.5 ± 2.9<br>38.6 ± 31.1 | 116.9 ± 5.3<br>73.6 ± 31.3 | 214.3 ± 11.9<br>138.2 ± 29.2 | 53 ± 26.4<br>41.1 ± 2.1 | 105.7 ± 25.1<br>78.6 ± 11 | 215.4 ± 22.2<br>140 ± 11.9 |
| 16-17 | 53.8 ± 1.2<br>41.7 ± 3 | 108.5 ± 2.5<br>88.6 ± 3.9 | 187.3 ± 5.4<br>138.1 ± 9 | 56.3 ± 1.5<br>32.9 ± 0.6 | 90.3 ± 2.5<br>68.1 ± 1.3 | 156.1 ± 5.5<br>114.7 ± 2.6 | 51.2 ± 1.1<br>34.7 ± 1 | 80.8 ± 1.7<br>60.1 ± 1.6 | 150.9 ± 3.1<br>91.6 ± 1.8 |
| 18-19 | 46.5 ± 5.2<br>35 ± 1.1 | 92.5 ± 5<br>65.1 ± 2 | 148.6 ± 7.2<br>100.9 ± 2.9 | 41.5 ± 8.4<br>32.2 ± 0.6 | 78.9 ± 7.7<br>59.8 ± 1.1 | 132.1 ± 7<br>94.4 ± 1.8 | 41.2 ± 7.5<br>30.9 ± 0.7 | 72.5 ± 6.7<br>46.9 ± 1 | 117 ± 6.1<br>68.2 ± 1.7 |
| 20-22 | 43.5 ± 0.7<br>33.3 ± 0.6 | 90.3 ± 1.4<br>65.2 ± 1.1 | 141.8 ± 2.8<br>100.7 ± 1.7 | 44.2 ± 1.3<br>31.1 ± 0.6 | 70.8 ± 2.2<br>53.2 ± 1.1 | 110.4 ± 4.1<br>79.6 ± 1.8 | 38.2 ± 6.7<br>27.6 ± 0.3 | 70.6 ± 6.5<br>43.4 ± 0.4 | 104.9 ± 6.5<br>57.9 ± 0.4 |
| 23-25 | 45.9 ± 4.4<br>31.7 ± 0.6 | 83.6 ± 7.1<br>60.2 ± 1.2 | 140.6 ± 6.9<br>97.2 ± 1.7 | 30.3 ± 0.9<br>30.7 ± 0.3 | 60.4 ± 1.7<br>47.9 ± 0.4 | 96.7 ± 21.3<br>75.2 ± 0.4 | 25.4 ± 0.8<br>29.5 ± 0.6 | 48.3 ± 1.6<br>41.3 ± 0.8 | 74.5 ± 2.9<br>53.6 ± 1.1 |
| South Asia |  |  |  |  |  |  |  |  |  |
|  | 2004-2008 |  |  | 2009-2013 |  |  | 2014-2018 |  |  |
|  | NNMR | IMR | U5MR | NNMR | IMR | U5MR | NNMR | IMR | U5MR |
| Maternal age at first birth (years) |  |  |  |  |  |  |  |  |  |
| < 16 | 82.9 ± 1.6 | 112.6 ± 2.1 | 145 ± 2.5 | 55.4 ± 2.7 | 81.7 ± 3.1 | 98.9 ± 6.4 | 59.7 ± 15 | 84.4 ± 20.8 | 95.7 ± 22.8 |
| 16-17 | 65.6 ± 2.3 | 94.8 ± 5 | 112.1 ± 4.9 | 50.8 ± 1.1 | 75 ± 2.8 | 86.9 ± 2.9 | 46.4 ± 3.7 | 64.4 ± 5.1 | 74 ± 5.7 |
| 18-19 | 53.2 ± 0.7 | 74.3 ± 1.1 | 86.5 ± 1.2 | 45.7 ± 1.1 | 61.6 ± 1.3 | 71 ± 2.3 | 38.7 ± 7.2 | 51.4 ± 7.3 | 58.7 ± 8.5 |
| 20-22 | 39.6 ± 1.1 | 56.4 ± 1 | 66.9 ± 1 | 32.5 ± 1.5 | 45.7 ± 1.5 | 53.6 ± 1.5 | 31.3 ± 4.6 | 41.9 ± 4.6 | 48.3 ± 4.6 |
| 23-25 | 35.5 ± 0.2 | 49.6 ± 0.3 | 57.5 ± 0.4 | 25.3 ± 0.1 | 36.4 ± 0.3 | 45.1 ± 0.3 | 26.3 ± 1.6 | 34.4 ± 3.3 | 40.7 ± 3.3 |

| South Asia |  |  |  |  |  |  |  |  |  |
| --- | --- | --- | --- | --- | --- | --- | --- | --- | --- |
|  | 2004-2008 |  |  | 2009-2013 |  |  | 2014-2018 |  |  |
|  | NNMR | IMR | U5MR | NNMR | IMR | U5MR | NNMR | IMR | U5MR |
| Urban/Rural Residency |  |  |  |  |  |  |  |  |  |
| < 16 | 80.2 ± 4.2<br>83.4 ± 1.6 | 107.3 ± 5.6<br>113.6 ± 2.1 | 132.1 ± 6.5<br>147.6 ± 2.5 | 48.8 ± 6.2<br>57.3 ± 2.8 | 63.5 ± 12<br>87.1 ± 4.2 | 86.4 ± 11.8<br>102.6 ± 7.5 | 56.4 ± 33.5<br>60.6 ± 52.8 | 71.8 ± 42.2<br>88 ± 51.4 | 77.1 ± 44.1<br>101.2 ± 50.5 |
| 16-17 | 51.1 ± 1.7<br>69 ± 2.2 | 71.5 ± 2.4<br>100.3 ± 4.9 | 87.1 ± 3.2<br>118.1 ± 4.7 | 34.4 ± 3.1<br>55.7 ± 3.7 | 53.8 ± 4.8<br>81.2 ± 4.3 | 60.5 ± 4.8<br>94.4 ± 4.4 | 27.3 ± 4.5<br>52.8 ± 6.1 | 44.4 ± 7.3<br>71.1 ± 6.7 | 54.5 ± 8.6<br>80.5 ± 10.1 |
| 18-19 | 36.5 ± 0.4<br>58.2 ± 0.8 | 53.3 ± 0.5<br>80.7 ± 1.2 | 61 ± 0.6<br>4.4 ± 1.4 | 31 ± 1.6<br>50.7 ± 1.6 | 48 ± 2.1<br>66.1 ± 1.8 | 52.6 ± 3.4<br>77.1 ± 2.6 | 30.4 ± 15.2<br>41.5 ± 1.9 | 40.5 ± 15.3<br>55.1 ± 2.5 | 45.8 ± 17.8<br>63.2 ± 2.8 |
| 20-22 | 28 ± 0.2<br>44.8 ± 1.2 | 39.4 ± 0.3<br>64.1 ± 1.1 | 45 ± 0.4<br>76.8 ± 1 | 21.3 ± 0.7<br>37.7 ± 1.7 | 30.1 ± 0.7<br>52.9 ± 1.7 | 35.1 ± 1<br>62.4 ± 1.7 | 23.1 ± 8.8<br>34.6 ± 0.9 | 31.2 ± 8.7<br>46.3 ± 1.4 | 35.1 ± 8.7<br>53.6 ± 1.5 |
| 23-25 | 27.1 ± 0.2<br>41.4 ± 0.3 | 37.2 ± 0.2<br>58.2 ± 0.5 | 43.5 ± 0.3<br>67.3 ± 0.6 | 19.2 ± 0.1<br>9.4 ± 0.2 | 28.7 ± 0.5<br>41.5 ± 0.3 | 35.6 ± 0.5<br>51.5 ± 0.3 | 16.6 ± 2.5<br>31.9 ± 0.4 | 21.9 ± 5<br>41.6 ± 0.6 | 26.8 ± 5<br>48.8 ± 0.7 |
| Maternal Education Status: Any Education/No Education |  |  |  |  |  |  |  |  |  |
| < 16 | 67.4 ± 1.9<br>100.1 ± 0 | 95 ± 2.7<br>132 ± 0 | 120 ± 3.2<br>170.9 ± 0.4 | 42.9 ± 3.2<br>100.2 ± 4.6 | 62.7 ± 3.4<br>149.4 ± 6.7 | 82.6 ± 7.6<br>159.5 ± 6.9 | 67.2 ± 17.9<br>45.1 ± 8.3 | 85.7 ± 22.5<br>81.5 ± 14.4 | 96 ± 24.3<br>94.6 ± 16.9 |
| 16-17 | 52.7 ± 3.9<br>83.1 ± 0.8 | 76.8 ± 3.7<br>118.8 ± 5.2 | 88.3 ± 3.6<br>142.5 ± 5 | 40.3 ± 0.9<br>94.5 ± 1.6 | 61 ± 3.1<br>132.8 ± 2.2 | 68 ± 3.1<br>160.5 ± 2.6 | 42.1 ± 3.2<br>56.8 ± 7.1 | 57.4 ± 4.4<br>80.8 ± 9.9 | 65 ± 4.8<br>94.3 ± 11.1 |
| 18-19 | 44.2 ± 0.7<br>68.9 ± 0.5 | 61.6 ± 1.4<br>96.6 ± 0.7 | 70.7 ± 1.4<br>113.2 ± 0.8 | 39.8 ± 1.1<br>72.6 ± 0.8 | 52.6 ± 1.3<br>102.5 ± 1.1 | 61.5 ± 2.6<br>113.6 ± 1.2 | 35.4 ± 1.3<br>47.7 ± 20.4 | 45.7 ± 1.6<br>66.8 ± 20.5 | 51.6 ± 6.7<br>77.4 ± 20.4 |
| 20-22 | 32.4 ± 1.5<br>59.1 ± 0.2 | 46.3 ± 1.4<br>83.8 ± 0.3 | 53.5 ± 1.<br>102.1 ± 2.9 | 28.2 ± 1.7<br>57.5 ± 0.3 | 40.3 ± 1.7<br>76.7 ± 0.4 | 47.3 ± 1.7<br>89.5 ± 0.5 | 28.7 ± 0.9<br>39.9 ± 16 | 37.2 ± 1.2<br>56.9 ± 15.8 | 42 ± 1.3<br>67.4 ± 15.7 |
| 23-25 | 29 ± 0.2<br>63.8 ± 0.8 | 39.3 ± 0.3<br>93.8 ± 1 | 45.7±0.4<br>107.2 ± 1 | 22.4 ± 0.1<br>46 ± 0.6 | 31.8 ± 0.4<br>68.9 ± 1 | 39.3 ± 0.4<br>85.4 ± 1.2 | 22.8 ± 1.9<br>40.8 ± 2.3 | 28.7 ± 1.9<br>57.1 ± 6.4 | 33.8 ± 1.9<br>67.5 ± 6.6 |
|  | 2004-2008 |  |  | 2009-2013 |  |  | 2014-2018 |  |  |
|  | NNMR | IMR | U5MR | NNMR | IMR | U5MR | NNMR | IMR | U5MR |
| Antenatal Care Visit (ANC): Any Visit/No Visit |  |  |  |  |  |  |  |  |  |
| < 16 | 23.4 ± 0.8<br>62.4 ± 5.6 | 56.3 ± 2<br>63.8 ± 5.7 | 67.1 ± 2.4<br>65.8 ± 5.9 | - | - | - | 29.2 ± 2<br>40.3 ± 30.9 | 29.2 ± 2<br>40.3 ± 30.9 | 55.7 ± 7.8<br>40.3 ± 31 |
| 16-17 | 26.4 ± 0.5<br>29.1 ± 0.7 | 38.9 ± 0.7<br>65.2 ± 11.9 | 56 ± 1.3<br>69.8 ± 11.9 | 19.3 ± 2.1<br>27.1 ± 1.4 | 23.7 ± 2.2<br>32 ± 5.1 | 27.1 ± 7<br>39.2 ± 5.3 | 19 ± 2<br>39.8 ± 3.2 | 25.9 ± 2.8<br>67 ± 5.6 | 28.4 ± 3.2<br>83 ± 7.2 |
| 18-19 | 26.8 ± 0.4<br>25.6 ± 0.4 | 35.5 ± 0.5<br>31.6 ± 7.9 | 40.3 ± 0.6<br>34.7 ± 7.8 | 19.9 ± 0.4<br>20.7 ± 0.8 | 24.1 ± 0.4<br>39.6 ± 1.5 | 27 ± 0.5<br>47.8 ± 1.8 | 17.8 ± 1.2<br>38.4 ± 2.7 | 25 ± 1.7<br>59.4 ± 4.1 | 27.3 ± 1.9<br>64.8 ± 4.7 |
| 20-22 | 23.3 ± 0.2<br>38.2 ± 0.9 | 31.4 ± 0.2<br>52.4 ± 1.3 | 36.3 ± 0.2<br>75.1 ± 2.1 | 12.9 ± 3.6<br>24.1 ± 6.5 | 16 ± 3.6<br>37.9 ± 6.5 | 17.5 ± 3.6<br>39.3 ± 8.3 | 16.6 ± 0.7<br>34.6 ± 6 | 22.9 ± 0.9<br>46.6 ± 8.1 | 25.8 ± 1<br>54.7 ± 8.4 |
| 23-25 | 16.8 ± 0.2<br>56.6 ± 3.2 | 22.7 ± 0.2<br>81.8 ± 4.5 | 23.2 ± 0.2<br>89 ± 5.1 | 11.9 ± 0.1<br>39.2 ± 4.3 | 18.5 ± 0.7<br>44.9 ± 5 | 24.9 ± 0.7<br>44.9 ± 5 | 15.7 ± 3.4<br>36 ± 2 | 20.4 ± 3.4<br>52 ± 2.9 | 24 ± 3.5<br>70.2 ± 3 |
| Place of Delivery: At Home/Health Facility |  |  |  |  |  |  |  |  |  |
| < 16 | 57.3 ± 2.2<br>74.1 ± 6.1 | 87.3 ± 3.5<br>109.3 ± 8.7 | 120.7 ± 5.1<br>109.3 ± 8.7 | 28.1 ± 1.8<br>40.7 ± 40.4 | 34.9 ± 2.3<br>65.7 ± 39.7 | 50.3 ± 35.8<br>65.7 ± 39.7 | 53.7 ± 14.9<br>55.7 ± 2.6 | 79.4 ± 24.1<br>75.3 ± 3.6 | 112.5 ± 24.2<br>141.1 ± 9.2 |
| 16-17 | 61.2 ± 3.3<br>62 ± 1.8 | 91.9 ± 5.4<br>83.6 ± 2.4 | 103.9 ± 5.3<br>94.3 ± 2.6 | 36.3 ± 0.8<br>38 ± 0.8 | 56.2 ± 1.1<br>38 ± 0.8 | 64.3 ± 1.1<br>47.7 ± 1.2 | 53.9 ± 4<br>41.7 ± 3.5 | 80.4 ± 5.9<br>56.9 ± 4.9 | 88.2 ± 6.1<br>67.4 ± 6.5 |
| 18-19 | 50.4 ± 1.2<br>47.2 ± 0.6 | 69.9 ± 3.6<br>60 ± 0.7 | 89 ± 3.5<br>66.8 ± 0.8 | 48.5 ± 0.9<br>32.6 ± 0.9 | 58.5 ± 1.1<br>40.9 ± 1.2 | 68.7 ± 6.3<br>46.2 ± 1.4 | 43.4 ± 2<br>34.7 ± 3 | 64.4 ± 3.2<br>46.5 ± 4.1 | 70.1 ± 3.3<br>52 ± 4.7 |
| 20-22 | 43.4 ± 2.6<br>30 ± 0.3 | 61 ± 2.5<br>38.3 ± 0.4 | 78.1 ± 3.8<br>43.1 ± 0.4 | 30.4 ± 0.5<br>26 ± 0.2 | 41.5 ± 0.7<br>32.3 ± 0.2 | 49.7 ± 0.9<br>38 ± 0.2 | 46 ± 4.7<br>27.2 ± 1.9 | 60.9 ± 6.4<br>37.1 ± 2.6 | 79.1 ± 10.1<br>43.7 ± 3 |
| 23-25 | 46.9 ± 0.6<br>29.3 ± 0.4 | 68.2 ± 0.8<br>36.7 ± 0.5 | 76.6 ± 0.9<br>41.8 ± 0.5 | 29.2 ± 0.5<br>19.2 ± 0.1 | 43.5 ± 0.7<br>27.2 ± 1.4 | 53.8 ± 0.9<br>33.2 ± 1.4 | 42.4 ± 1.5<br>24.4 ± 3 | 58.1 ± 9.2<br>31.3 ± 3.2 | 73.2 ± 9.2<br>38.2 ± 3.3 |

<sup>a</sup> NNMR: neonatal mortality rate, <sup>b</sup> IMR: infant mortality rate, <sup>c</sup> U5MR: under-5 mortality rate, <sup>d</sup> births that occurred in the three/five years preceding the survey.

The majority of women across all ages and time periods lived in rural areas. About 60% and 43% of mothers aged <16 years old respectively, in SSA and South Asia, in the 2004-2008 period had no formal education. This rate decreased with age and time in SSA, but a similar trend was not observed in survey data from South Asia (online supplemental Table 2). In both regions, about 80% of women had at least one antenatal care visit. About 45% to 48% of pregnant women aged <16 years old in both regions gave birth at home during the 2014-2018 period, 2-3 times more often than mothers over 20 years old. The gap has not narrowed substantially between 2004-2008 and 2014-2018 (online supplemental Tables 1 and 2).

The mortality rates for different child outcomes and their sampling errors are given in Table 1 for SSA and South Asia and online supplemental Tables 3 and 4, respectively. In SSA, neonates, infants and children under-5 born to mothers aged <16 were at about two times higher risk of death (54·5±4·5, 95·9±5·5 and 156·5±6·6 deaths per 1000 live births, respectively, in 2014-2018) than those born to mothers aged 23-25 years old (28±1·6, 44·5±1·9 and 64·8±2·4, respectively). The mortality rates for children of the <16 age group in South Asia were two to three times higher (59·7±15, 84·4±20.8 and 95·7±22·8, respectively) than the oldest age group (26·3±1·6, 34·4±3·3 and 40·7±3·3, respectively), however the uncertainty intervals are wide for the youngest age group. Overall, mortality rates decreased with time, but the risk gradient versus age has remained similar.

In both regions, the risk gradient versus age appears in both rural and urban locations (Figure 1). Similar age gradients were observed when dividing mothers by maternal education status and other variables (Table 1)

**Figure 1.**
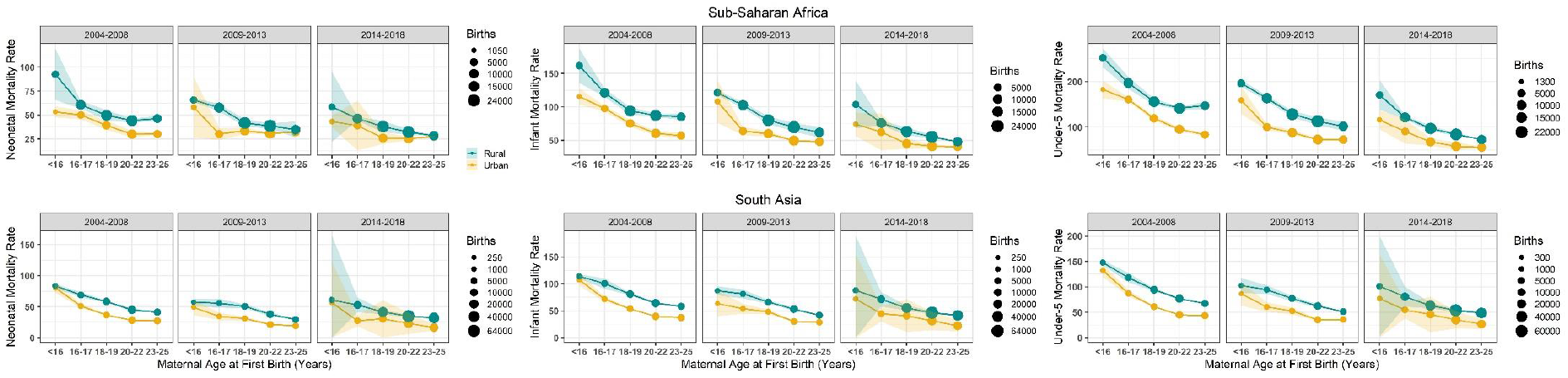
Neonatal (left), infant (center) and under-5 (right) mortality rates and their sampling errors within each age group and urban (yellow) and rural (green) locations in SSA (top) and South Asia (bottom) by 5-year time period; 2004-2008, 2009-2013, and 2014-2018. The circle size represents number of births within each group.

### Multivariate Analysis Adjusting for Risk Factors

Estimates from Models 0, 1 and 2 for the period 2014-2018 are shown in Figures 2-4. According to these models, adjusted maternal-age effects are consistent with the patterns of Table 1 and Figure 1. Mortality risk for all endpoints increases with younger age, and children in both regions born to mothers aged <16 faced 2-4 times higher mortality risk than those born to mothers aged 23-25 years old at all stages, from stillbirth to under-5 mortality (Figure 2), even after adjustment for demographic factors (Figure 3). The odds ratio (OR) for stillbirth is particularly high with under-16 mothers, around 4 or more in both regions and models. Adjusting for health-seeking variables reduced the age effect for neonatal and infant mortality but not for under-5 mortality in both regions (Figure 4 and online supplemental Figure 4). However, for stillbirths, the risk gradient versus age was stronger after adjustment for health-seeking, suggesting that some health-seeking recorded in the survey could be related to pregnancy complications or even to the stillbirths themselves. It should be emphasized that the dataset used in Figures 4 and S4 is smaller than the one used in Figures 1-3 since the healthcare variables were available only for the last birth in the three/five years preceding the survey. Further adjustment for wealth quintile did not modify the age effect significantly (Model 3, online supplemental Figure 3). For the 2009-2013 time period in SSA, similar patterns were observed (online supplemental Figures 5-9), and in South Asia where this time period had a particularly small sample size, the age effect was reduced. For the 2004-2008 time period, the risk gradient versus age appeared in both regions for the majority of child survival outcomes (online supplemental Figures 10-14).

**Figure 2.**
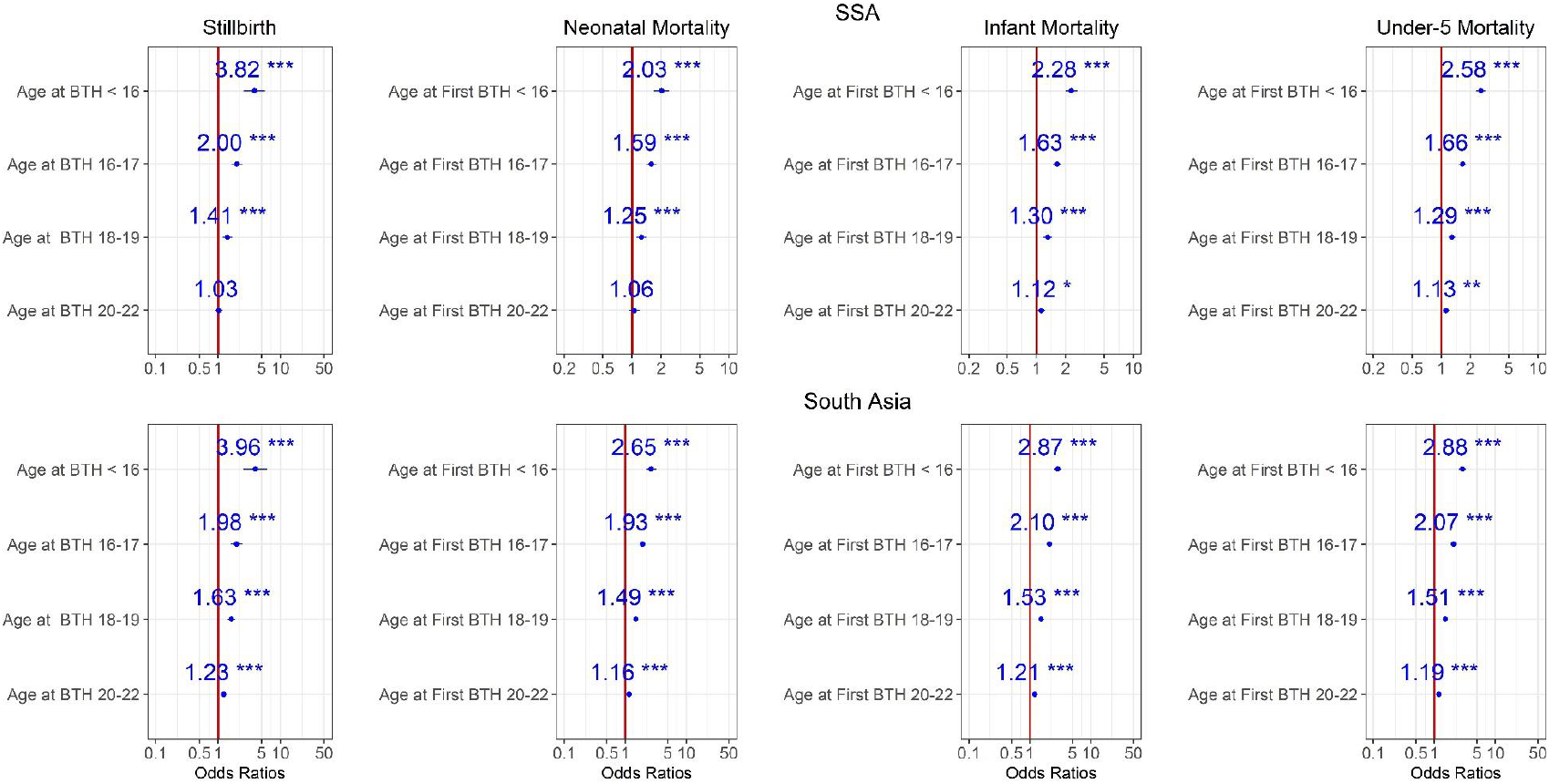
Ratios associated with neonate, infant, child, 1-59 months, under-5 years, and stillbirth in SSA and South Asia for the 2014-2018 survey period. Risk factors reducing the probability of death have odds ratios lower than 1 to the left of the vertical red line. Odds ratios (blue points), 95% confidence intervals (horizontal blue lines) are given. P-values are shown with the asterisk signs (‘***’ 0.001 ‘**’ 0.01 ‘*’ 0.05 ‘.’ 0.1 ‘’ 1). Reference group is mothers aged 23-25 years old (Model 0).

**Figure 3.**
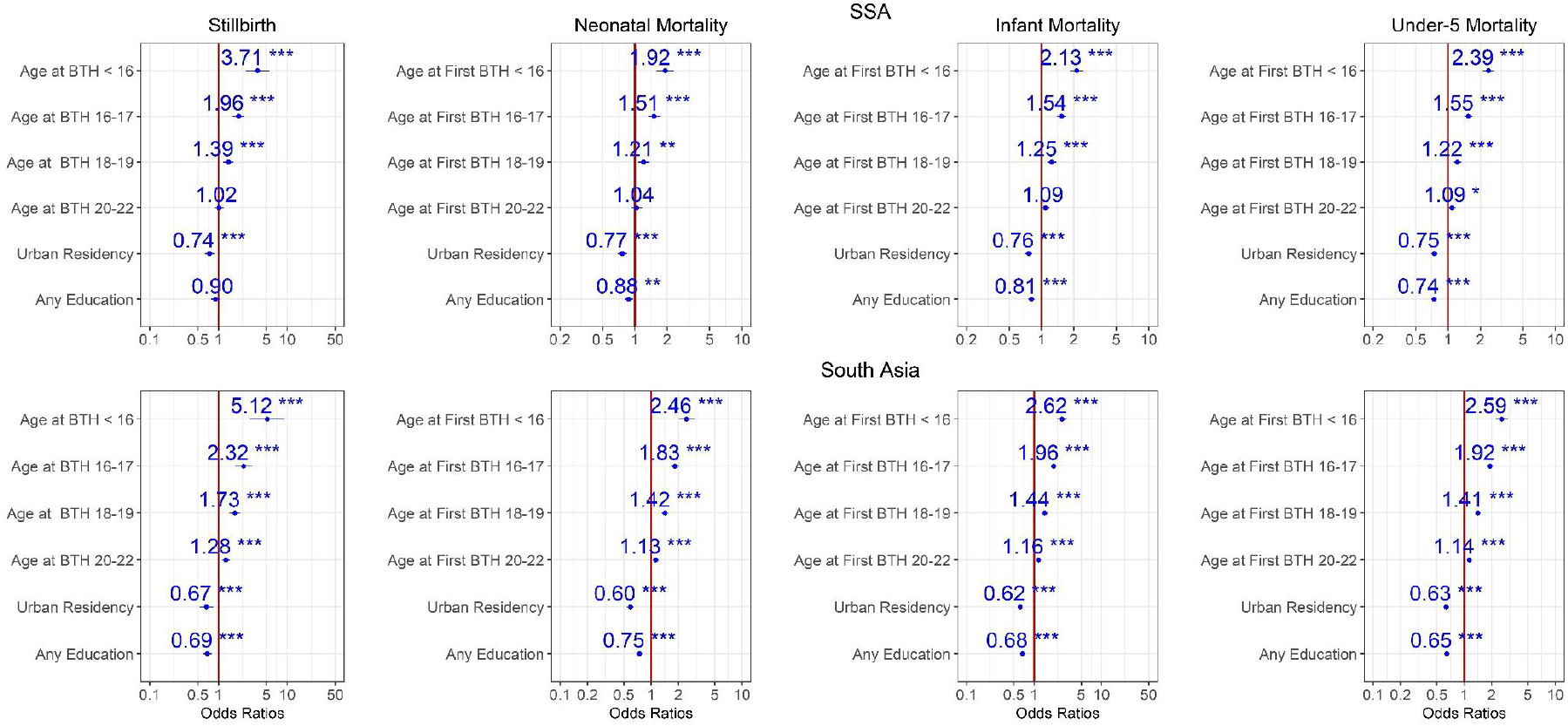
Ratios associated with neonate, infant, child, 1-59 months, under-5 years, and stillbirth in SSA and South Asia for the 2014-2018 survey period. Risk factors reducing the probability of death have odds ratios lower than 1 to the left of the vertical red line. Odds ratios (blue points), 95% confidence intervals (horizontal blue lines) are given. P-values are shown with the asterisk signs (‘***’ 0.001 ‘**’ 0.01 ‘*’ 0.05 ‘.’ 0.1 ‘ ‘ 1). Reference group is mothers aged 23-25 years old who live in rural areas and have no formal education (Model 1).

**Figure 4.**
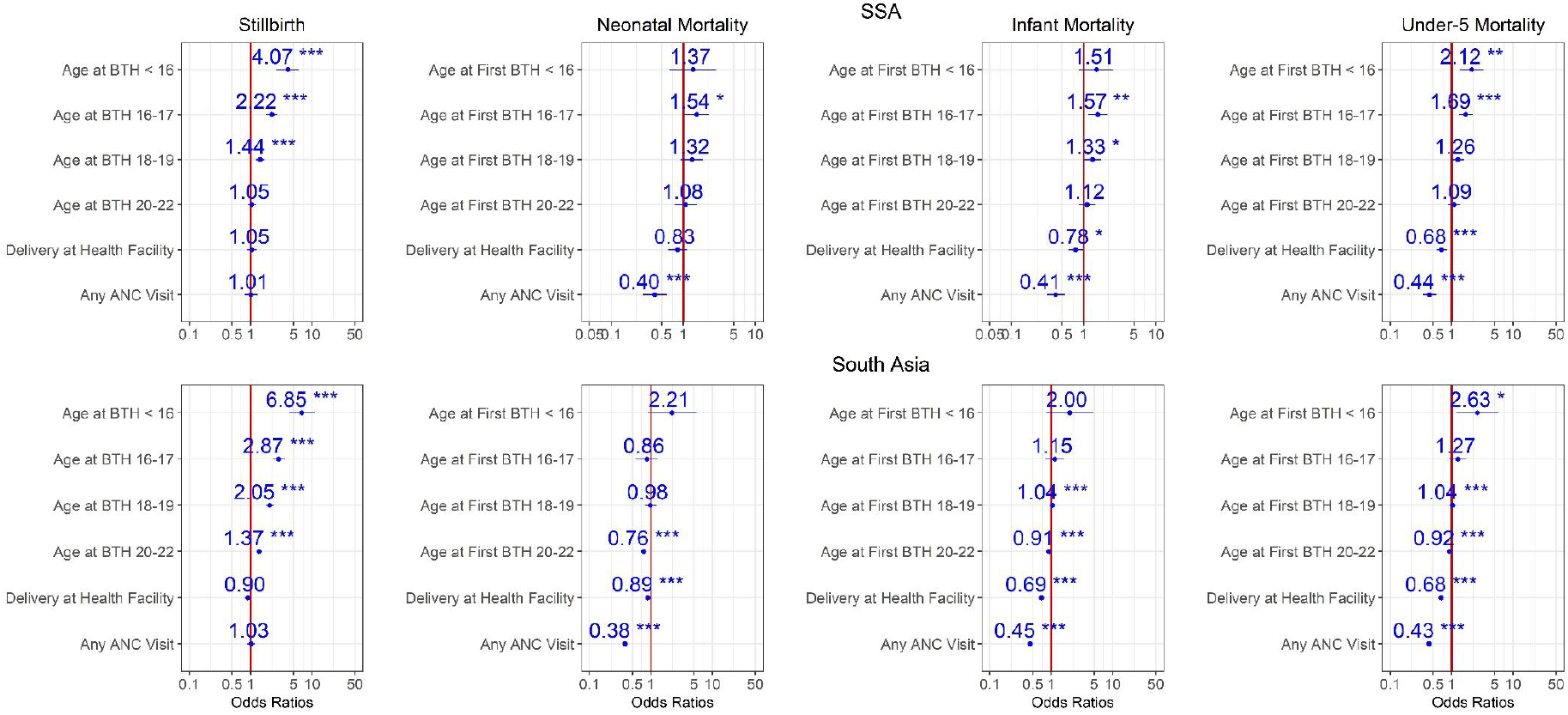
Ratios associated with neonate, infant, child, 1-59 months, under-5 years, and stillbirth in SSA and South Asia for the 2014-2018 survey period. Risk factors reducing the probability of death have odds ratios lower than 1 to the left of the vertical red line. Odds ratios (blue points), 95% confidence intervals (horizontal blue lines) are given. P-values are shown with the asterisk signs (‘***’ 0.001 ‘**’ 0.01 ‘*’ 0.05 ‘.’ 0.1 ‘ ‘ 1). Reference group is mothers aged 23-25 years old who delivered at home, and had no ANC visit (Model 2).

## Discussion

Among young mothers in SSA and South Asia, there was a consistent risk gradient versus maternal age at all stages of child mortality and all survey periods. Compared with other known risk factors, young maternal age appears to be among the strongest risk factors of child mortality. Our findings confirm and substantially expand the conclusions from previous studies about the association between early childbearing and adverse child health outcomes,^2,23, 24^ and suggest that the increased risk to children of younger mothers continues to linger even in regions with dropping adolescent pregnancy rates such as South Asia. Even in adjusted analyses, after controlling for several risk factors, the associations between adolescent pregnancy and child survival remained similar, except for neonatal and infant mortality where the effect was reduced after adjustment for health-seeking variables. This suggests that ensuring young mothers receive quality maternal care could reduce some of the early childbearing effects. Provision of antenatal and postnatal health services to adolescents can further be improved by recognizing their biological and social needs and vulnerabilities,^25^ considering that adolescents may experience social stigma from healthcare providers, besides the socioeconomic limitations they deal with. ^26^ The overall risk of death is higher among neonates, infants, and children of mothers in the poorest wealth quantile living in rural areas with no formal education, across all age groups. The heterogenous and limited progress in reducing adolescent pregnancies amongst these vulnerable groups emphasizes the inequity as well as inadequate distribution of resources and health services.^12,27^ However, the risk trend among younger mothers was evident across all SES groups, suggesting that beyond external social factors placing children of younger mothers at a higher level of disadvantage than other ages, underlying biological or behavioral immaturity of the mother were likely also at play.^16, 27^ Past studies have found that coinciding pregnancy with growth in young adolescents may lead to maternal-fetal competition for nutrients and consequently, to increased risk of low birth weight, neonatal mortality, and preterm delivery.^28, 29^

The high mortality risk of children under-5 highlights the likely long-term impact of limited education and livelihood opportunities for adolescent mothers.^30^ This can initiate a poverty cycle in their families, in addition to mental health and psychological challenges from social stigma that young mothers may deal with.^31,32^ To reduce neonatal and under-5 mortality rates towards Sustainable Development Goals (SDG) targets, it is necessary to focus efforts on reducing unintended pregnancies in adolescents in countries where they are prevalent, and ensure adolescent girls and young women have access to sexual and reproductive health services to both regulate their own fertility and prevent poor health outcomes for themselves and their children. Our analyses highlight where the risk is the highest and where we need to learn more in order to ensure girls, young women, and their children have more favorable health outcomes.

Previous studies have found strong associations between adolescent motherhood and child or early marriage, particularly in African and Asian contexts where marriage usually precedes childbearing.^18^ Moreover, early marriage before age 18 has been positively associated with higher fertility, poorer maternal and reproductive health, and poorer health and developmental outcomes among their children, through pathways including biological factors, social risks, and maternal behavior.^31,32^ Despite existing laws and human rights frameworks calling to eliminate marriage of girls before age 18, 650 million girls and women alive today married before their 18^th^ birthday; 40% of those women live in South Asia while 18% live in SSA.^33^ Strengthening measures to delay age at marriage may help reduce adolescent pregnancies in regions where both are strongly linked.

Our work has some limitations. There is likely under-reporting of mortality at early stages of child life, especially neonatal death and stillbirth. Further, survey responses rely on recall data,^2^ and respondents may overstate their ages at births during the interview due to social pressure. In addition, the survey data represent the socio-demographic characteristics of respondents at the time of interview, and not during the birth events. Besides the risk factors we accounted for in our study, other covariates related to the pregnancy-induced complications,^18^ which are not available in the DHS datasets, could help better explain the observed patterns. In addition, for the children under-5 mortality endpoint, other risk factors such as infectious diseases along with preterm birth complications, birth asphyxia, and trauma and congenital anomalies which are the leading causes of death for children under-5, were not included in this analysis due to lack of data. ^34^ The limited sample size of the group suffering the greatest disparities, under-16 mothers, constrained the number of variables we could consider in any single model, particularly in South Asia, and even more so with respect to health-seeking variables.

The limited sample size also required us to group all mothers under 16 in the main analysis. In sensitivity analysis, we divided the under-16 age group into two roughly equal groups of 10-14 and 15 years old in SSA for the unadjusted Model 0 as well as Model 1 and the period 2014-2018 (online supplemental Figures 15 and 16). Results showed a strong age effect for all child health outcomes with children of 10-14 years old mothers doing even worse than the combined under-16 group. However, the small sample size did not allow for further analysis. Similarly, after examining the maternal mortality DHS module we decided to not include maternal mortality in our analysis, due to the small sample size and lack of risk factors in this module, whose data are based on interviews with siblings of deceased mothers. All the above-mentioned limitations in retrospective analyses of cross-sectional survey data restrain the ability to disentangle the underlying biological, behavioral, and environmental mechanisms, and to rule out residual confounding factors. There is a need for longitudinal studies and follow-up data in diverse contexts to help tease apart the drivers of adverse child outcomes for young mothers.

## Conclusions

Our study highlights the strong differences in child health outcomes within the under-20 maternal age group, and provides quantitative evidence on the necessity for age-disaggregated reporting and survey data on adolescent pregnancy, given the specific biological and social risks to adolescent mothers and their babies,^25^ to better understand its associations with child outcomes and how their nature, scale, and impact vary by age. Revising the future classification of maternal age, and reporting of adolescent reproductive health will help better develop and monitor the progress of age-specific programs aimed at achieving the SDGs of reducing adolescent pregnancy.^35^ By building upon previous studies and policies, our work is more cognizant of empirical and health-seeking contexts, and suggests the path forward with respect to policy modification, while recognizing that adolescents biological and social needs and vulnerabilities should be accounted for when improving health services and developing age-specific policies. Building the capacity of adolescents to make their own decision and choices on their reproductive health, which is shaped by numerous social, cultural, and economic circumstances,^36^ is vital. Some of the contributing factors to this problem are beliefs, attitudes, and norms in the community as well as healthcare providers about adolescent sexuality that put them at risk for poor health outcomes.^26^ Among women who would want to avoid pregnancy in lower-middle-income countries (LMICs), the unmet need for modern contraception is much higher for adolescents than for all women aged 15-49. About 44% of adolescent women in LMICs who want to avoid pregnancy have an unmet need for modern contraception. ^36^ Interventions focused on expanding contraceptive access and use are key toward shifting social and gender norms at family and community levels,^31^ addressing early pregnancy, and subsequently improving child outcomes.^37^ Some age-specific programs could be related to increasing awareness on the importance of interventions, laws, and enforcement, and advocacy and outreach addressing individual and community barriers to delaying first pregnancy, including delaying marriage through establishing and enforcing laws,^38^ addressing underlying social and economic drivers and norms, empowering young women to choose if, when, and whom they marry, and enabling young women to continue and attain higher levels of education and to reduce unintended adolescent pregnancies as well as rapid repeat pregnancies.

## Data Availability

This study was based on secondary analysis of Demographic and Health Surveys (DHS) data. The ethical clearance was provided by the Institutional Review Board of ICF International. Therefore, this secondary analysis was exempt from ethical review approval, since it used publicly available, de-identified data.

## Contributors

JLP and APO helped develop the research concept and approach. NN analyzed the data and wrote the initial draft of the manuscript. YE and JLP provided subject-matter expertise and guidance. All authors contributed to the interpretation of results and writing of the manuscript and have read and approved the final manuscript.

## Acknowledgement

We would like to thank Dr. Guillaume Chabot-Couture and Dr. Edward Wenger for their useful and constructive comments and insights.

## Supplementary Material

Supplementary Material.pdf

