## Supplementary Material for "The Effect of Adolescent Pregnancy on Child Mortality in 46 Low- and Middle-Income Countries"

**Appendix 1.** List of countries in Sub-Saharan Africa (SSA) and South Asia with survey data
**Table 1.** Characteristics of the study population for the child health outcomes in SSA (Unweighted data). All numbers represent women's first birth out of their entire birth history, unless noted otherwise.
**Table 2.** Characteristics of the study population for the child health outcomes in South Asia (Unweighted data). All numbers represent women's first birth out of their entire birth history, unless noted otherwise.
**Table 3.** Child and 1-59 months mortality rates and their sampling errors in SSA
**Table 4.** Child and 1-59 months mortality rates and their sampling errors in South Asia
**Figure1.** Ratios associated with 1-59 months and child mortality in SSA and South Asia for the 2014-2018 survey period. Reference group is mothers aged 23-25 years old who live in rural areas and have no formal education (Model 1).
**Figure 2.** Ratios associated with 1-59 months and child mortality in SSA and South Asia for the 2014-2018 survey period. Reference group is mothers aged 23-25 years old who delivered at home, and had no ANC visit (Model 2)
**Figure 3.** Ratios associated with neonate, infant, child, 1-59 months, under-5 years, and stillbirth in SSA and South Asia for the 2014-2018 survey period. Reference group is mothers aged 23-25 years old who live in rural areas, have no formal education, and are in the poorer/middle/richer wealth quintile (Model 3).
**Figure 4.** Ratios associated with neonate, infant, child, 1-59 months, under-5 years, and stillbirth in SSA and South Asia for the 2014-2018 survey period. Reference group is mothers aged 23-25 years old who live in rural areas, have no formal education, are in the poorer/middle/richer wealth quintile, and had no ANC visit and at home delivery (Model 4).
**Figure 5.** Ratios associated with neonate, infant, child, 1-59 months, under-5 years, and stillbirth in SSA and South Asia for the 2009-2013 survey period. Reference group is mothers aged 23-25 years old (Model 0).

**Figure 6.** Ratios associated with neonate, infant, child, 1-59 months, under-5 years, and stillbirth in SSA and South Asia for the 2009-2013 survey period. Reference group is mothers aged 23-25 years old who live in rural areas and have no formal education (Model 1).
**Figure 7.** Ratios associated with neonate, infant, child, 1-59 months, under-5 years, and stillbirth in SSA and South Asia for the 2009-2013 survey period. Reference group is mothers aged 23-25 years old who delivered at home, and had no ANC visit (Model 2).
**Figure 8.** Ratios associated with neonate, infant, child, 1-59 months, under-5 years, and stillbirth in SSA for the 2009-2013 survey period. Reference group is mothers aged 23-25 years old who live in rural areas, have no formal education, and are in the poorer/middle/richer wealth quintile (Model 3).
**Figure 9.** Ratios associated with neonate, infant, child, 1-59 months, under-5 years, and stillbirth in SSA for the 2009-2013 survey period. Reference group is mothers aged 23-25 years old who live in rural areas, have no formal education, are in the poorer/middle/richer wealth quintile, and had no ANC visit and at home delivery (Model 4).
**Figure 10.** Ratios associated with neonate, infant, child, 1-59 months, under-5 years, and stillbirth in SSA and South Asia for the 2004-2008 survey period. Reference group is mothers aged 23-25 years old (Model 0).
**Figure 11.** Ratios associated with neonate, infant, child, 1-59 months, under-5 years, and stillbirth in SSA and South Asia for the 2004-2008 survey period. Reference group is mothers aged 23-25 years old who live in rural areas and have no formal education (Model 1).
**Figure 12.** Ratios associated with neonate, infant, child, 1-59 months, under-5 years, and stillbirth in SSA and South Asia for the 2004-2008 survey period. Reference group is mothers aged 23-25 years old who delivered at home and have no ANC visits (Model 2).
**Figure 13.** Ratios associated with neonate, infant, child, 1-59 months, under-5 years, and stillbirth in SSA and South Asia for the 2004-2008 survey period. Reference group is mothers aged 23-25 years old who live in rural areas, have no formal education, and are in the poorer/middle/richer wealth quintile, (Model 3).
**Figure 14.** Ratios associated with neonate, infant, child, 1-59 months, under-5 years, and stillbirth in SSA and South Asia for the 2004-2008 survey period. Reference group is mothers aged 23-25 years old who live in rural areas, have no formal education, are in the poorer/middle/richer wealth quintile, and had no ANC visit and at home delivery (Model 4).
**Figure 15.** Ratios associated with neonate, infant, child, 1-59 months, under-5 years, and stillbirth in SSA for the 2014-2018 survey period. Reference group is mothers aged 23-25 years old.
**Figure 16.** Ratios associated with neonate, infant, child, 1-59 months, under-5 years, and stillbirth in SSA for the 2014-2018 survey period. Reference group is mothers aged 23-25 years old who live in rural areas, and have no formal education.

### Appendix 1. List of countries in Sub-Saharan Africa (SSA) and South Asia with survey data

Angola, Benin, Burkina Faso, Burundi, Democratic Republic of Congo, Congo, Côte d'ivoire, Cameroon, Ethiopia, Gabon, Ghana, Gambia, Guinea, Kenya, Comoros, Liberia, Lesotho, Madagascar, Mali, Malawi, Mozambique, Nigeria, Niger, Namibia, Rwanda, Sierra Leone, Senegal, Sao Tome and Principe, Chad, Togo, Tanzania, Uganda, South Africa, Zambia, Zimbabwe, Afghanistan, Bangladesh, India, Indonesia, Cambodia, Myanmar, Maldives, Nepal, Philippines, Pakistan, Timor-Leste.

Table 1. Characteristics of the study population for the child health outcomes in SSA (Unweighted data). All numbers represent women's first birth out of their entire birth history, unless noted otherwise.

| | 2004-2008 | | | | | 2009-2013 | | | | | 2014-2018 | | | | |
|---|---|---|---|---|---|---|---|---|---|---|---|---|---|---|---|
| **Maternal age at first birth** | **<16** | **16-17** | **18-19** | **20-22** | **23-25** | **< 16** | **16-17** | **18-19** | **20-22** | **23-25** | **< 16** | **16-17** | **18-19** | **20-22** | **23-25** |
| Total births (%[a]) | 4404 (5.3) | 14639 (17.7) | 23258 (28.1) | 27688 (33.4) | 12942 (15.6) | 5356 (4.9) | 18569 (16.9) | 31204 (28.5) | 37062 (33.8) | 17451 (15.9) | 4157 (4.1) | 16429 (16.4) | 27787 (27.7) | 34526 (34.4) | 17544 (17.5) |
| Stillbirth (%[b]) | 22 (1.5) | 67 (0.8) | 119 (0.7) | 187 (0.5) | 245 (0.6) | 22 (1.2) | 98 (0.9) | 150 (0.6) | 275 (0.6) | 269 (0.5) | 26 (1.9) | 96 (1) | 175 (0.8) | 305 (0.7) | 307 (0.7) |
| Neonatal deaths (%[c]) | 361 (8.2) | 813 (5.6) | 1040 (4.5) | 1111 (4) | 513 (4) | 348 (6.5) | 925 (5) | 1187 (3.8) | 1302 (3.5) | 556 (3.2) | 212 (5.2) | 690 (4.3) | 939 (3.5) | 996 (3) | 483 (2.8) |
| Infant deaths (%) | 647 (14.7) | 1623 (11.1) | 1968 (8.5) | 2104 (7.6) | 935 (7.2) | 618 (11.5) | 1637 (8.8) | 2190 (7) | 2231 (6) | 928 (5.3) | 392 (9.6) | 1107 (6.9) | 1550 (5.7) | 1656 (4.9) | 758 (4.5) |
| Child deaths (%) | 316 (7.2) | 776 (5.3) | 888 (3.8) | 986 (3.6) | 418 (3.2) | 296 (5.5) | 694 (3.7) | 874 (2.8) | 943 (2.5) | 392 (2.2) | 190 (4.6) | 464 (2.9) | 575 (2.1) | 579 (1.7) | 242 (1.4) |
| 1-59 months deaths (%) | 603 (13.7) | 1588 (10.8) | 1820 (7.8) | 1983 (7.2) | 842 (6.5) | 570 (10.6) | 1407 (7.6) | 1877 (6) | 1873 (5.1) | 764 (4.4) | 370 (9) | 881 (5.5) | 1186 (4.4) | 1241 (3.7) | 517 (3) |
| Under-5 deaths (%) | 963 (21.9) | 2399 (16.4) | 2856 (12.3) | 3090 (11.2) | 1353 (10.5) | 914 (17.1) | 2331 (12.6) | 3064 (9.8) | 3174 (8.6) | 1320 (7.6) | 582 (14.2) | 1571 (9.8) | 2125 (7.8) | 2235 (6.6) | 1000 (5.9) |
| Residency | | | | | | | | | | | | | | | |
| Rural (%)[d] | 3407 (77.4) | 10623 (72.6) | 16249 (69.9) | 18320 (66.2) | 7594 (58.7) | 3941 (73.6) | 13299 (71.6) | 21969 (70.4) | 24444 (66) | 10131 (58.1) | 3052 (73.4) | 12030 (73.2) | 19655 (70.7) | 22688 (65.7) | 9813 (55.9) |
| Urban (%) | 997 (22.6) | 4016 (27.4) | 7009 (30.1) | 9368 (33.8) | 5348 (41.3) | 1415 (26.4) | 5270 (28.4) | 9235 (29.6) | 12618 (34) | 7320 (41.9) | 1105 (26.6) | 4399 (26.8) | 8132 (29.3) | 11838 (34.3) | 7731 (44.1) |
| Maternal Education | | | | | | | | | | | | | | | |
| No education (%) | 2672 (60.7) | 7174 (49) | 8907 (38.3) | 9223 (33.3) | 3977 (30.7) | 2709 (50.6) | 7581 (40.8) | 11129 (35.7) | 12489 (33.7) | 5640 (32.3) | 1867 (44.9) | 6255 (38.1) | 8710 (31.3) | 9896 (28.7) | 4734 (27) |
| Any level of education (%) | 1732 (39.3) | 7465 (51) | 14351 (61.7) | 18465 (66.7) | 8965 (69.3) | 2647 (49.4) | 10988 (59.2) | 20075 (64.3) | 24573 (66.3) | 11811 (67.7) | 2290 (55.1) | 101174 (61.9) | 19077 (68.7) | 24630 (71.3) | 12810 (73) |

| | 2004-2008 | | | | | 2009-2013 | | | | | 2014-2018 | | | | |
|---|---|---|---|---|---|---|---|---|---|---|---|---|---|---|---|
| **Maternal age at first birth** | **< 16** | **16-17** | **18-19** | **20-22** | **23-25** | **< 16** | **16-17** | **18-19** | **20-22** | **23-25** | **< 16** | **16-17** | **18-19** | **20-22** | **23-25** |
| Marital Status | | | | | | | | | | | | | | | |
| Never in union (%) | 327 (7.4) | 1378 (9.4) | 2253 (9.7) | 2479 (9) | 1089 (8.4) | 672 (12.5) | 2677 (14.4) | 3846 (12.3) | 3913 (10.6) | 1574 (9) | 549 (13.2) | 2187 (13.3) | 3434 (12.4) | 3845 (11.1) | 1731 (9.9) |
| Currently/Formerly in union/living with a man (%) | 4077 (92.6) | 13261 (90.6) | 21004 (90.3) | 25207 (91) | 11853 (91.6) | 4684 (87.5) | 15891 (85.6) | 27357 (87.7) | 33148 (89.4) | 15876 (91) | 3608 (86.8) | 14242 (86.7) | 24353 (87.6) | 30681 (88.9) | 15813 (90.1) |
| Place of Delivery [b] | | | | | | | | | | | | | | | |
| At home (%) | 797 (60.8) | 3734 (53.5) | 5372 (44.8) | 5383 (38) | 2160 (31.1) | 667 (40.8) | 3441 (37.7) | 5140 (31.5) | 4903 (25.7) | 1968 (20.8) | 562 (45.4) | 3002 (37.9) | 4200 (28.8) | 3788 (21.4) | 1510 (16.3) |
| Health facility (%) | 506 (38.6) | 3208 (46) | 6552 (54.6) | 8679 (61.3) | 4740 (68.2) | 955 (58.4) | 5625 (61.7) | 11058 (67.9) | 14045 (73.6) | 7459 (78.7) | 666 (53.8) | 4856 (61.4) | 10246 (70.3) | 13733 (77.8) | 7655 (82.7) |
| Other (%) | 8 (0.6) | 37 (0.5) | 69 (0.6) | 89 (0.6) | 53 (0.8) | 13 (0.8) | 51 (0.6) | 94 (0.6) | 131 (0.7) | 56 (0.6) | 9 (0.7) | 53 (0.7) | 125 (0.9) | 142 (0.8) | 90 (1) |
| Antenatal Care Visit [b] | | | | | | | | | | | | | | | |
| No visit (%) | 164 (24.4) | 857 (21.2) | 1149 (15.5) | 959 (11.1) | 366 (8.6) | 153 (17.1) | 638 (11.8) | 944 (9.1)9 | 807 (6.7) | 347 (5.9) | 139 (20.2) | 662 (14) | 880 (9.3) | 707 (6.2) | 323 (5.4) |
| 1+ visit (%) | 508 (75.6) | 3180 (78.8) | 6273 (84.5) | 7678 (88.9) | 3881 (91.4) | 740 (82.9) | 4785 (88.2) | 403 (90.9) | 11155 (93.3) | 5529 (94.1) | 548 (79.8) | 4079 (86) | 8586 (90.7) | 10682 (93.8) | 5697 (94.6) |

[a] % births in the dataset (age-group distribution).
[b] Stillbirths, place of delivery and antenatal care utilization data were available for births that took place 3 or 5 years before the survey.
[c] % of cases that ended in death (crude mortality rate).
[d] % of cases belonging to subgroup (within-cell distribution).

Table 2. Characteristics of the study population for the child health outcomes in South Asia (Unweighted data). All numbers represent women's first birth out of their entire birth history, unless noted otherwise.

| | **2004-2008** | | | | | **2009-2013** | | | | | **2014-2018** | | | | |
|---|---|---|---|---|---|---|---|---|---|---|---|---|---|---|---|
| **Maternal age at first birth** | **< 16** | **16-17** | **18-19** | **20-22** | **23-25** | **< 16** | **16-17** | **18-19** | **20-22** | **23-25** | **< 16** | **16-17** | **18-19** | **20-22** | **23-25** |
| Total births (%[a]) | 1835 (3.2) | 7084 (12.5) | 13405 (23.6) | 21767 (38.3) | 12737 (22.4) | 729 (2.2) | 3260 (9.7) | 7161 (21.3) | 13352 (39.7) | 9131 (27.1) | 2674 (1.4) | 13302 (6.7) | 38819 (19.7) | 86902 (44.1) | 55522 (28.2) |
| Stillbirth (%[b]) | 21 (3.3) | 75 (2.2) | 124 (1.4) | 207 (0.9) | 181 (0.8) | 5 (2.1) | 33 (2.1) | 37 (0.8) | 105 (0.9) | 93 (0.7) | 21 (3.1) | 78 (1.4) | 247 (1.1) | 601 (0.8) | 605 (0.7) |
| Neonatal deaths (%[c]) | 160 (8.7) | 451 (6.4) | 690 (5.1) | 786 (3.6) | 412 (3.2) | 41 (5.6) | 157 (4.8) | 295 (4.1) | 408 (3.1) | 228 (2.5) | 148 (5.5) | 643 (4.8) | 1552 (4) | 2771 (3.2) | 1514 (2.7) |
| Infant deaths (%) | 220 (12) | 647 (9.1) | 976 (7.3) | 1133 (5.2) | 585 (4.6) | 63 (8.6) | 229 (7) | 415 (5.8) | 575 (4.3) | 329 (3.6) | 219 (8.2) | 897 (6.7) | 2078 (5.4) | 3710 (4.3) | 1964 (3.5) |
| Child deaths (%) | 40 (2.2) | 101 (1.4) | 129 (1) | 164 (0.8) | 68 (0.5) | 8 (1.1) | 29 (0.9) | 55 (0.8) | 89 (0.7) | 43 (0.5) | 32 (1.2) | 108 (0.8) | 227 (0.6) | 437 (0.5) | 218 (0.4) |
| 1-59 months deaths (%) | 101 (5.5) | 299 (4.2) | 419 (3.1) | 514 (2.4) | 242 (1.9) | 30 (4.1) | 101 (3.1) | 175 (2.4) | 259 (1.9) | 144 (1.6) | 103 (3.9) | 362 (2.7) | 753 (1.9) | 1377 (1.6) | 670 (1.2) |
| Under-5 deaths (%) | 260 (14.2) | 748 (10.6) | 1105 (8.2) | 1297 (6) | 653 (5.1) | 71 (9.7) | 258 (7.9) | 470 (6.6) | 664 (5) | 372 (4.1) | 251 (9.4) | 1005 (7.6) | 2305 (5.9) | 4147 (4.8) | 2182 (3.9) |
| Residency | | | | | | | | | | | | | | | |
| Rural (%)[d] | 1335 (72.8) | 5088 (71.8) | 9139 (68.2) | 13336 (61.3) | 6716 (52.7) | 523 (71.7) | 2302 (70.6) | 4999 (69.8) | 8794 (65.9) | 2054 (76.8) | 10047 (75.5) | 29711 (76.5) | 64977 (74.8) | 38311 (69) | 2054 (76.8) |
| Urban (%) | 500 (27.2) | 1996 (28.2) | 4266 (31.8) | 8431 (38.7) | 6021 (47.3) | 206 (28.3) | 958 (29.4) | 2162 (30.2) | 4558 (34.1) | 620 (23.2) | 3255 (24.5) | 9108 (23.5) | 21925 (25.2) | 17211 (31) | 620 (23.2) |

| | 2004-2008 | | | | | 2009-2013 | | | | | 2014-2018 | | | | |
|---|---|---|---|---|---|---|---|---|---|---|---|---|---|---|---|
| **Maternal age at first birth** | **< 16** | **16-17** | **18-19** | **20-22** | **23-25** | **< 16** | **16-17** | **18-19** | **20-22** | **23-25** | **< 16** | **16-17** | **18-19** | **20-22** | **23-25** |
| Maternal Education | | | | | | | | | | | | | | | |
| No education (%) | 789 (43) | 2724 (38.5) | 4289 (32) | 5180 (23.8) | 2084 (16.4) | 161 (22.1) | 631 (19.4) | 1274 (17.8) | 1988 (14.9) | 1125 (12.3) | 933 (34.9) | 4287 (32.2) | 11204 (28.9) | 21739 (25) | 11502 (20.7) |
| Any level of education (%) | 1046 (57) | 4360 (61.5) | 9116 (68) | 16587 (76.2) | 10653 (83.6) | 568 (77.9) | 2629 (80.6) | 5887 (82.2) | 11364 (85.1) | 8006 (87.7) | 1741 (65.1) | 9015 (67.8) | 27615 (71.1) | 65163 (75) | 44020 (79.3) |
| Marital Status | | | | | | | | | | | | | | | |
| Never in union (%) | 1 (0.1) | 13 (0.2) | 29 (0.2) | 58 (0.3) | 35 (0.3) | 1 (0.1) | 19 (0.6) | 47 (0.7) | 76 (0.6) | 43 (0.5) | 12 (0.4) | 59 (0.4) | 126 (0.3) | 252 (0.3) | 167 (0.3) |
| Currently/Formerly in union/living with a man (%) | 1834 (99.9) | 7071 (99.8) | 13376 (99.8) | 21709 (99.7) | 12702 (99.7) | 728 (99.9) | 3241 (99.4) | 7114 (99.3) | 13276 (99.4) | 9088 (99.5) | 2662 (99.6) | 13243 (99.6) | 38693 (99.7) | 86650 (99.7) | 55355 (99.7) |
| Place of Delivery b | | | | | | | | | | | | | | | |
| At home (%) | 463 (79.1) | 2111 (74.1) | 4029 (64.1) | 5878 (52.3) | 2593 (38.5) | 155 (73.5) | 923 (66.1) | 1939 (55.2) | 2921 (42.4) | 1692 (33.5) | 225 (48.2) | 1533 (35.2) | 4406 (26) | 8332 (18.9) | 4350 (14.3) |
| Health facility (%) | 120 (20.5) | 731 (25.7) | 2232 (35.5) | 5328 (47.4) | 4136 (61.4) | 56 (26.5) | 471 (33.7) | 1564 (44.5) | 3931 (57.1) | 3333 (66.1) | 239 (51.2) | 2810 (64.4) | 12462 (73.6) | 35723 (80.9) | 25892 (85.3) |
| Other (%) | 2 (0.3) | 7 (0.2) | 20 (0.3) | 28 (0.2) | 7 (0.1) | 0 (0) | 3 (0.2) | 9 (0.3) | 32 (0.5) | 20 (0.4) | 3 (0.6) | 18 (0.4) | 60 (0.4) | 123 (0.3) | 114 (0.4) |
| Antenatal Care Visit b | | | | | | | | | | | | | | | |
| No visit (%) | 104 (32.2) | 462 (26.8) | 683 (17.5) | 806 (11) | 308 (6.7) | 42 (29) | 196 (20.1) | 304 (12.5) | 346 (7.1) | 171 (4.7) | 62 (22.5) | 452 (17.6) | 1613 (16.1) | 3235 (12.1) | 1830 (9.2) |
| 1+ visit (%) | 219 (67.8) | 1264 (73.2) | 3216 (82.5) | 6537 (89) | 4307 (93.3) | 103 (71) | 780 (79.9) | 2129 (87.5) | 4504 (92.9) | 3431 (95.3) | 213 (77.5) | 2120 (82.4) | 8434 (83.9) | 23503 (87.9) | 18135 (90.8) |

[a] % births in the dataset (age-group distribution).
[b] Stillbirths, place of delivery and antenatal care utilization data and were available for births taking place 3 or 5 years before the survey."
[c] % of cases that ended in death (crude mortality rate).
[d] % of cases belonging to subgroup (within-cell distribution).

Table 3. Child and 1-59 months mortality rates and their sampling errors in SSA

| | 2004-2008 | | 2009-2013 | | 2014-2018 | |
|---|---|---|---|---|---|---|
| | **CMR**[a] | **1-59MR**[b] | **CMR** | **1-59MR** | **CMR** | **1-59MR** |
| Maternal age at first birth (years) | | | | | | |
| < 16 | 101.2±5.2 | 153.4±6.0 | 77.6±4.4 | 122.5±5.2 | 67 ± 4.5 | 101.9 ± 5.3 |
| 16-17 | 82.0±2.9 | 130±3.4 | 59.3±2.2 | 95.4±2.6 | 43.4 ± 2 | 68.4 ± 2.5 |
| 18-19 | 62.6±2.1 | 99.1±2.4 | 45.0±1.6 | 76.4±1.9 | 32 ± 1.4 | 53.8 ± 1.8 |
| 20-22 | 52.2±1.8 | 87.4±2.2 | 38.6±1.3 | 62.8±1.6 | 25.5 ± 1.1 | 44.8 ± 1.4 |
| 23-25 | 52.9±2.6 | 83.3±3.2 | 35.1±1.8 | 55.2±2.2 | 21.2 ± 1.5 | 36.7 ± 1.8 |
| Urban/Rural Residency | | | | | | |
| < 16 | 75.8 ± 7.6<br>107.7 ± 3.4 | 129.4 ± 8<br>159.3 ± 4.4 | 57.1±2.7<br>85.1±3.8 | 100.8±5.9<br>130.3±3.5 | 45.1 ± 3.2<br>75 ± 0.6 | 72.7 ± 8.8<br>112.3 ± 4.7 |
| 16-17 | 69.6 ± 1<br>86 ± 8.2 | 110.3 ± 1.6<br>136.1 ± 7.4 | 38.9±1.3<br>68.1±2.4 | 69.9±1.4<br>105.8±2.4 | 30.1 ± 0.9<br>48.7 ± 0.9 | 51.6 ± 6.2<br>74.9 ± 1.3 |
| 18-19 | 47.7 ± 1<br>68.3 ± 2 | 80 ± 1.7<br>106.2 ± 2.4 | 29.7±2.6<br>52.4±2.7 | 54.3±2.6<br>86.8±3.7 | 23.3 ± 1<br>35.9 ± 1.5 | 41.6 ± 2.9<br>59.2 ± 1.4 |
| 20-22 | 36.8 ± 2.1<br>59.1 ± 1.9 | 65.2 ± 1.7<br>97.2 ± 2.2 | 24.6±1.4<br>46.6±3.6 | 42.2±1.4<br>74.3±4.6 | 17.3 ± 5.1<br>30.3 ± 0.4 | 32.4 ± 4.6<br>51.8 ± 0.6 |
| 23-25 | 28 ± 0.3<br>67.7 ± 3.9 | 53 ± 2<br>101 ± 3.8 | 25.7±0.9<br>42.3±3.3 | 40.0±1.2<br>67.0±3.2 | 15.5 ± 0.3<br>26.1 ± 0.2 | 27.8 ± 0.5<br>44.5 ± 0.4 |
| Maternal Education Status: Any Education/No Education | | | | | | |
| < 16 | 69.3 ± 0.8<br>121.5 ± 5.1 | 115.8 ± 1.1<br>176.3 ± 6.7 | 64.4 ± 1.3<br>87.6 ± 4.7 | 110.3 ± 2.5<br>131.5 ± 4.9 | 52.3 ± 2.5<br>83.6 ± 1.1 | 85 ± 6<br>121.1 ± 6.7 |
| 16-17 | 57.2 ± 0.5<br>106.1 ± 10.9 | 98.5 ± 0.8<br>159.8 ± 9.2 | 47.9 ± 0.5<br>72.7 ± 3.6 | 79.5 ± 0.8<br>114.4 ± 3.4 | 32.7 ± 0.5<br>59.8 ± 0.9 | 57 ± 3.8<br>85.7 ± 1.1 |
| 18-19 | 46.9 ± 0.5<br>86.6 ± 2.1 | 80.1 ± 0.9<br>127.4 ± 3.2 | 34.9 ± 1.9<br>60.9 ± 4.2 | 63.5 ± 3.8<br>97 ± 5.2 | 25.4 ± 0.8<br>45.5 ± 3 | 44.9 ± 1.3<br>72.4 ± 2.8 |
| 20-22 | 39 ± 0.2<br>78 ± 3.5 | 69 ± 0.4<br>122.2 ± 3.7 | 29.8 ± 2<br>53.8 ± 5.7 | 51.8 ± 1.9<br>82 ± 7.7 | 21 ± 0.5<br>36 ± 0.3 | 37.3 ± 0.9<br>62.8 ± 0.6 |
| 23-25 | 39.6 ± 0.3<br>81.5 ± 6.6 | 67 ± 0.6<br>117.7 ± 6.4 | 28.3 ± 0.2<br>47.5 ± 5.4 | 46 ± 0.3<br>72.5 ± 5.3 | 16.4 ± 0.3<br>33.8 ± 0.1 | 30.6 ± 0.5<br>53.2 ± 0.2 |
| Antenatal Care Visit (ANC): Any Visit/No Visit | | | | | | |
| < 16 | 116.9 ± 13.8<br>93 ± 14.7 | 153.9 ± 13.9<br>102.7 ± 14.4 | 51.7 ± 24.4<br>99.2 ± 48.1 | 82.6 ± 26.1<br>127.2 ± 43.8 | - | - |
| 16-17 | 36.2 ± 11.6<br>87.8 ± 5.3 | 67.2 ± 11<br>115.7 ± 11.4 | 43.7 ± 1.1<br>33.7 ± 1.7 | 68.3 ± 1.5<br>67.1 ± 3 | 26.1 ± 0.4<br>67.2 ± 3.3 | 40.6 ± 0.7<br>100 ± 4 |
| 18-19 | 33.9 ± 1.2<br>60 ± 0.7 | 55.1 ± 1.7<br>91.8 ± 2 | 42.1 ± 0.4<br>123.4 ± 8.3 | 64.2 ± 3.6<br>139.6 ± 10.6 | 21 ± 0.6<br>43.8 ± 1.7 | 33 ± 0.9<br>65.6 ± 2.2 |
| 20-22 | 30.2 ± 0.5<br>117.2 ± 17.8 | 54.4 ± 0.8<br>153.9 ± 17.4 | 19.5 ± 0.2<br>81.6 ± 9.8 | 37.2 ± 0.5<br>93.9 ± 13.7 | 11.1 ± 0.2<br>47.2 ± 2.2 | 22.3 ± 0.3<br>82.6 ± 2.9 |
| 23-25 | 32 ± 0.7<br>93.1 ± 259.9 | 56.6 ± 1.3<br>107.5 ± 248.8 | 29.8 ± 0.2<br>19 ± 1.3 | 44.1 ± 0.3<br>42.5 ± 2.6 | 9.4 ± 0.6<br>46.2 ± 6.1 | 17.7 ± 0.8<br>80.8 ± 8.8 |

| | 2004-2008 | | 2009-2013 | | 2014-2018 | |
|---|---|---|---|---|---|---|
| | **CMR** | **1-59MR** | **CMR** | **1-59MR** | **CMR** | **1-59MR** |
| Place of Delivery: At Home/Health Facility | | | | | | |
| < 16 | 109.8 ± 6.1<br>104.5 ± 28.2 | 165.1 ± 6.9<br>129 ± 28.4 | 110.2 ± 8.5<br>69.8 ± 0.6 | 148.7 ± 9.2<br>99.6 ± 9.3 | 122.6 ± 1.5<br>66.6 ± 3.9 | 162.3 ± 5.1<br>98.9 ± 11.3 |
| 16-17 | 88.4 ± 3.6<br>54.3 ± 8.9 | 133.6 ± 4.2<br>96.5 ± 8.2 | 72.3 ± 3.6<br>50.1 ± 1.6 | 99.7 ± 4<br>81.8 ± 2.1 | 76.3 ± 1.8<br>33.4 ± 0.3 | 99.7 ± 2.1<br>56.9 ± 0.9 |
| 18-19 | 61.8 ± 6.2<br>38.3 ± 1.2 | 102.1 ± 5.7<br>65.8 ± 1.9 | 57.7 ± 2.1<br>36.8 ± 0.9 | 90.6 ± 2.9<br>62.2 ± 1.2 | 48 ± 0.4<br>22.4 ± 0.9 | 75.9 ± 1.5<br>37.3 ± 1.1 |
| 20-22 | 56.6 ± 1.6<br>38 ± 0.7 | 98.3 ± 2.2<br>67.5 ± 1.2 | 42.6 ± 2.1<br>27.8 ± 0.8 | 66.2 ± 2.8<br>48.5 ± 1.2 | 36.9 ± 1<br>15.2 ± 0.1 | 66.6 ± 1.7<br>30.3 ± 0.2 |
| 23-25 | 62.2 ± 2.4<br>39.4 ± 0.8 | 94.7 ± 5.4<br>65.5 ± 1.2 | 38.6 ± 23.7<br>28.7 ± 0 | 66.3 ± 21.8<br>44.5 ± 0.1 | 27.5 ± 1.4<br>12.8 ± 0.4 | 49.1 ± 2.1<br>24.1 ± 0.6 |

[a] CMR: Child mortality rate, [b] 1-59MR: 1-59 months mortality rate

Table 4. Child and 1-59 months mortality rates and their sampling errors in South Asia

| | 2004-2008 | | 2009-2013 | | 2014-2018 | |
|---|---|---|---|---|---|---|
| | **CMR** | **1-59MR** | **CMR** | **1-59MR** | **CMR** | **1-59MR** |
| Maternal age at first birth (years) | | | | | | |
| < 16 | 36.5 ± 0.6 | 62.1 ± 1 | 18.8 ± 6.8 | 43.6 ± 6.2 | 12.4 ± 2.6 | 36 ± 7.8 |
| 16-17 | 19.2 ± 0.3 | 46.6 ± 2.6 | 12.8 ± 0.3 | 36 ± 2.6 | 10.3 ± 0.7 | 27.6 ± 2 |
| 18-19 | 13.1 ± 0.2 | 33.3 ± 0.9 | 10 ± 2 | 25.3 ± 1.9 | 7.8 ± 5.5 | 20 ± 5.1 |
| 20-22 | 11.1 ± 0.1 | 27.3 ± 0.2 | 8.3 ± 0.1 | 21.1 ± 0.2 | 6.6 ± 0.2 | 16.9 ± 0.5 |
| 23-25 | 8.3 ± 0.1 | 22 ± 0.2 | 9 ± 0.04 | 19.8 ± 0.3 | 6.6 ± 0.2 | 14.4 ± 1.7 |
| Urban/Rural Residency | | | | | | |
| < 16 | 27.7 ± 1.2<br>38.3 ± 0.6 | 51.8 ± 2.3<br>64.2 ± 1 | 24.5 ± 1.1<br>17 ± 7.3 | 37.7 ± 10.8<br>45.3 ± 6.7 | 5.6 ± 2.4<br>14.5 ± 0.7 | 20.7 ± 10.6<br>40.6 ± 3.2 |
| 16-17 | 16.9 ± 5.8<br>19.8 ± 0.3 | 36 ± 4.8<br>49.1 ± 2.5 | 7 ± 1.2<br>14.4 ± 0.5 | 26.1 ± 4.3<br>38.8 ± 2.6 | 10.6 ± 1.5<br>10.2 ± 7.9 | 27.2 ± 4.1<br>27.7 ± 7.5 |
| 18-19 | 8.1 ± 0.1<br>14.8 ± 0.2 | 24.5 ± 0.2<br>36.2 ± 1 | 4.8 ± 1.7<br>11.7 ± 2.5 | 21.6 ± 2.2<br>26.5 ± 2.3 | 5.5 ± 11<br>8.6 ± 0.3 | 15.4 ± 10.4<br>21.6 ± 0.9 |
| 20-22 | 5.8 ± 0.1<br>13.6 ± 0.1 | 17 ± 0.2<br>32 ± 0.3 | 5.1 ± 0.7<br>10 ± 0.1 | 13.8 ± 0.6<br>24.7 ± 0.2 | 4.1 ± 0.2<br>7.7 ± 0.1 | 12 ± 0.7<br>19 ± 1.1 |
| 23-25 | 6.5 ± 0<br>9.7 ± 0.1 | 16.4 ± 0.1<br>26 ± 0.2 | 7.1 ± 0.1<br>10.4 ± 0.1 | 16.4 ± 0.5<br>22.1 ± 0.1 | 5 ± 0.2<br>7.5 ± 0.2 | 10.3 ± 2.5<br>16.8 ± 0.3 |
| Maternal Education Status: Any Education/No Education | | | | | | |
| < 16 | 27.7 ± 0.7<br>44.8 ± 0.5 | 52.6 ± 1.3<br>70.8 ± 0.4 | 21.2 ± 7.8<br>11.9 ± 0.4 | 39.6 ± 7.2<br>59.3 ± 2.3 | 11.2 ± 2.3<br>14.2 ± 3 | 28.8 ± 6.4<br>49.4 ± 8.6 |
| 16-17 | 12.4 ± 0.2<br>26.9 ± 0.3 | 35.7 ± 0.7<br>59.4 ± 5.4 | 7.4 ± 0.2<br>31.9 ± 0.6 | 27.7 ± 3<br>66 ± 1 | 8 ± 0.5<br>14.7 ± 1.5 | 22.9 ± 1.6<br>37.5 ± 4.1 |
| 18-19 | 9.7 ± 0.2<br>18.3 ± 0.2 | 26.4 ± 1.3<br>44.3 ± 0.4 | 9.5 ± 2.4<br>12.3 ± 0.1 | 21.7 ± 2.2<br>41 ± 0.4 | 6.2 ± 7.9<br>11.4 ± 0.9 | 16.2 ± 7.4<br>29.7 ± 2.6 |
| 20-22 | 7.6 ± 0.1<br>20.1 ± 3.4 | 21.1 ± 0.2<br>43 ± 3.1 | 7.3 ± 0.1<br>13.8 ± 0.1 | 19.1 ± 0.2<br>32 ± 0.2 | 5 ± 0.1<br>11.1 ± 0.5 | 13.4 ± 0.4<br>27.5 ± 1.2 |
| 23-25 | 6.7 ± 0.1<br>14.8 ± 0 | 16.8 ± 0.1<br>43.4 ± 0.2 | 7.7 ± 0<br>17.8 ± 0.3 | 16.8 ± 0.4<br>39.4 ± 0.5 | 5.2 ± 0.1<br>11 ± 0.8 | 11 ± 0.1<br>26.7 ± 6.2 |
| Antenatal Care Visit (ANC): Any Visit/No Visit | | | | | | |
| < 16 | 11.4 ± 0.5<br>2.2 ± 0.2 | 43.7 ± 1.6<br>3.5 ± 0.3 | - | - | 27.3 ± 6.1<br>0 ± 0 | 26.5 ± 5.8<br>0 ± 0 |

| | 2004-2008 | | 2009-2013 | | 2014-2018 | |
|---|---|---|---|---|---|---|
| | **CMR** | **1-59MMR** | **CMR** | **1-59MR** | **CMR** | **1-59MR** |
| 16-17 | 17.8 ± 0.7<br>5 ± 0.2 | 29.6 ± 0.9<br>40.7 ± | 3.4 ± 7.2<br>7.4 ± 0.7 | 7.8 ± 7<br>12.1 ± 4.9 | 2.6 ± 0.3<br>17.1 ± 2 | 9.4 ± 1.1<br>43.2 ± 4.1 |
| 18-19 | 4.9 ± 0.1<br>3.2 ± 0.1 | 13.5 ± 0.2<br>9.1 ± 8 | 2.9 ± 0.1<br>8.6 ± 0.5 | 7.1 ± 0.2<br>27.1 ± 1.1 | 2.4 ± 0.2<br>5.8 ± 0.7 | 9.5 ± 0.7<br>26.4 ± 2.1 |
| 20-22 | 5 ± 0<br>24 ± 1 | 13 ± 0.1<br>36.9 ± 1.3 | 1.5 ± 0<br>1.5 ± 5.6 | 4.6 ± 0.1<br>15.2 ± 5.4 | 3 ± 0.1<br>8.5 ± 0.5 | 9.2 ± 0.4<br>20.1 ± 2.4 |
| 23-25 | 0.6 ± 0<br>7.9 ± 0.8 | 6.5 ± 0.1<br>32.4 ± 2 | 6.5 ± 0.1<br>0 ± 0 | 13 ± 0.7<br>5.7 ± 0.7 | 3.7 ± 0<br>9.2 ± 0.3 | 8.3 ± 0.2<br>34.2 ± 1.1 |
| Place of Delivery: At Home/Health Facility | | | | | | |
| < 16 | 36.5 ± 2<br>0 ± 0 | 63.4 ± 2.9<br>35.2 ± 2.7 | 15.9 ± 37.9<br>0 ± 0 | 22.2 ± 36.4<br>24.9 ± 2.6 | 36.1 ± 4.4<br>71.1 ± 6.8 | 58.8 ± 11.6<br>85.3 ± 6.9 |
| 16-17 | 13.3 ± 0.2<br>11.7 ± 0.3 | 42.8 ± 5.1<br>32.4 ± 0.9 | 8.7 ± 0.1<br>10.2 ± 0.4 | 28 ± 0.4<br>9.8 ± 0.4 | 8.5 ± 0.4<br>11.1 ± 2.2 | 34.3 ± 2.3<br>25.7 ± 3.2 |
| 18-19 | 20.6 ± 0.2<br>7.2 ± 0.1 | 38.6 ± 3.5<br>19.6 ± 0.3 | 10.8 ± 7.2<br>5.5 ± 0.2 | 20.2 ± 6.7<br>13.7 ± 0.5 | 6.1 ± 0.2<br>5.8 ± 0.7 | 26.7 ± 1.4<br>17.3 ± 1.7 |
| 20-22 | 18.1 ± 4<br>5 ± 0.1 | 34.7 ± 3.6<br>13 ± 0.1 | 8.5 ± 0.2<br>5.9 ± 0 | 19.2 ± 0.3<br>12 ± 0.1 | 19.4 ± 4.2<br>6.8 ± 0.5 | 33.1 ± 5.4<br>16.5 ± 1.1 |
| 23-25 | 9.1 ± 0.1<br>5.3 ± 0.1 | 29.7 ± 0.3<br>12.5 ± 0.2 | 10.8 ± 0.2<br>6.2 ± 0 | 24.6 ± 0.5<br>14 ± 1.5 | 16.1 ± 0.8<br>7.1 ± 0.3 | 30.8 ± 9.4<br>13.8 ± 0.6 |

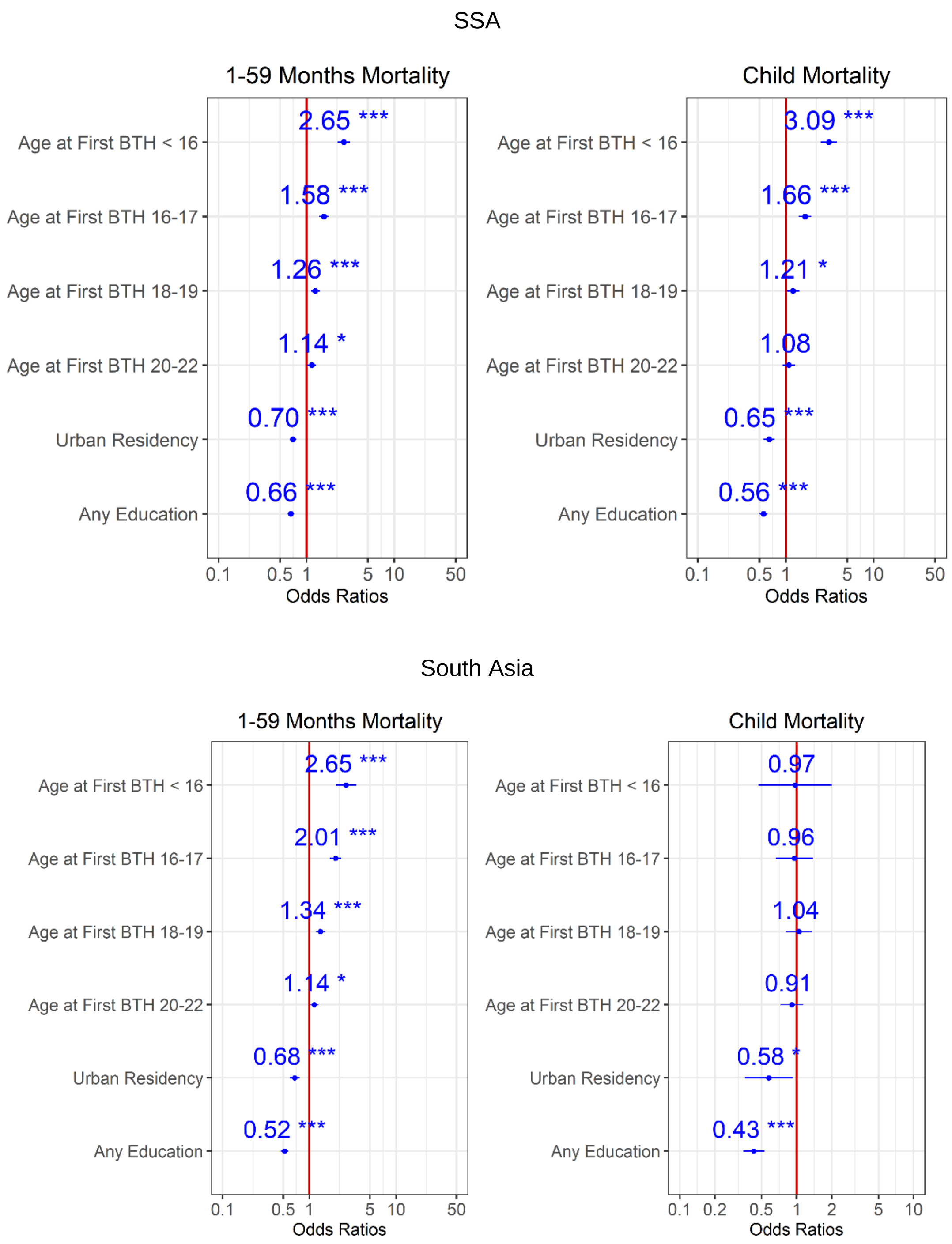


Figure 1. Ratios associated with 1-59 months and child mortality in SSA and South Asia for the 2014-2018 survey period. Risk factors reducing the probability of death have odds ratios lower than 1 to the left of the vertical red line. Odds ratios (blue points), 95% confidence intervals (horizontal blue lines) are given. P-values are shown with the asterisk signs (‘***’ 0.001 ‘**’ 0.01 ‘*’ 0.05 ‘.’ 0.1 ‘ ’ 1). Reference group is mothers aged 23-25 years old who live in rural areas and have no formal education (Model 1).

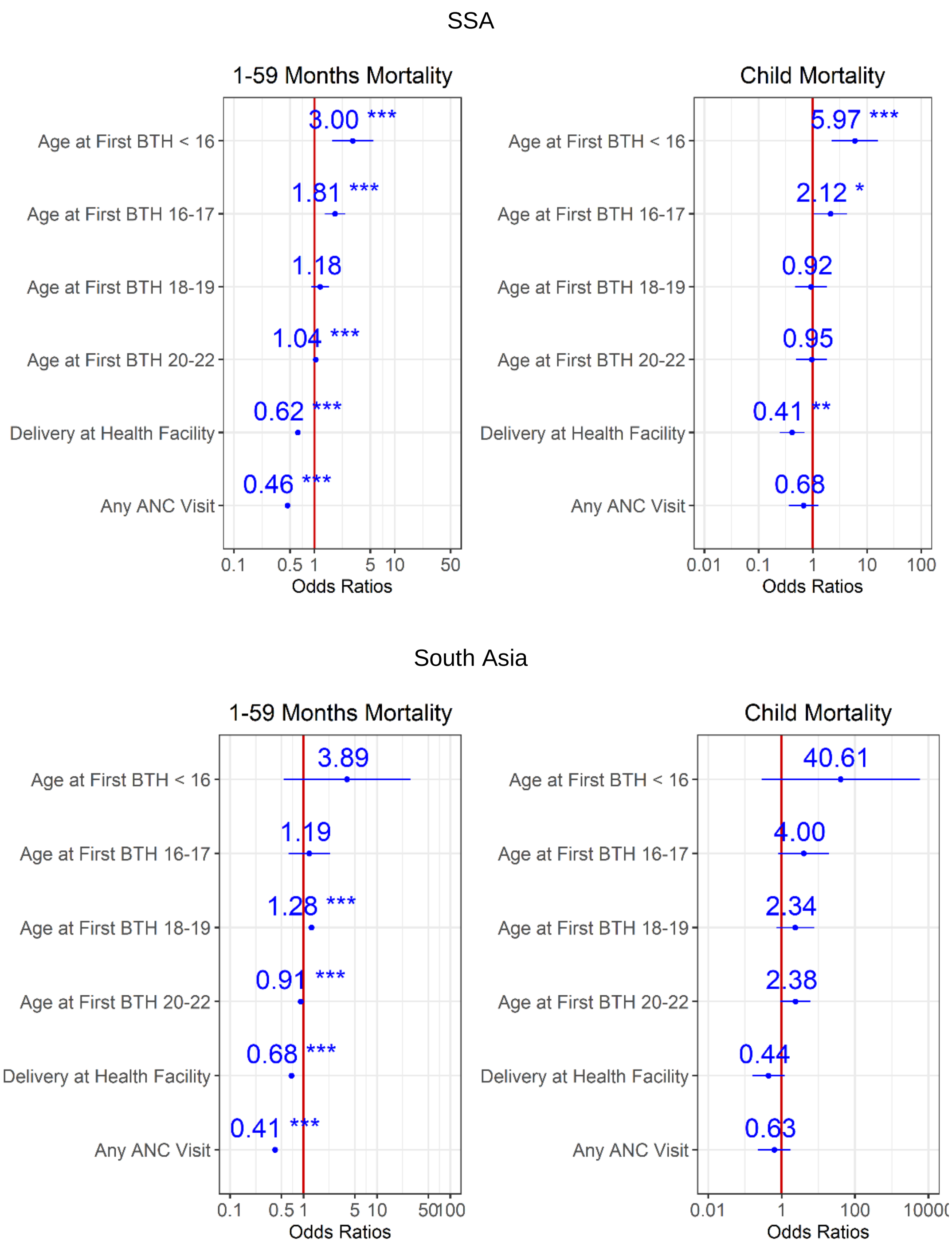


Figure 2. Ratios associated with 1-59 months and child mortality in SSA and South Asia for the 2014-2018 survey period. Risk factors reducing the probability of death have odds ratios lower than 1 to the left of the vertical red line. Odds ratios (blue points), 95% confidence intervals (horizontal blue lines) are given. P-values are shown with the asterisk signs (‘***’ 0.001 ‘**’ 0.01 ‘*’ 0.05 ‘.’ 0.1 ‘ ’ 1). Reference group is mothers aged 23-25 years old who delivered at home, and had no ANC visit (Model 2).

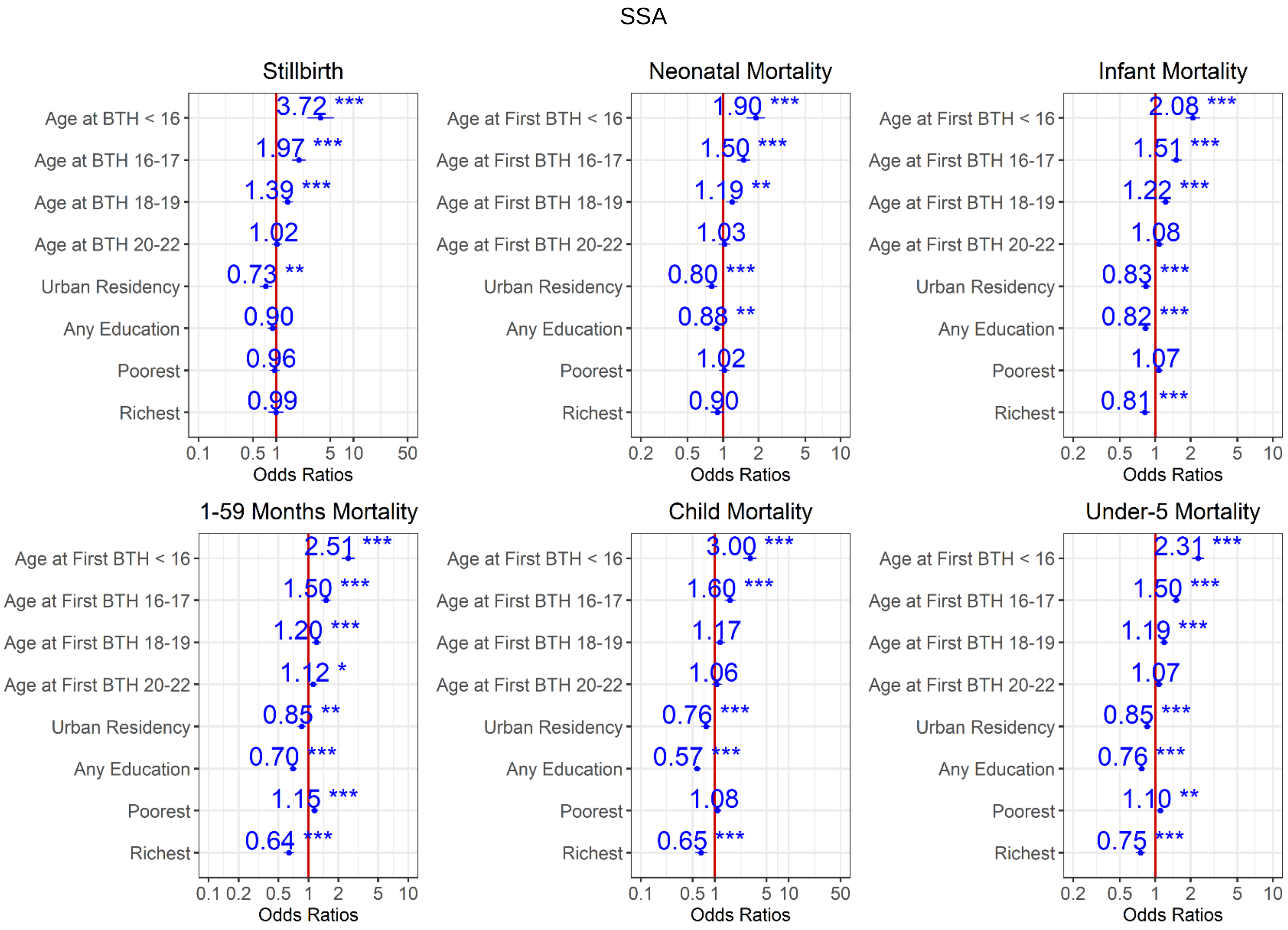
SSA
Stillbirth
Age at BTH < 16
Age at BTH 16-17
Age at BTH 18-19
Age at BTH 20-22
Urban Residency
Any Education
Poorest
Richest
3.72 ***
1.97 ***
1.39 ***
1.02
0.73 **
0.90
0.96
0.99
0.1 0.5 1 5 10 50
Odds Ratios
Neonatal Mortality
Age at First BTH < 16
Age at First BTH 16-17
Age at First BTH 18-19
Age at First BTH 20-22
Urban Residency
Any Education
Poorest
Richest
1.90 ***
1.50 ***
1.19 **
1.03
0.80 ***
0.88 **
1.02
0.90
0.2 0.5 1 2 5 10
Odds Ratios
Infant Mortality
Age at First BTH < 16
Age at First BTH 16-17
Age at First BTH 18-19
Age at First BTH 20-22
Urban Residency
Any Education
Poorest
Richest
2.08 ***
1.51 ***
1.22 ***
1.08
0.83 ***
0.82 ***
1.07
0.81 ***
0.2 0.5 1 2 5 10
Odds Ratios
1-59 Months Mortality
Age at First BTH < 16
Age at First BTH 16-17
Age at First BTH 18-19
Age at First BTH 20-22
Urban Residency
Any Education
Poorest
Richest
2.51 ***
1.50 ***
1.20 ***
1.12 *
0.85 **
0.70 ***
1.15 ***
0.64 ***
0.1 0.2 0.5 1 2 5 10
Odds Ratios
Child Mortality
Age at First BTH < 16
Age at First BTH 16-17
Age at First BTH 18-19
Age at First BTH 20-22
Urban Residency
Any Education
Poorest
Richest
3.00 ***
1.60 ***
1.17
1.06
0.76 ***
0.57 ***
1.08
0.65 ***
0.1 0.5 1 5 10 50
Odds Ratios
Under-5 Mortality
Age at First BTH < 16
Age at First BTH 16-17
Age at First BTH 18-19
Age at First BTH 20-22
Urban Residency
Any Education
Poorest
Richest
2.31 ***
1.50 ***
1.19 ***
1.07
0.85 ***
0.76 ***
1.10 **
0.75 ***
0.2 0.5 1 2 5 10
Odds Ratios

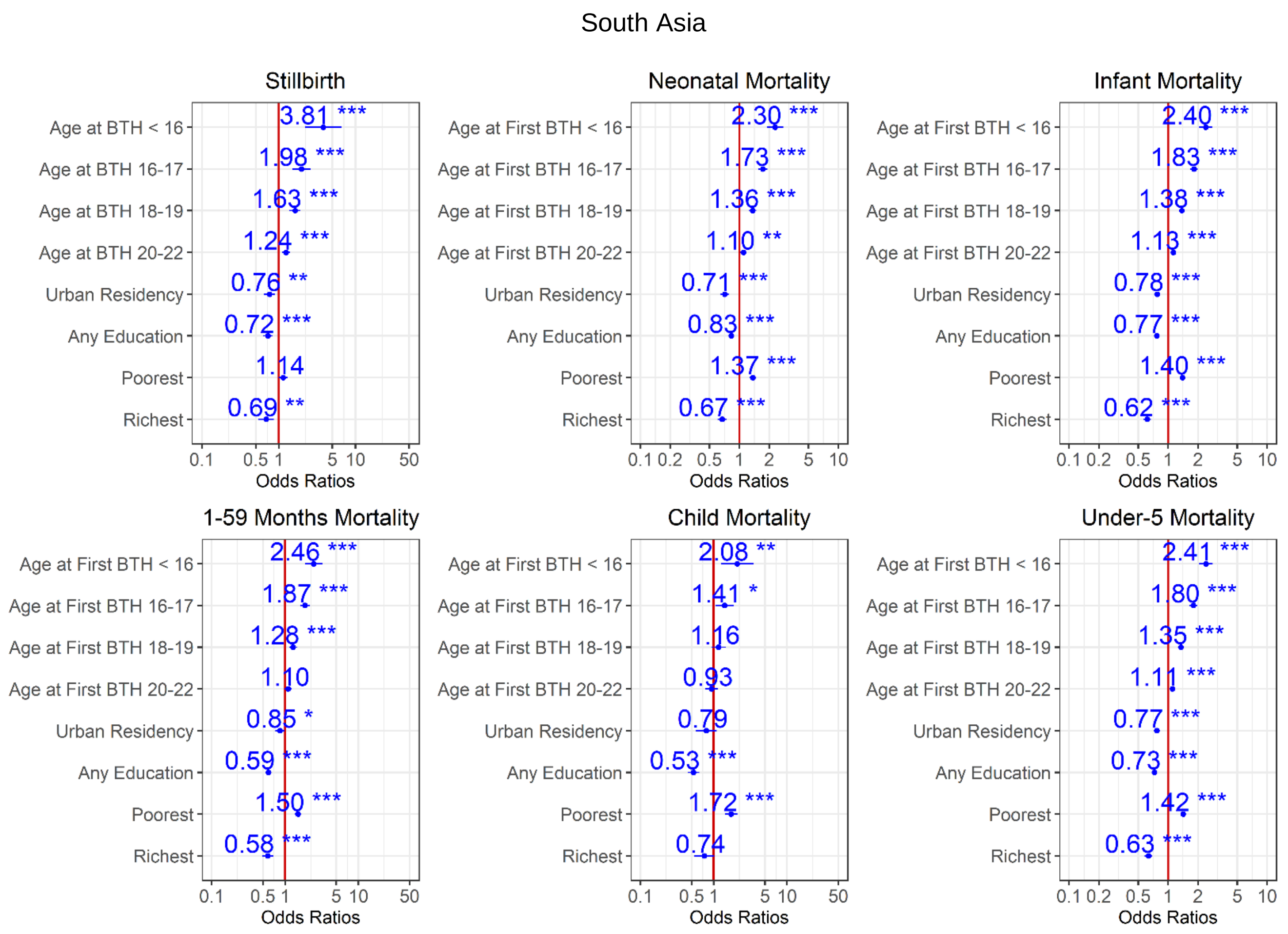


Figure 3. Ratios associated with neonate, infant, child, 1-59 months, under-5 years, and stillbirth in SSA and South Asia for the 2014-2018 survey period. Risk factors reducing the probability of death have odds ratios lower than 1 to the left of the vertical red line. Odds ratios (blue points), 95% confidence intervals (horizontal blue lines) are given. P-values are shown with the asterisk signs (‘***’ 0.001 ‘**’ 0.01 ‘*’ 0.05 ‘.’ 0.1 ‘ ’ 1). Reference group is mothers aged 23-25 years old who live in rural areas, have no formal education, and are in the poorer/middle/richer wealth quintile (Model 3).

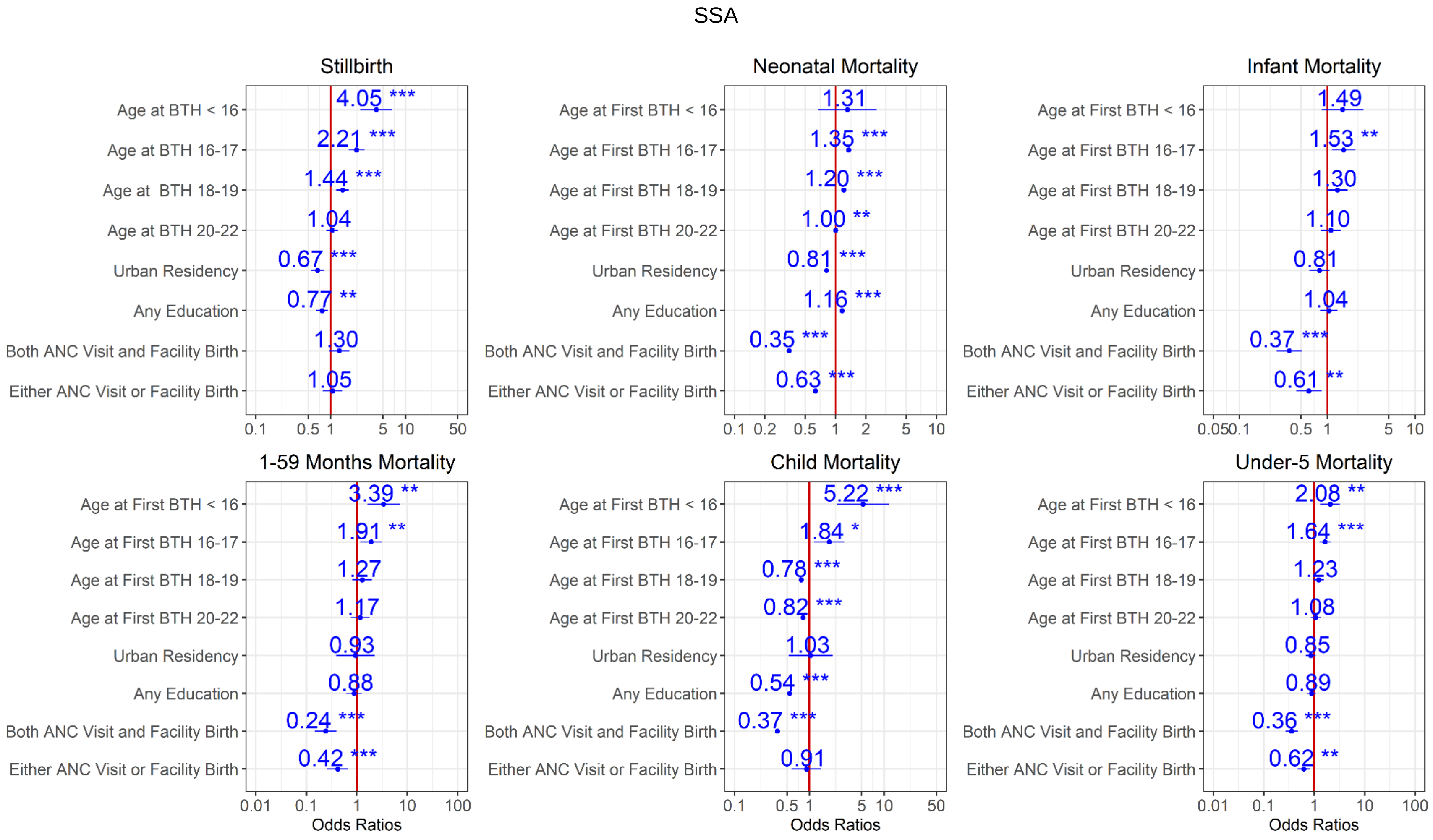
SSA
Stillbirth
Neonatal Mortality
Infant Mortality
1-59 Months Mortality
Child Mortality
Under-5 Mortality
Age at BTH < 16
Age at BTH 16-17
Age at BTH 18-19
Age at BTH 20-22
Age at First BTH < 16
Age at First BTH 16-17
Age at First BTH 18-19
Age at First BTH 20-22
Urban Residency
Any Education
Both ANC Visit and Facility Birth
Either ANC Visit or Facility Birth
4.05 ***
2.21 ***
1.44 ***
1.04
0.67 ***
0.77 **
1.30
1.05
1.31
1.35 ***
1.20 ***
1.00 **
0.81 ***
1.16 ***
0.35 ***
0.63 ***
1.49
1.53 **
1.30
1.10
0.81
1.04
0.37 ***
0.61 **
3.39 **
1.91 **
1.27
1.17
0.93
0.88
0.24 ***
0.42 ***
5.22 ***
1.84 *
0.78 ***
0.82 ***
1.03
0.54 ***
0.37 ***
0.91
2.08 **
1.64 ***
1.23
1.08
0.85
0.89
0.36 ***
0.62 **
Odds Ratios

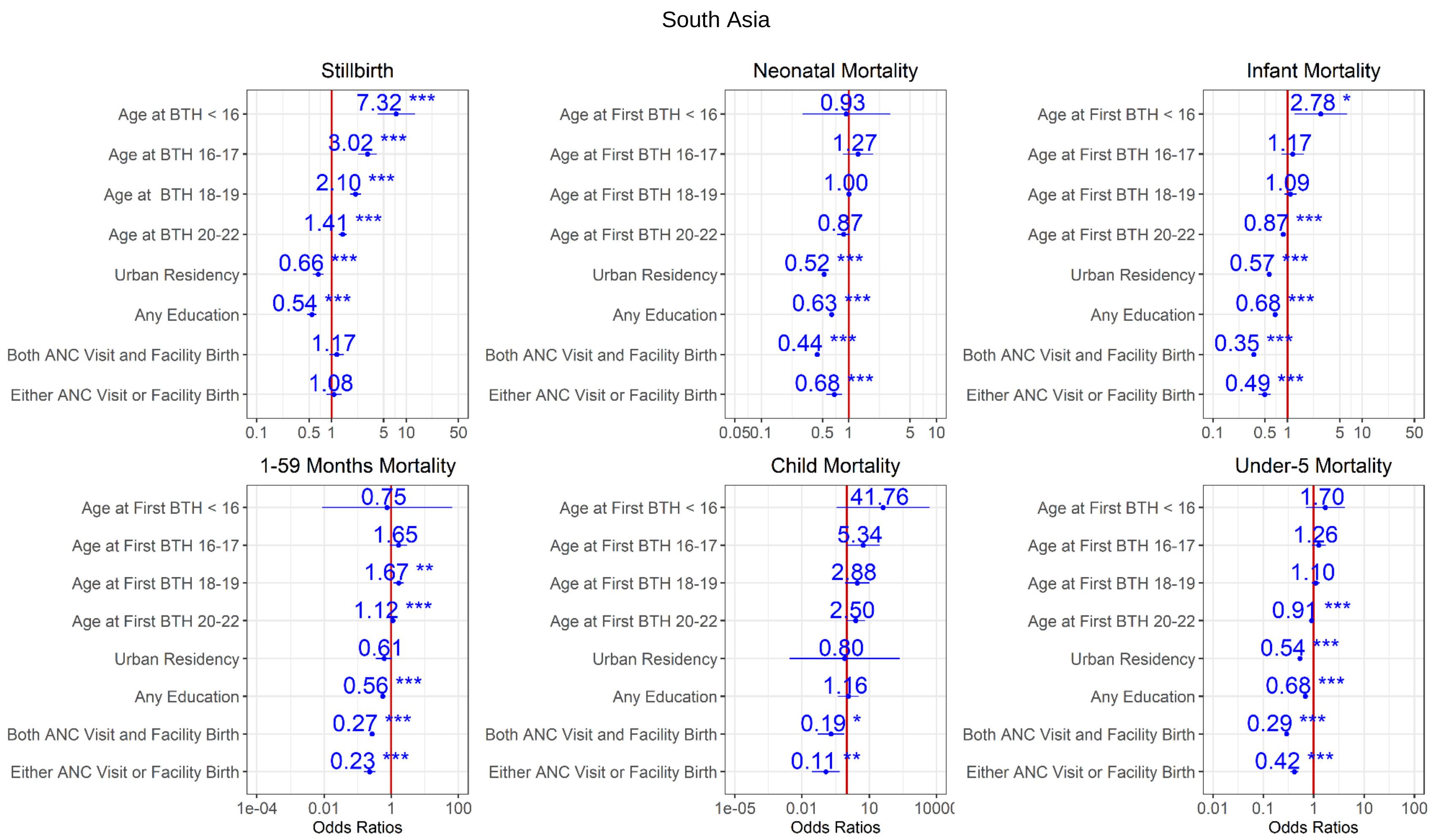


Figure 4. Ratios associated with neonate, infant, child, 1-59 months, under-5 years, and stillbirth in SSA and South Asia for the 2014-2018 survey period. Risk factors reducing the probability of death have odds ratios lower than 1 to the left of the vertical red line. Odds ratios (blue points), 95% confidence intervals (horizontal blue lines) are given. P-values are shown with asterisk signs (‘***’ 0.001 ‘**’ 0.01 ‘*’ 0.05 ‘.’ 0.1 ‘ ’ 1). Reference group is mothers aged 23-25 years old who live in rural areas, have no formal education, are in the poorer/middle/richer wealth quintile, and had no ANC visit and at home delivery (Model 4).

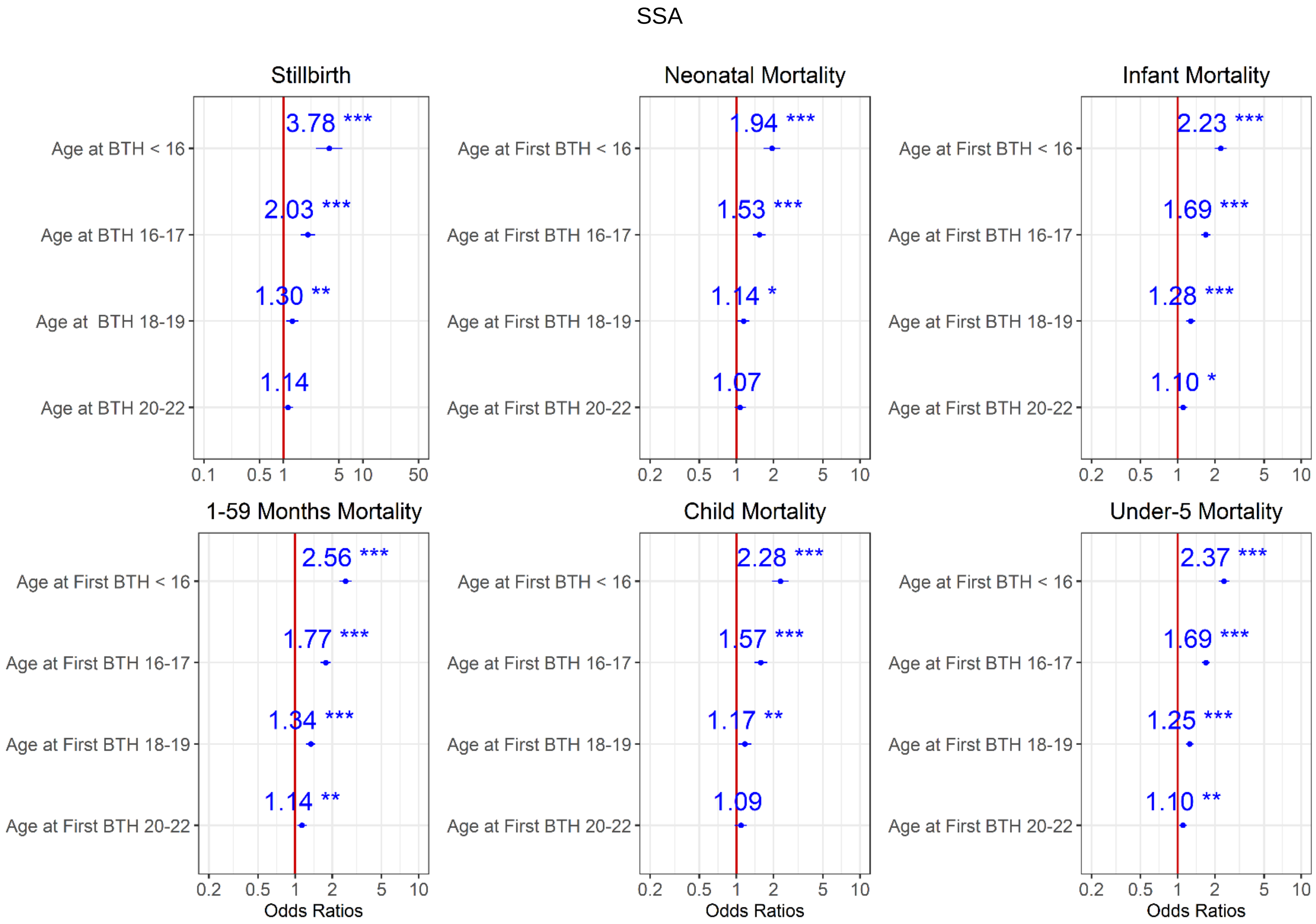
SSA
Stillbirth
Age at BTH < 16
3.78 ***
Age at BTH 16-17
2.03 ***
Age at BTH 18-19
1.30 **
Age at BTH 20-22
1.14
0.1 0.5 1 5 10 50
Neonatal Mortality
Age at First BTH < 16
1.94 ***
Age at First BTH 16-17
1.53 ***
Age at First BTH 18-19
1.14 *
Age at First BTH 20-22
1.07
0.2 0.5 1 2 5 10
Infant Mortality
Age at First BTH < 16
2.23 ***
Age at First BTH 16-17
1.69 ***
Age at First BTH 18-19
1.28 ***
Age at First BTH 20-22
1.10 *
0.2 0.5 1 2 5 10
1-59 Months Mortality
Age at First BTH < 16
2.56 ***
Age at First BTH 16-17
1.77 ***
Age at First BTH 18-19
1.34 ***
Age at First BTH 20-22
1.14 **
0.2 0.5 1 2 5 10
Odds Ratios
Child Mortality
Age at First BTH < 16
2.28 ***
Age at First BTH 16-17
1.57 ***
Age at First BTH 18-19
1.17 **
Age at First BTH 20-22
1.09
0.2 0.5 1 2 5 10
Odds Ratios
Under-5 Mortality
Age at First BTH < 16
2.37 ***
Age at First BTH 16-17
1.69 ***
Age at First BTH 18-19
1.25 ***
Age at First BTH 20-22
1.10 **
0.2 0.5 1 2 5 10
Odds Ratios

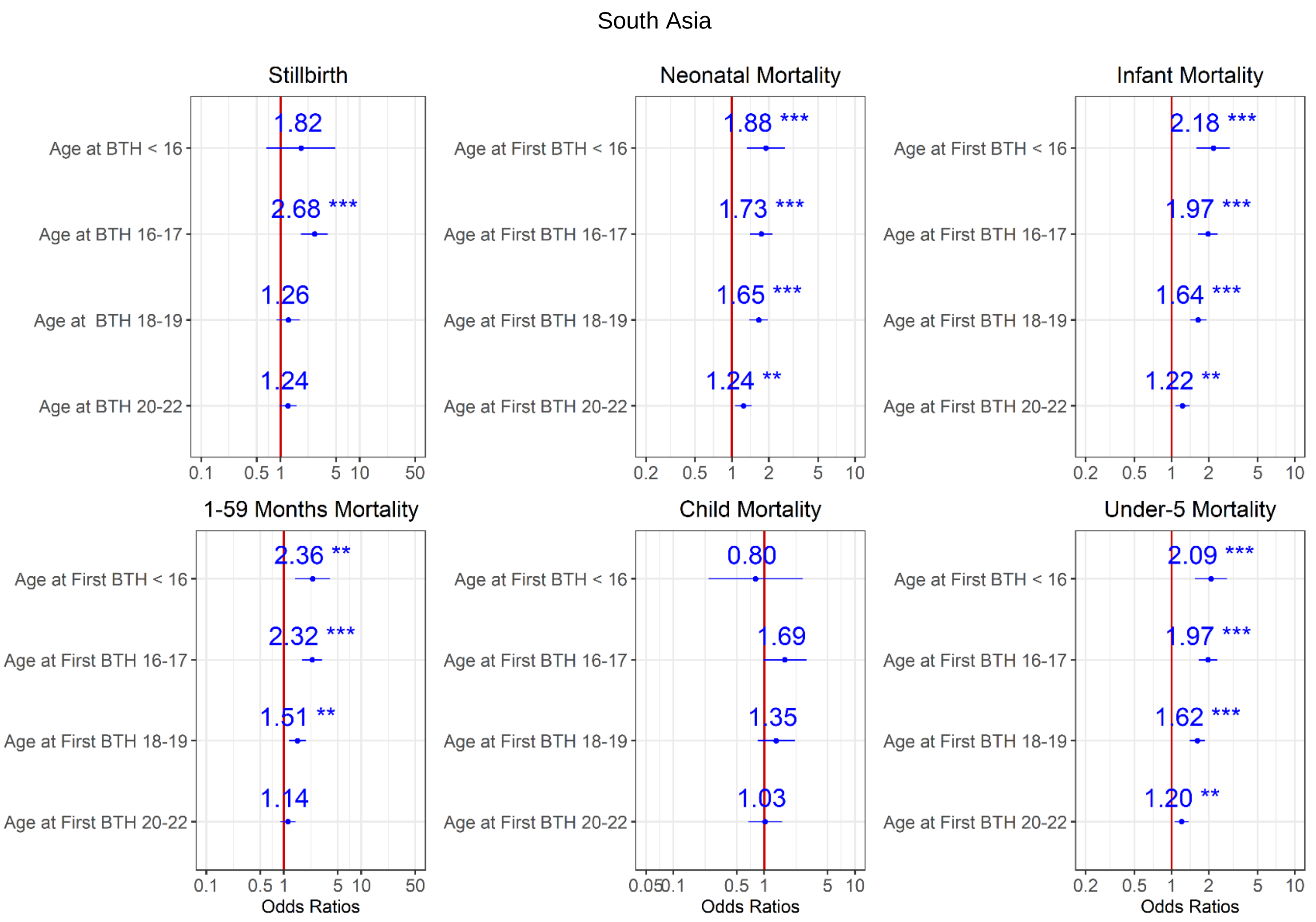


Figure 5. Ratios associated with neonate, infant, child, 1-59 months, under-5 years, and stillbirth in SSA and South Asia for the 2009-2013 survey period. Risk factors reducing the probability of death have odds ratios lower than 1 to the left of the vertical red line. Odds ratios (blue points), 95% confidence intervals (horizontal blue lines) are given. P-values are shown with the asterisk signs (‘***’ 0.001 ‘**’ 0.01 ‘*’ 0.05 ‘.’ 0.1 ‘ ’ 1). Reference group is mothers aged 23-25 years old (Model 0).

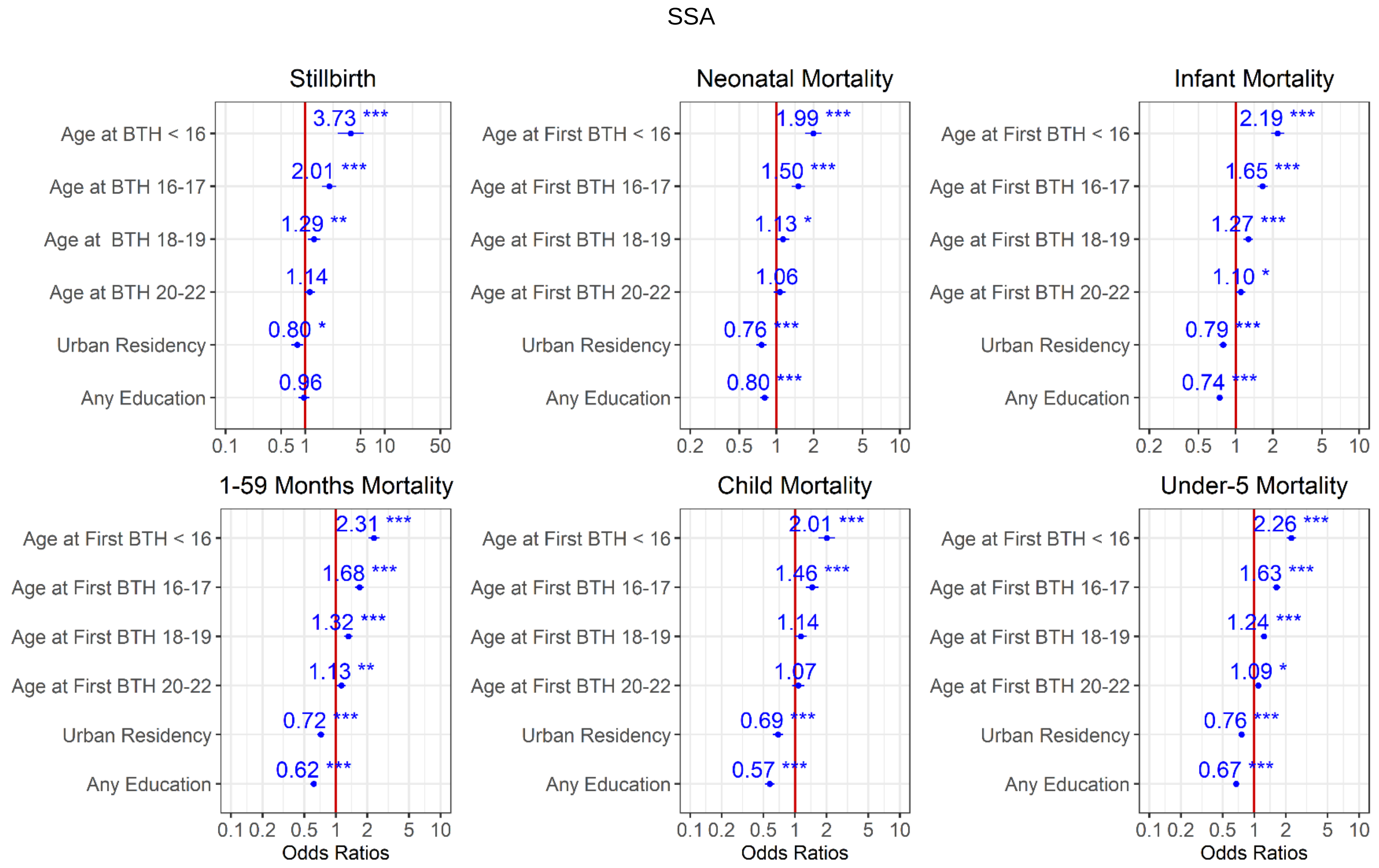
SSA
Stillbirth
Age at BTH < 16
Age at BTH 16-17
Age at  BTH 18-19
Age at BTH 20-22
Urban Residency
Any Education
3.73 ***
2.01 ***
1.29 **
1.14
0.80 *
0.96
0.1 0.5 1 5 10 50
Neonatal Mortality
Age at First BTH < 16
Age at First BTH 16-17
Age at First BTH 18-19
Age at First BTH 20-22
Urban Residency
Any Education
1.99 ***
1.50 ***
1.13 *
1.06
0.76 ***
0.80 ***
0.2 0.5 1 2 5 10
Infant Mortality
2.19 ***
1.65 ***
1.27 ***
1.10 *
0.79 ***
0.74 ***
0.2 0.5 1 2 5 10
1-59 Months Mortality
2.31 ***
1.68 ***
1.32 ***
1.13 **
0.72 ***
0.62 ***
0.1 0.2 0.5 1 2 5 10
Odds Ratios
Child Mortality
2.01 ***
1.46 ***
1.14
1.07
0.69 ***
0.57 ***
Under-5 Mortality
2.26 ***
1.63 ***
1.24 ***
1.09 *
0.76 ***
0.67 ***

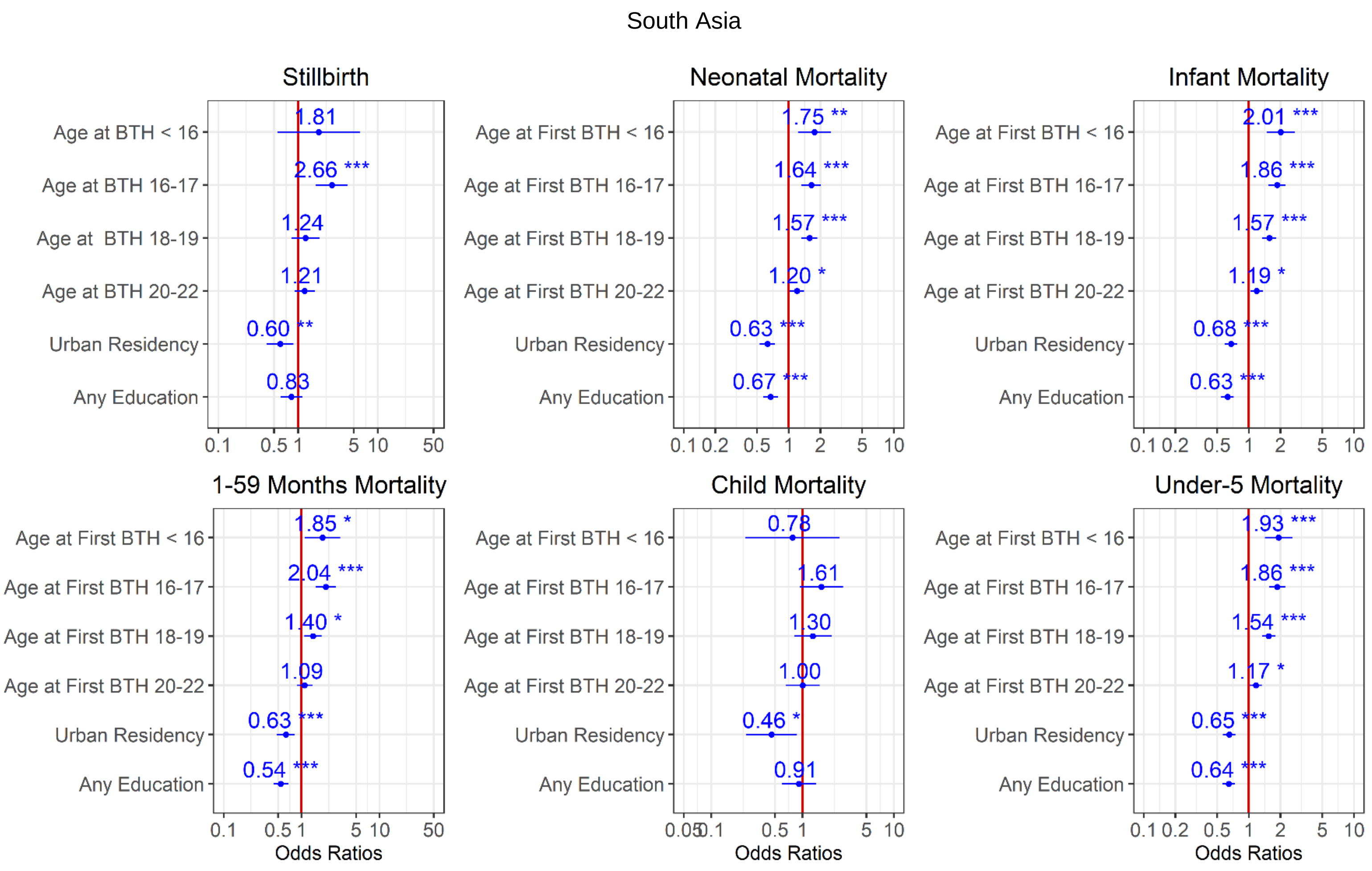


Figure 6. Ratios associated with neonate, infant, child, 1-59 months, under-5 years, and stillbirth in SSA and South Asia for the 2009-2013 survey period. Risk factors reducing the probability of death have odds ratios lower than 1 to the left of the vertical red line. Odds ratios (blue points), 95% confidence intervals (horizontal blue lines) are given. P-values are shown with the asterisk signs ('***' 0.001 '**' 0.01 '*' 0.05 '.' 0.1 ' ' 1). Reference group is mothers aged 23-25 years old who live in rural areas and have no formal education (Model 1).

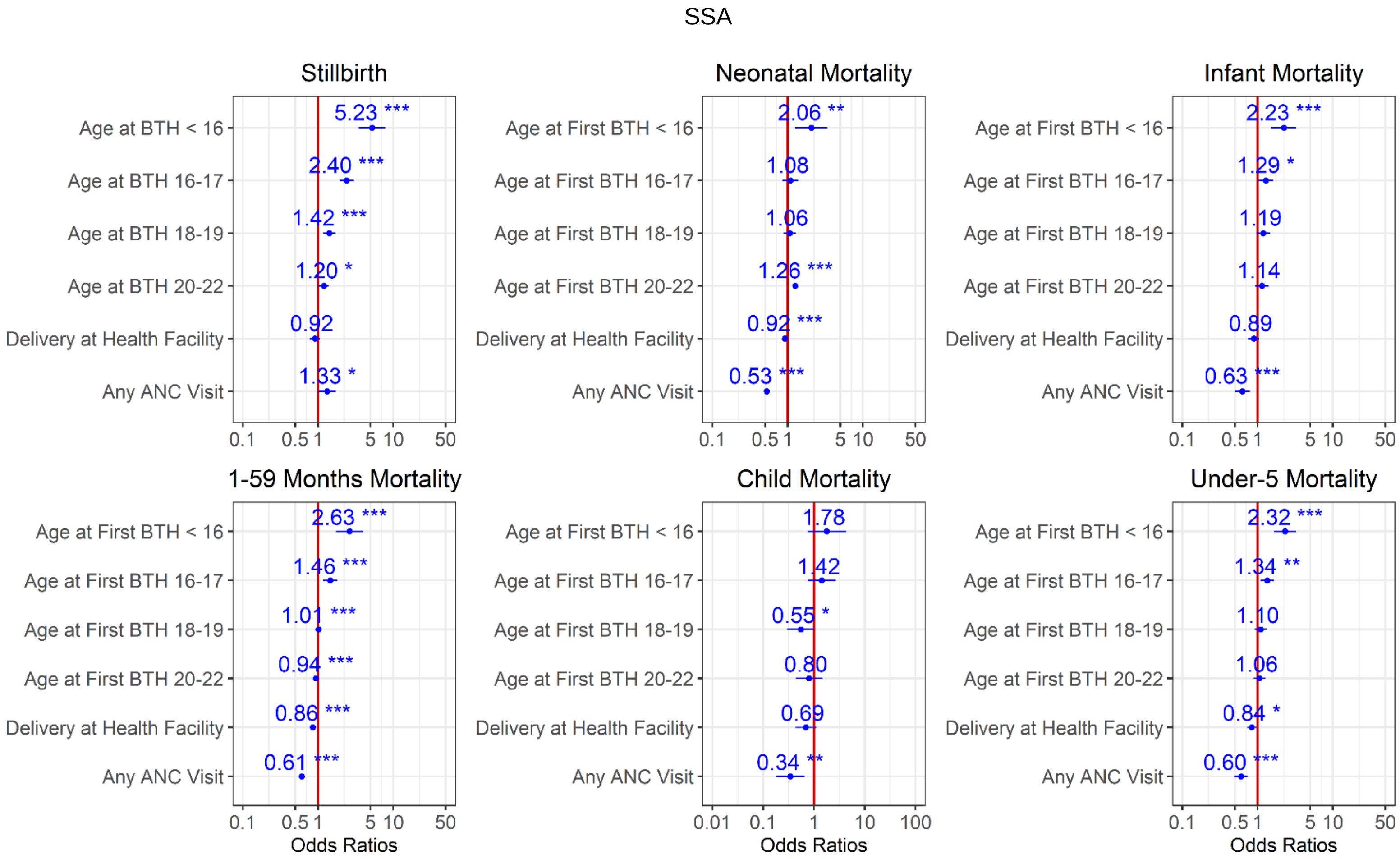


Figure 7. Ratios associated with neonate, infant, child, 1-59 months, under-5 years, and stillbirth in SSA and South Asia for the 2009-2013 survey period. Risk factors reducing the probability of death have odds ratios lower than 1 to the left of the vertical red line. Odds ratios (blue points), 95% confidence intervals (horizontal blue lines) are given. P-values are shown with the asterisk signs ('***' 0.001 '**' 0.01 '*' 0.05 '.' 0.1 ' ' 1). Reference group is mothers aged 23-25 years old who delivered at home, and had no ANC visit (Model 2).

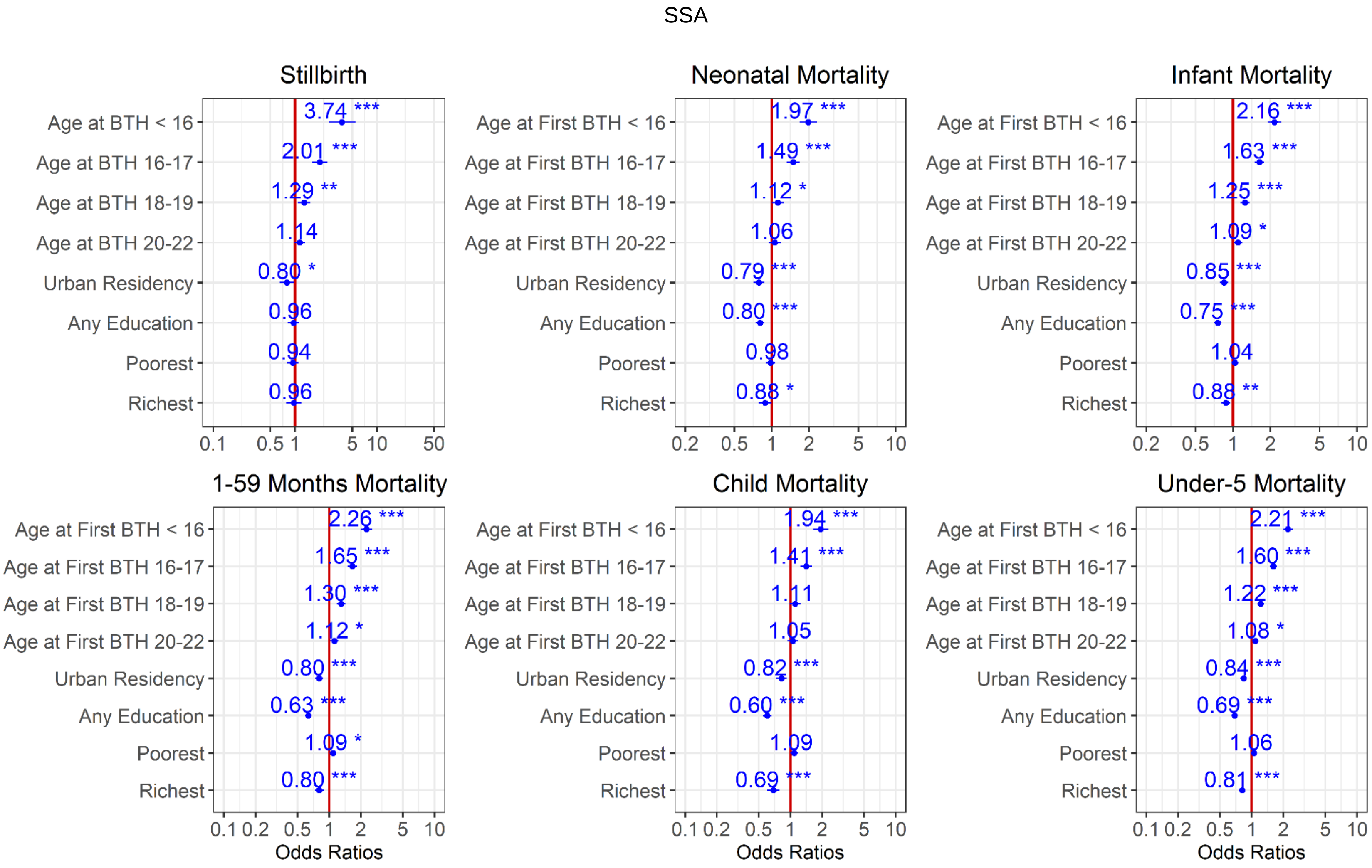
SSA
Stillbirth
Age at BTH < 16
Age at BTH 16-17
Age at BTH 18-19
Age at BTH 20-22
Urban Residency
Any Education
Poorest
Richest
3.74 ***
2.01 ***
1.29 **
1.14
0.80 *
0.96
0.94
0.96
0.1 0.5 1 5 10 50
Neonatal Mortality
Age at First BTH < 16
Age at First BTH 16-17
Age at First BTH 18-19
Age at First BTH 20-22
1.97 ***
1.49 ***
1.12 *
1.06
0.79 ***
0.80 ***
0.98
0.88 *
0.2 0.5 1 2 5 10
Infant Mortality
2.16 ***
1.63 ***
1.25 ***
1.09 *
0.85 ***
0.75 ***
1.04
0.88 **
1-59 Months Mortality
2.26 ***
1.65 ***
1.30 ***
1.12 *
0.80 ***
0.63 ***
1.09 *
0.80 ***
0.1 0.2 0.5 1 2 5 10
Odds Ratios
Child Mortality
1.94 ***
1.41 ***
1.11
1.05
0.82 ***
0.60 ***
1.09
0.69 ***
Under-5 Mortality
2.21 ***
1.60 ***
1.22 ***
1.08 *
0.84 ***
0.69 ***
1.06
0.81 ***

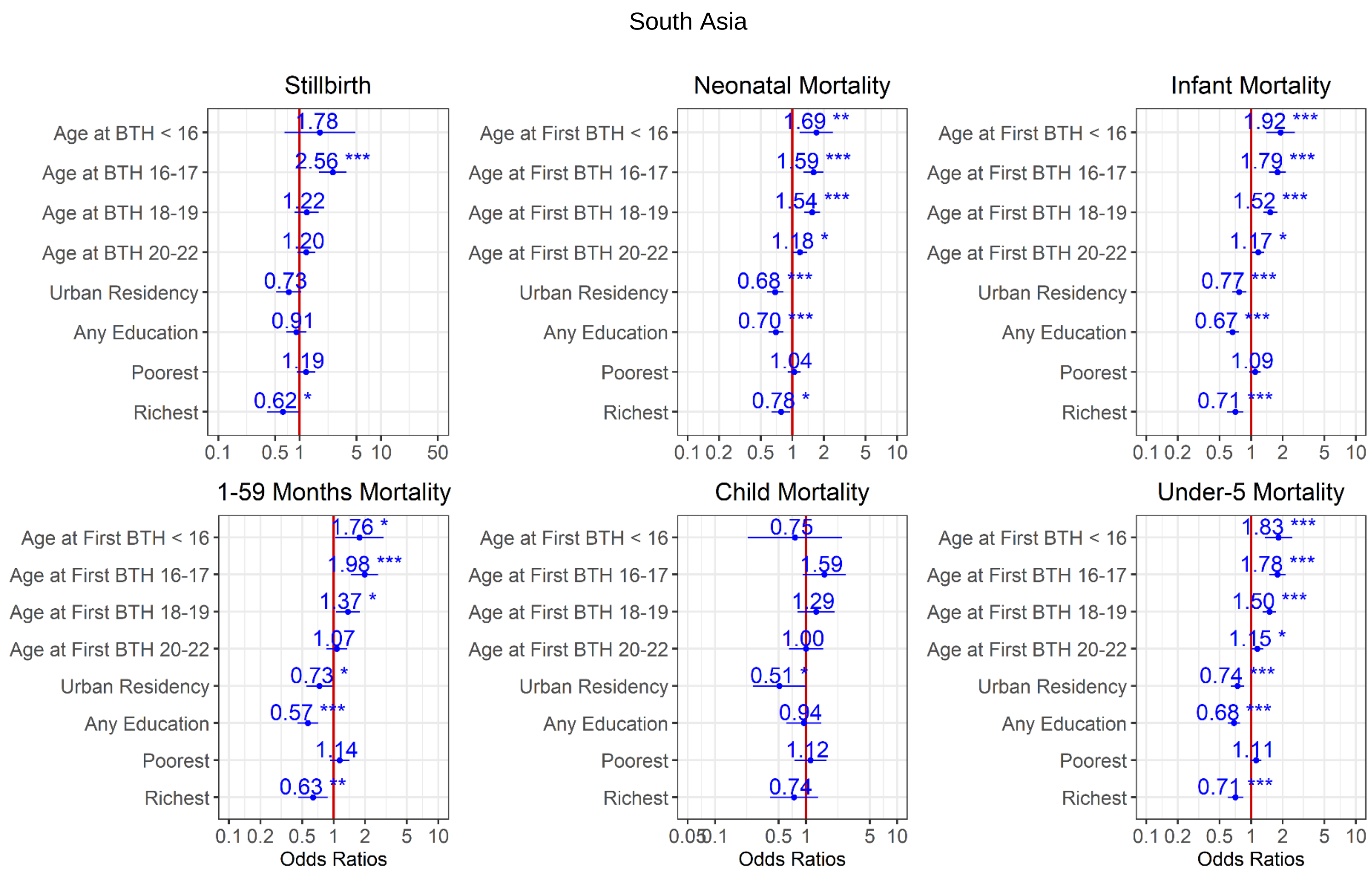


Figure 8. Ratios associated with neonate, infant, child, 1-59 months, under-5 years, and stillbirth in SSA for the 2009-2013 survey period. Risk factors reducing the probability of death have odds ratios lower than 1 to the left of the vertical red line. Odds ratios (blue points), 95% confidence intervals (horizontal blue lines) are given. P-values are shown with asterisk signs ('***' 0.001 '**' 0.01 '*' 0.05 '.' 0.1 ' ' 1). Reference group is mothers aged 23-25 years old who live in rural areas, have no formal education, and are in the poorer/middle/richer wealth quintile (Model 3).

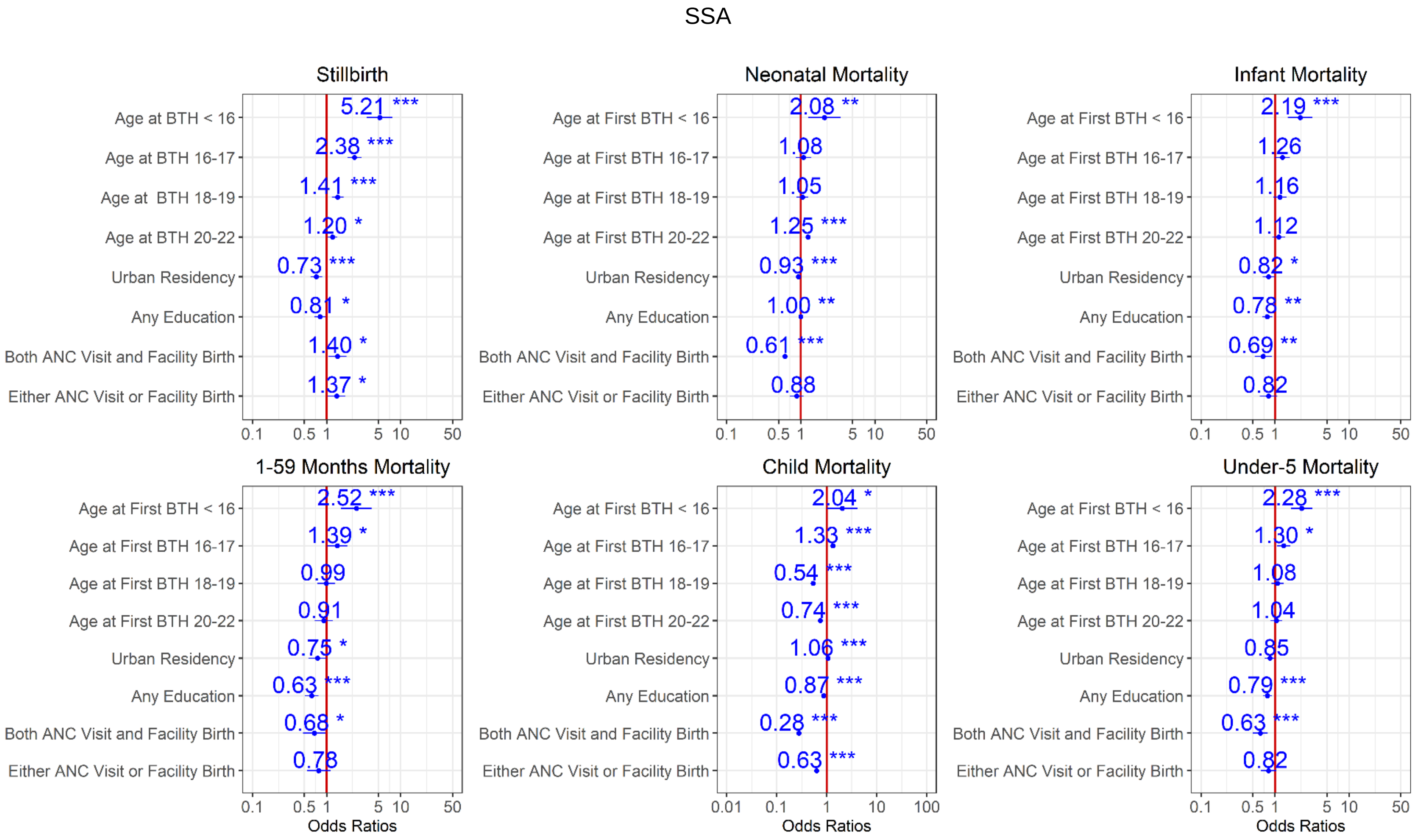


Figure 9. Ratios associated with neonate, infant, child, 1-59 months, under-5 years, and stillbirth in SSA for the 2009-2013 survey period. Risk factors reducing the probability of death have odds ratios lower than 1 to the left of the vertical red line. Odds ratios (blue points), 95% confidence intervals (horizontal blue lines) are given. P-values are shown with the asterisk signs (‘***’ 0.001 ‘**’ 0.01 ‘*’ 0.05 ‘.’ 0.1 ‘ ’ 1). Reference group is mothers aged 23-25 years old who live in rural areas, have no formal education, are in the poorer/middle/richer wealth quintile, and had no ANC visit and at home delivery (Model 4).

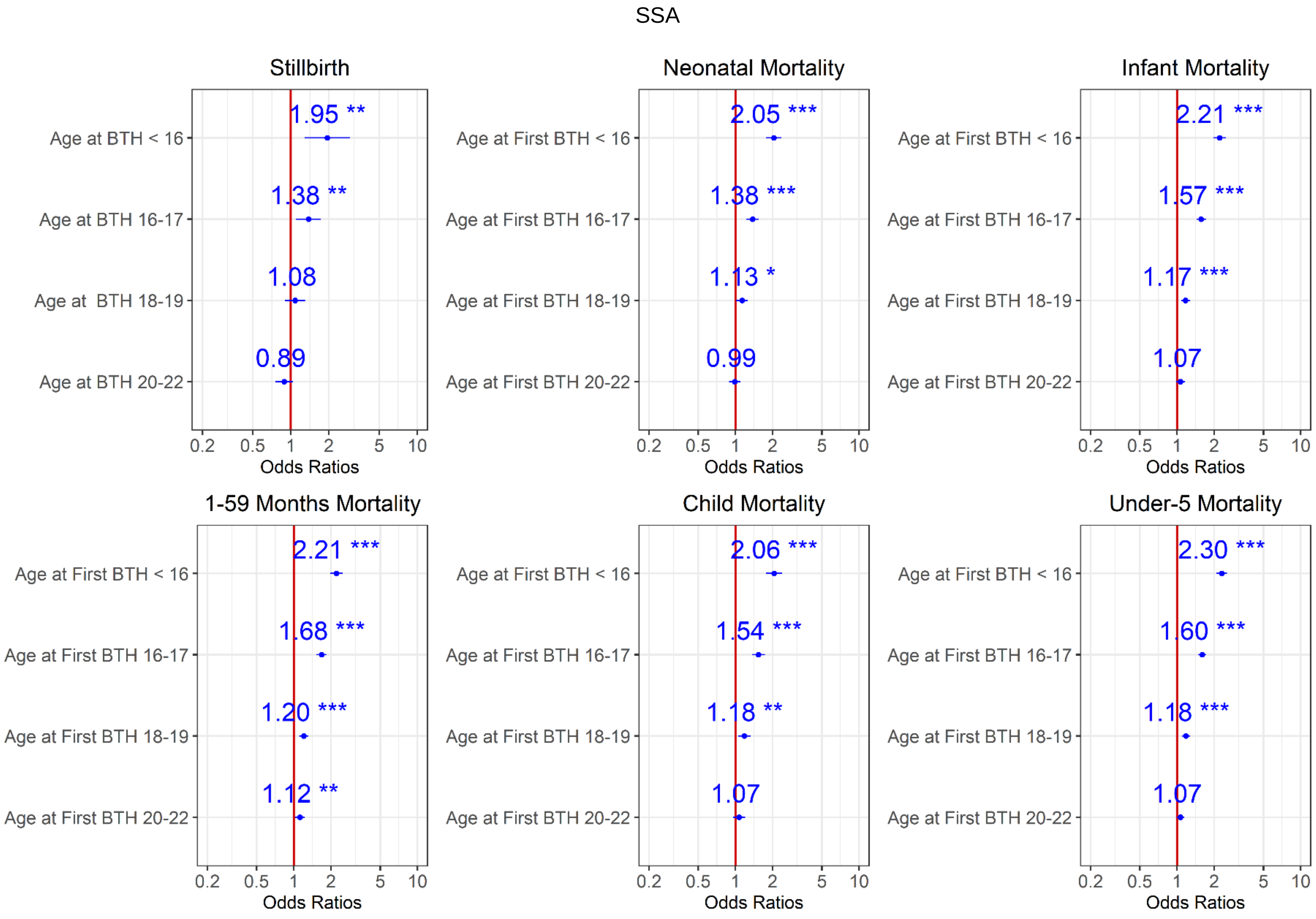
SSA
Stillbirth
Neonatal Mortality
Infant Mortality
1-59 Months Mortality
Child Mortality
Under-5 Mortality
Age at BTH < 16
Age at BTH 16-17
Age at  BTH 18-19
Age at BTH 20-22
Age at First BTH < 16
Age at First BTH 16-17
Age at First BTH 18-19
Age at First BTH 20-22
1.95 **
1.38 **
1.08
0.89
2.05 ***
1.38 ***
1.13 *
0.99
2.21 ***
1.57 ***
1.17 ***
1.07
2.21 ***
1.68 ***
1.20 ***
1.12 **
2.06 ***
1.54 ***
1.18 **
1.07
2.30 ***
1.60 ***
1.18 ***
1.07
0.2
0.5
1
2
5
10
Odds Ratios

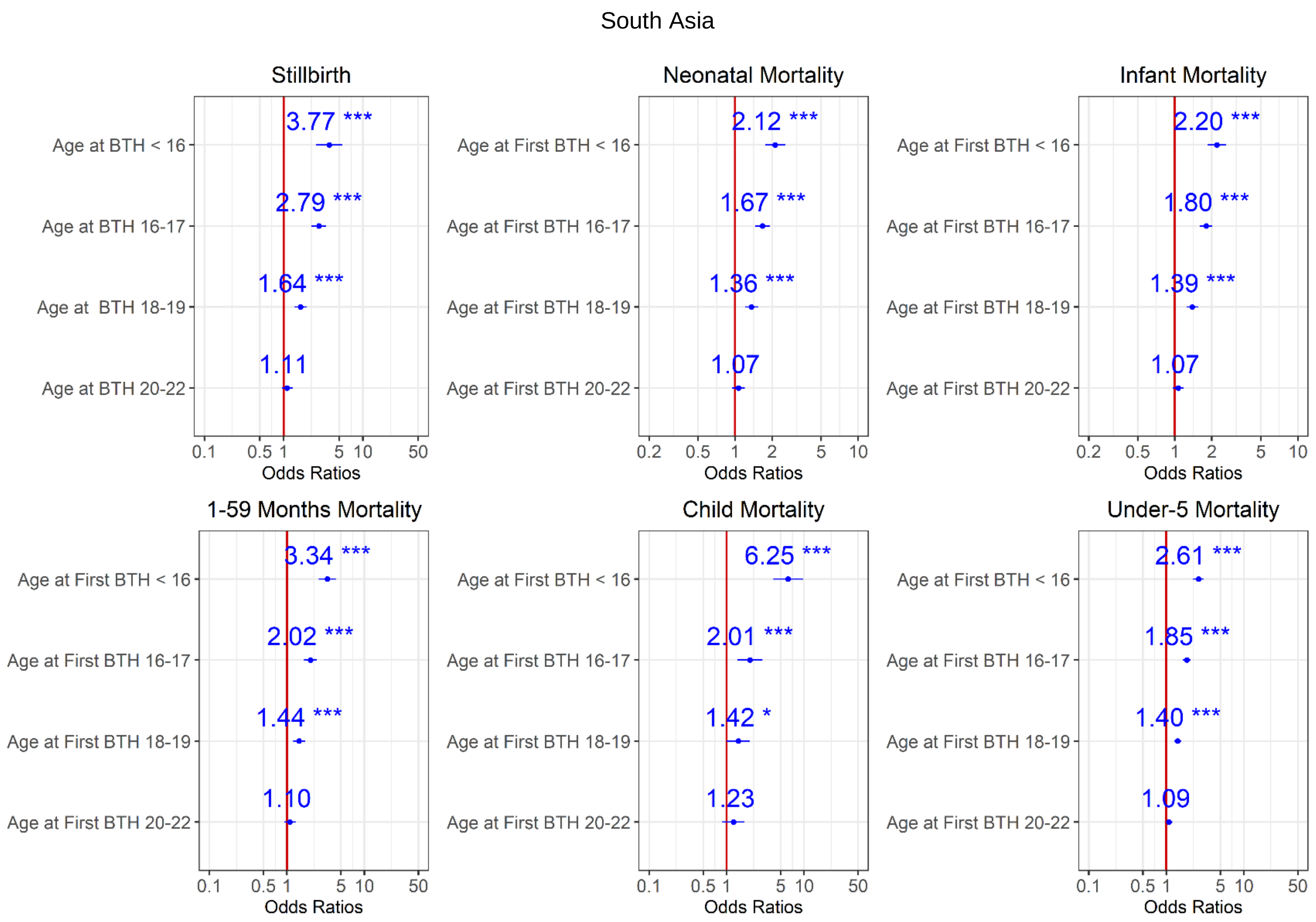


Figure 10. Ratios associated with neonate, infant, child, 1-59 months, under-5 years, and stillbirth in SSA and South Asia for the 2004-2008 survey period. Risk factors reducing the probability of death have odds ratios lower than 1 to the left of the vertical red line. Odds ratios (blue points), 95% confidence intervals (horizontal blue lines) are given. P-values are shown with the asterisk signs ('***' 0.001 '**' 0.01 '*' 0.05 '.' 0.1 ' ' 1). Reference group is mothers aged 23-25 years old (Model 0).

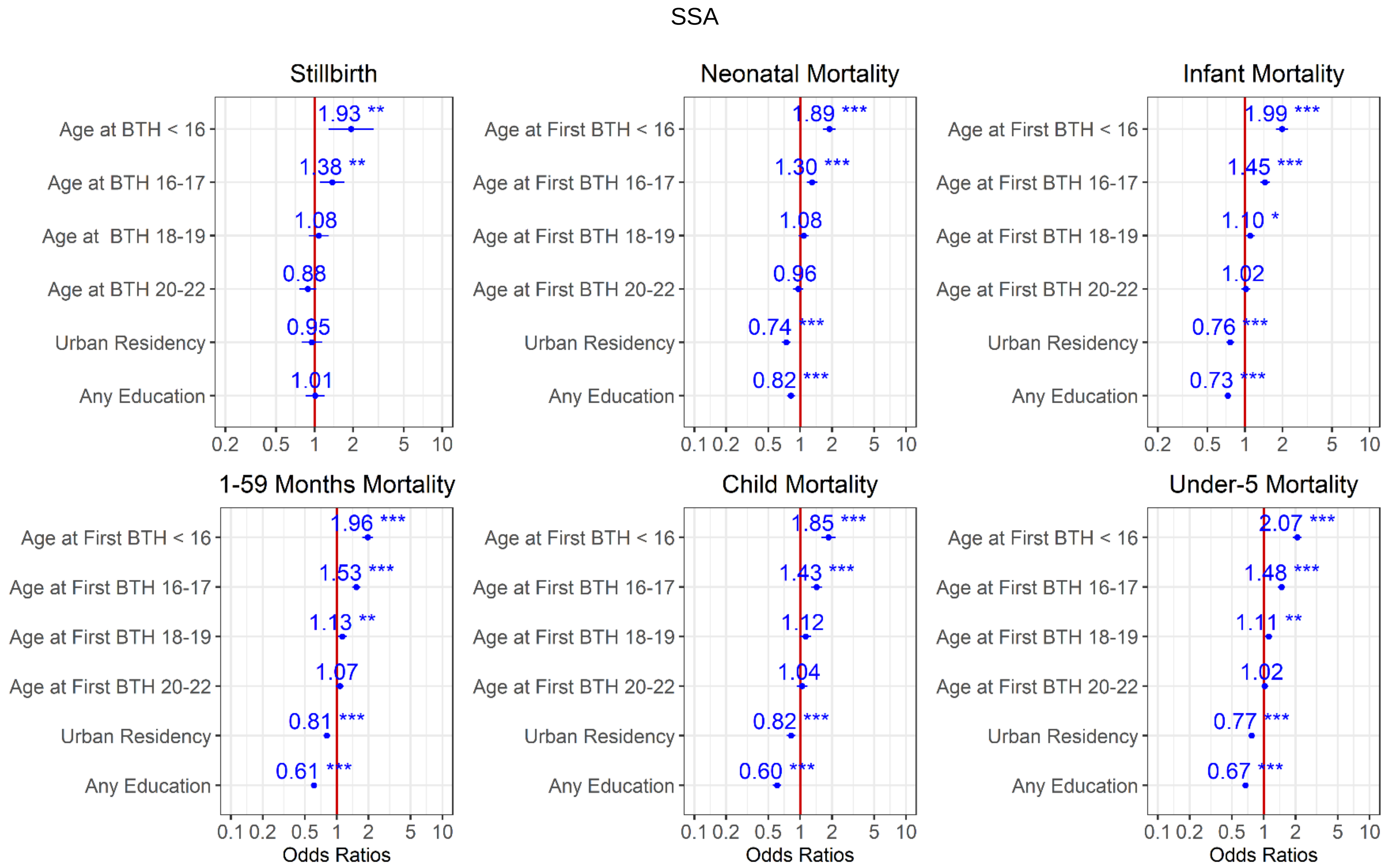
SSA
Stillbirth
Neonatal Mortality
Infant Mortality
1-59 Months Mortality
Child Mortality
Under-5 Mortality
Age at BTH < 16
Age at BTH 16-17
Age at BTH 18-19
Age at BTH 20-22
Age at First BTH < 16
Age at First BTH 16-17
Age at First BTH 18-19
Age at First BTH 20-22
Urban Residency
Any Education
1.93 **
1.38 **
1.08
0.88
0.95
1.01
1.89 ***
1.30 ***
1.08
0.96
0.74 ***
0.82 ***
1.99 ***
1.45 ***
1.10 *
1.02
0.76 ***
0.73 ***
1.96 ***
1.53 ***
1.13 **
1.07
0.81 ***
0.61 ***
1.85 ***
1.43 ***
1.12
1.04
0.82 ***
0.60 ***
2.07 ***
1.48 ***
1.11 **
1.02
0.77 ***
0.67 ***
0.1 0.2 0.5 1 2 5 10
Odds Ratios

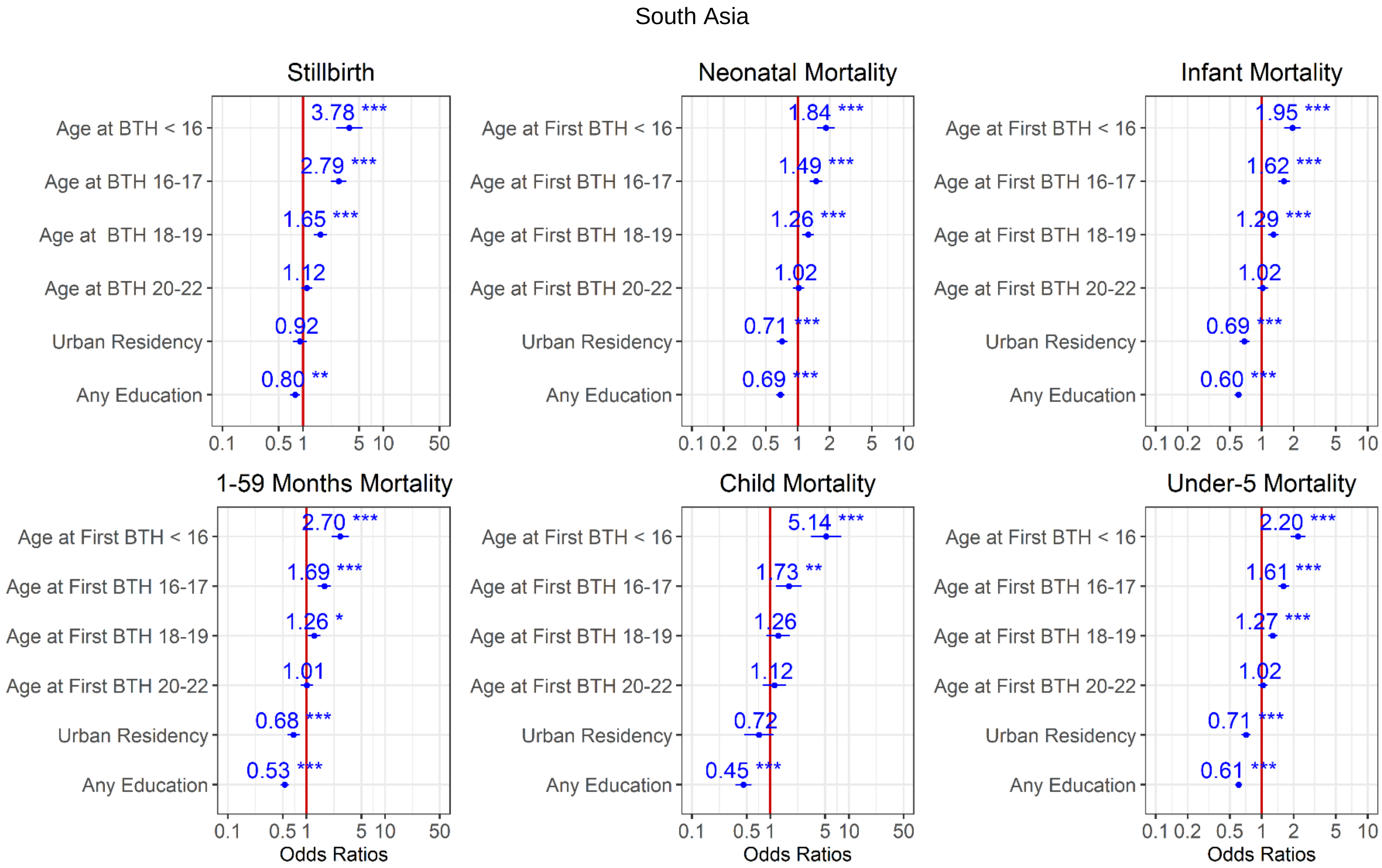


Figure 11. Ratios associated with neonate, infant, child, 1-59 months, under-5 years, and stillbirth in SSA and South Asia for the 2004-2008 survey period. Risk factors reducing the probability of death have odds ratios lower than 1 to the left of the vertical red line. Odds ratios (blue points), 95% confidence intervals (horizontal blue lines) are given. P-values are shown with the asterisk signs ('***' 0.001 '**' 0.01 '*' 0.05 '.' 0.1 ' ' 1). Reference group is mothers aged 23-25 years old who live in rural areas and have no formal education (Model 1).

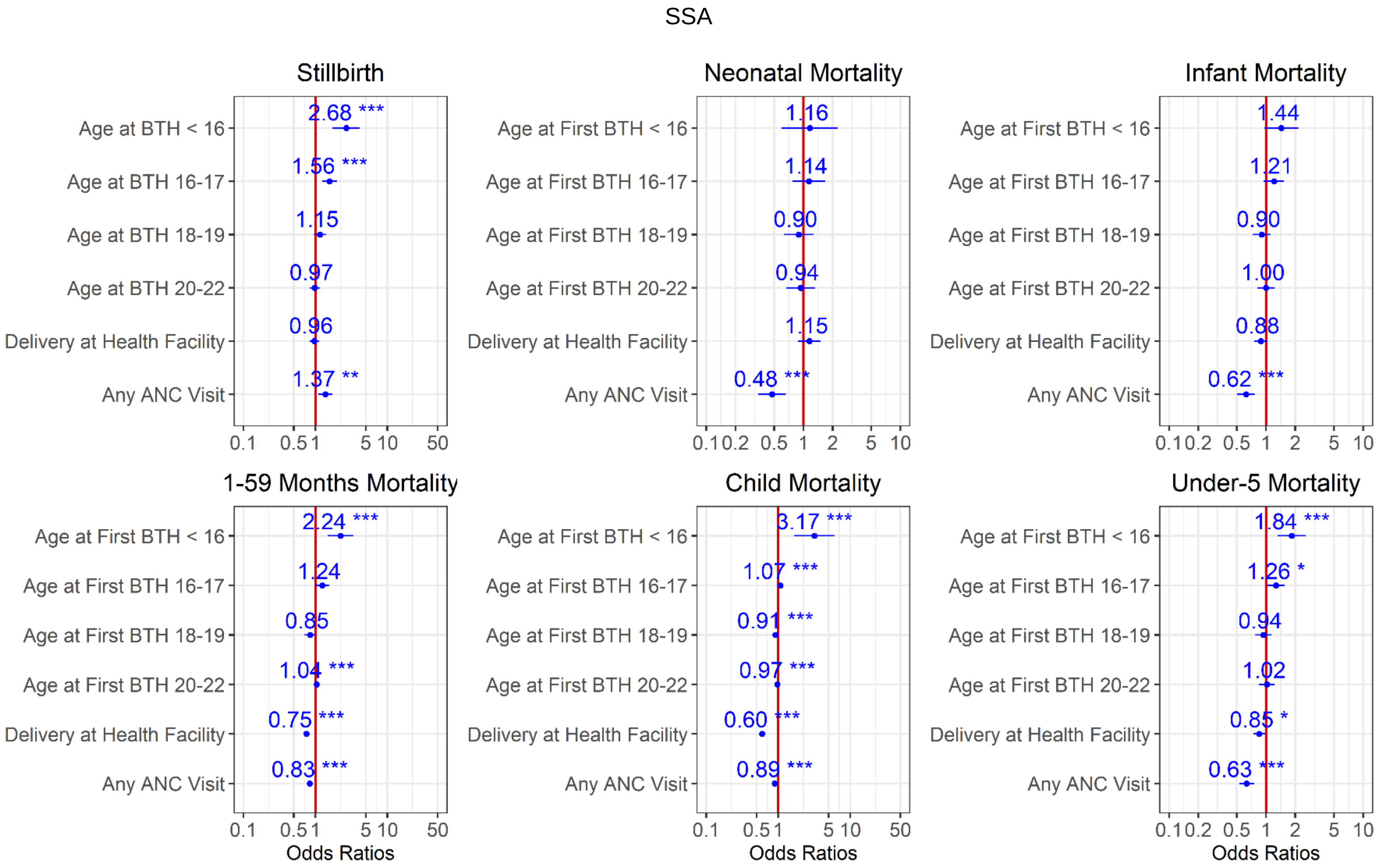
SSA
Stillbirth
Neonatal Mortality
Infant Mortality
1-59 Months Mortality
Child Mortality
Under-5 Mortality
Age at BTH < 16
Age at BTH 16-17
Age at BTH 18-19
Age at BTH 20-22
Age at First BTH < 16
Age at First BTH 16-17
Age at First BTH 18-19
Age at First BTH 20-22
Delivery at Health Facility
Any ANC Visit
2.68 ***
1.56 ***
1.15
0.97
0.96
1.37 **
1.16
1.14
0.90
0.94
1.15
0.48 ***
1.44
1.21
0.90
1.00
0.88
0.62 ***
2.24 ***
1.24
0.85
1.04 ***
0.75 ***
0.83 ***
3.17 ***
1.07 ***
0.91 ***
0.97 ***
0.60 ***
0.89 ***
1.84 ***
1.26 *
0.94
1.02
0.85 *
0.63 ***
Odds Ratios

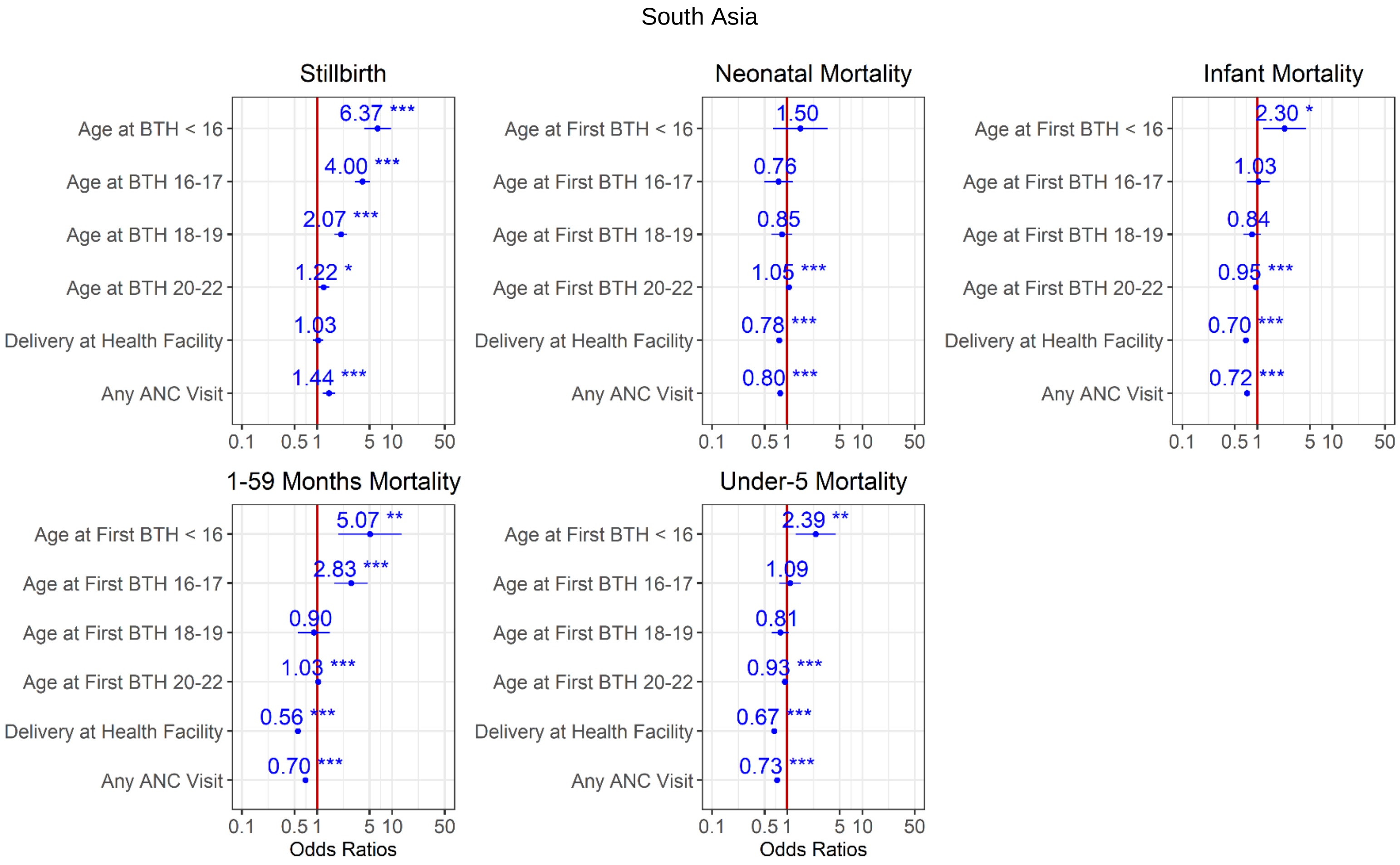


Figure 12. Ratios associated with neonate, infant, child, 1-59 months, under-5 years, and stillbirth in SSA and South Asia for the 2004-2008 survey period. Risk factors reducing the probability of death have odds ratios lower than 1 to the left of the vertical red line. Odds ratios (blue points), 95% confidence intervals (horizontal blue lines) are given. P-values are shown with the asterisk signs (‘***’ 0.001 ‘**’ 0.01 ‘*’ 0.05 ‘.’ 0.1 ‘ ’ 1). Reference group is mothers aged 23-25 years old who delivered at home and have no ANC visits (Model 2).

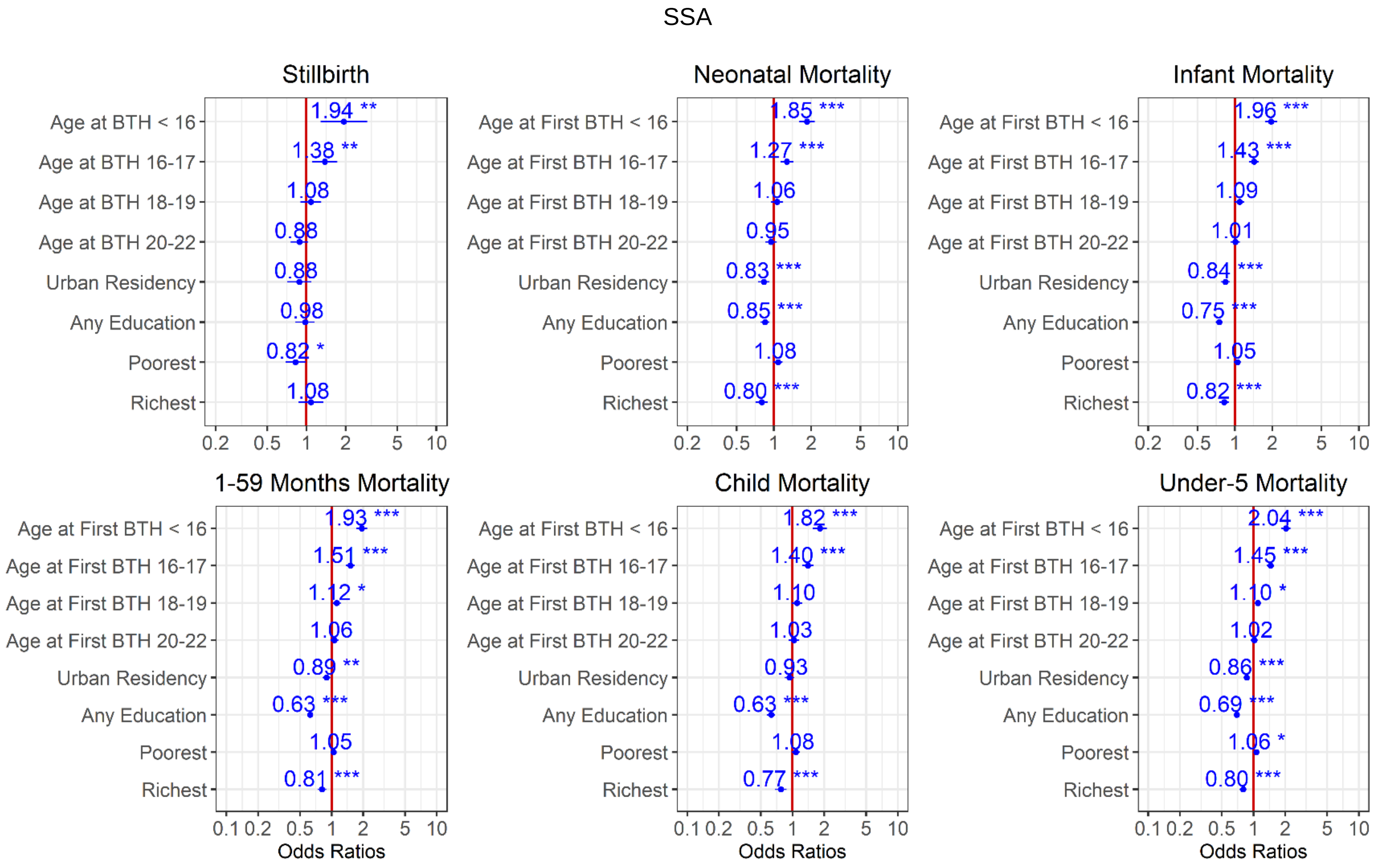
SSA
Stillbirth
Neonatal Mortality
Infant Mortality
1-59 Months Mortality
Child Mortality
Under-5 Mortality
Age at BTH < 16
Age at BTH 16-17
Age at BTH 18-19
Age at BTH 20-22
Age at First BTH < 16
Age at First BTH 16-17
Age at First BTH 18-19
Age at First BTH 20-22
Urban Residency
Any Education
Poorest
Richest
1.94 **
1.38 **
1.08
0.88
0.88
0.98
0.82 *
1.08
1.85 ***
1.27 ***
1.06
0.95
0.83 ***
0.85 ***
1.08
0.80 ***
1.96 ***
1.43 ***
1.09
1.01
0.84 ***
0.75 ***
1.05
0.82 ***
1.93 ***
1.51 ***
1.12 *
1.06
0.89 **
0.63 ***
1.05
0.81 ***
1.82 ***
1.40 ***
1.10
1.03
0.93
0.63 ***
1.08
0.77 ***
2.04 ***
1.45 ***
1.10 *
1.02
0.86 ***
0.69 ***
1.06 *
0.80 ***
0.1 0.2 0.5 1 2 5 10
Odds Ratios

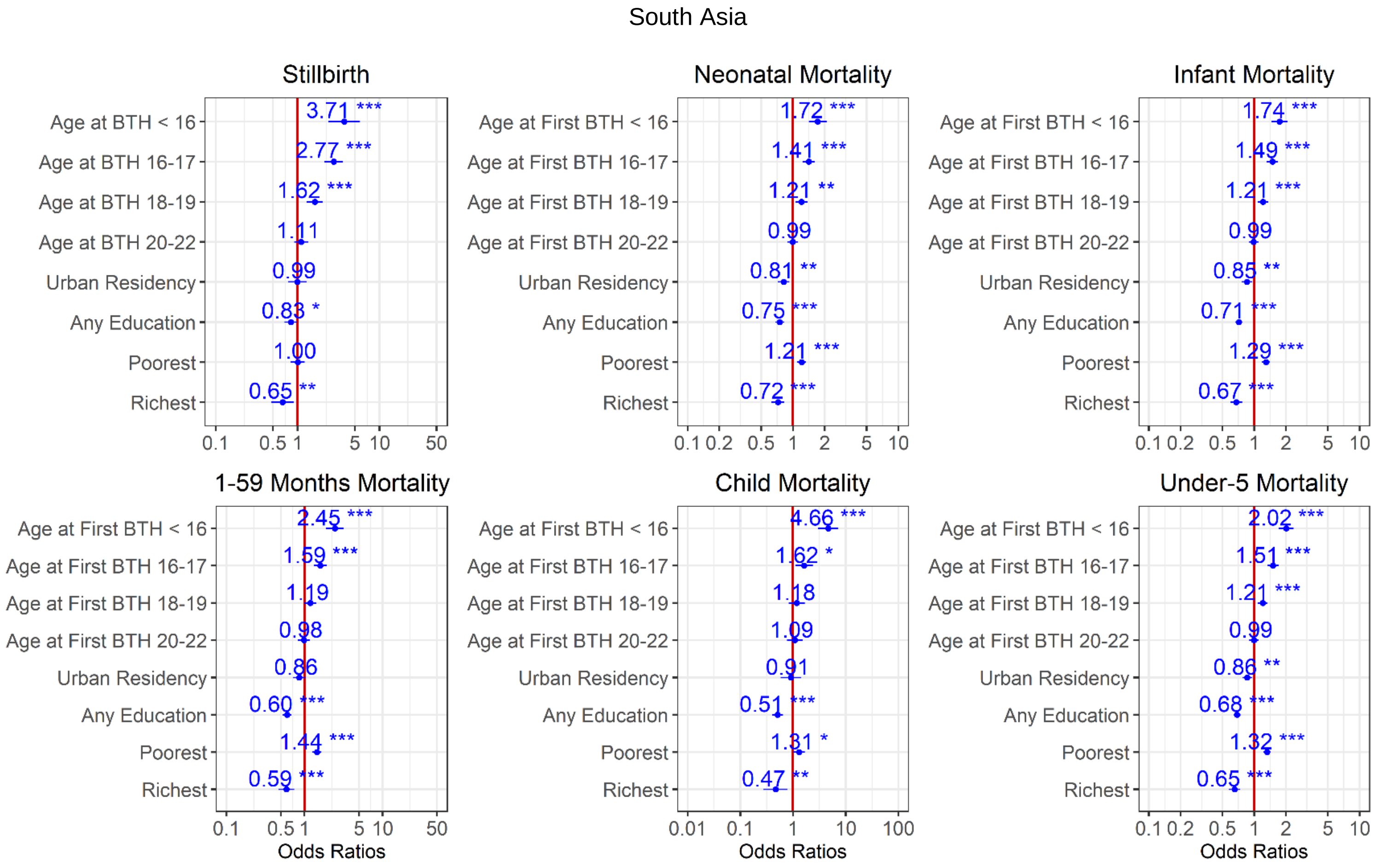


Figure 13. Ratios associated with neonate, infant, child, 1-59 months, under-5 years, and stillbirth in SSA and South Asia for the 2004-2008 survey period. Risk factors reducing the probability of death have odds ratios lower than 1 to the left of the vertical red line. Odds ratios (blue points), 95% confidence intervals (horizontal blue lines) are given. P-values are shown with the asterisk signs ('***' 0.001 '**' 0.01 '*' 0.05 '.' 0.1 ' ' 1). Reference group is mothers aged 23-25 years old who live in rural areas, have no formal education, and are in the poorer/middle/richer wealth quintile, (Model 3).

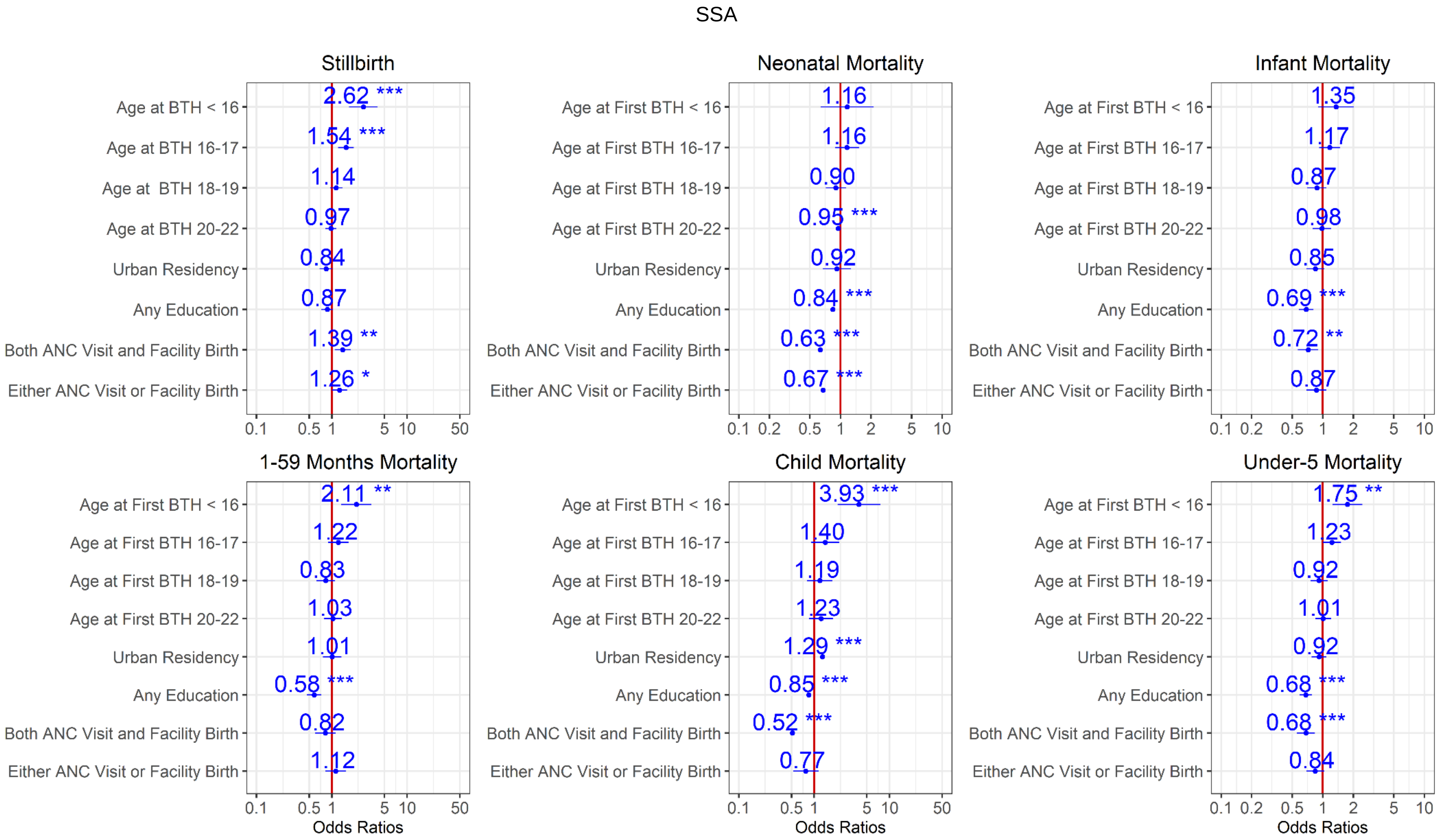
SSA
Stillbirth
Age at BTH < 16
Age at BTH 16-17
Age at BTH 18-19
Age at BTH 20-22
Urban Residency
Any Education
Both ANC Visit and Facility Birth
Either ANC Visit or Facility Birth
2.62 ***
1.54 ***
1.14
0.97
0.84
0.87
1.39 **
1.26 *
Neonatal Mortality
Age at First BTH < 16
Age at First BTH 16-17
Age at First BTH 18-19
Age at First BTH 20-22
1.16
1.16
0.90
0.95 ***
0.92
0.84 ***
0.63 ***
0.67 ***
Infant Mortality
1.35
1.17
0.87
0.98
0.85
0.69 ***
0.72 **
0.87
1-59 Months Mortality
2.11 **
1.22
0.83
1.03
1.01
0.58 ***
0.82
1.12
Child Mortality
3.93 ***
1.40
1.19
1.23
1.29 ***
0.85 ***
0.52 ***
0.77
Under-5 Mortality
1.75 **
1.23
0.92
1.01
0.92
0.68 ***
0.68 ***
0.84
0.1 0.5 1 5 10 50
0.1 0.2 0.5 1 2 5 10
Odds Ratios

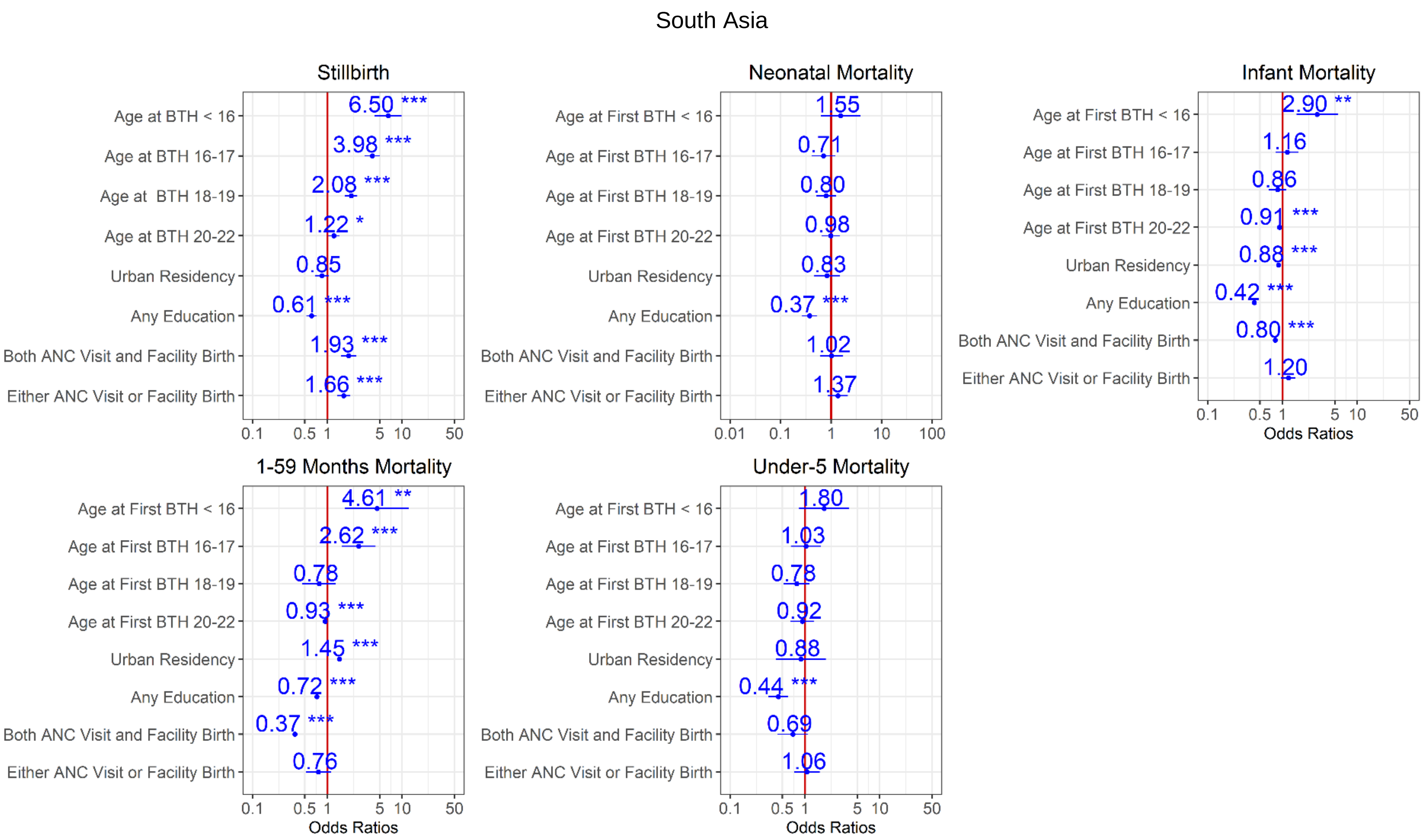


Figure 14. Ratios associated with neonate, infant, child, 1-59 months, under-5 years, and stillbirth in SSA and South Asia for the 2004-2008 survey period. Risk factors reducing the probability of death have odds ratios lower than 1 to the left of the vertical red line. Odds ratios (blue points), 95% confidence intervals (horizontal blue lines) are given. P-values are shown with the asterisk signs (‘***’ 0.001 ‘**’ 0.01 ‘*’ 0.05 ‘.’ 0.1 ‘ ’ 1). Reference group is mothers aged 23-25 years old who live in rural areas, have no formal education, are in the poorer/middle/richer wealth quintile, and had no ANC visit and at home delivery (Model 4).

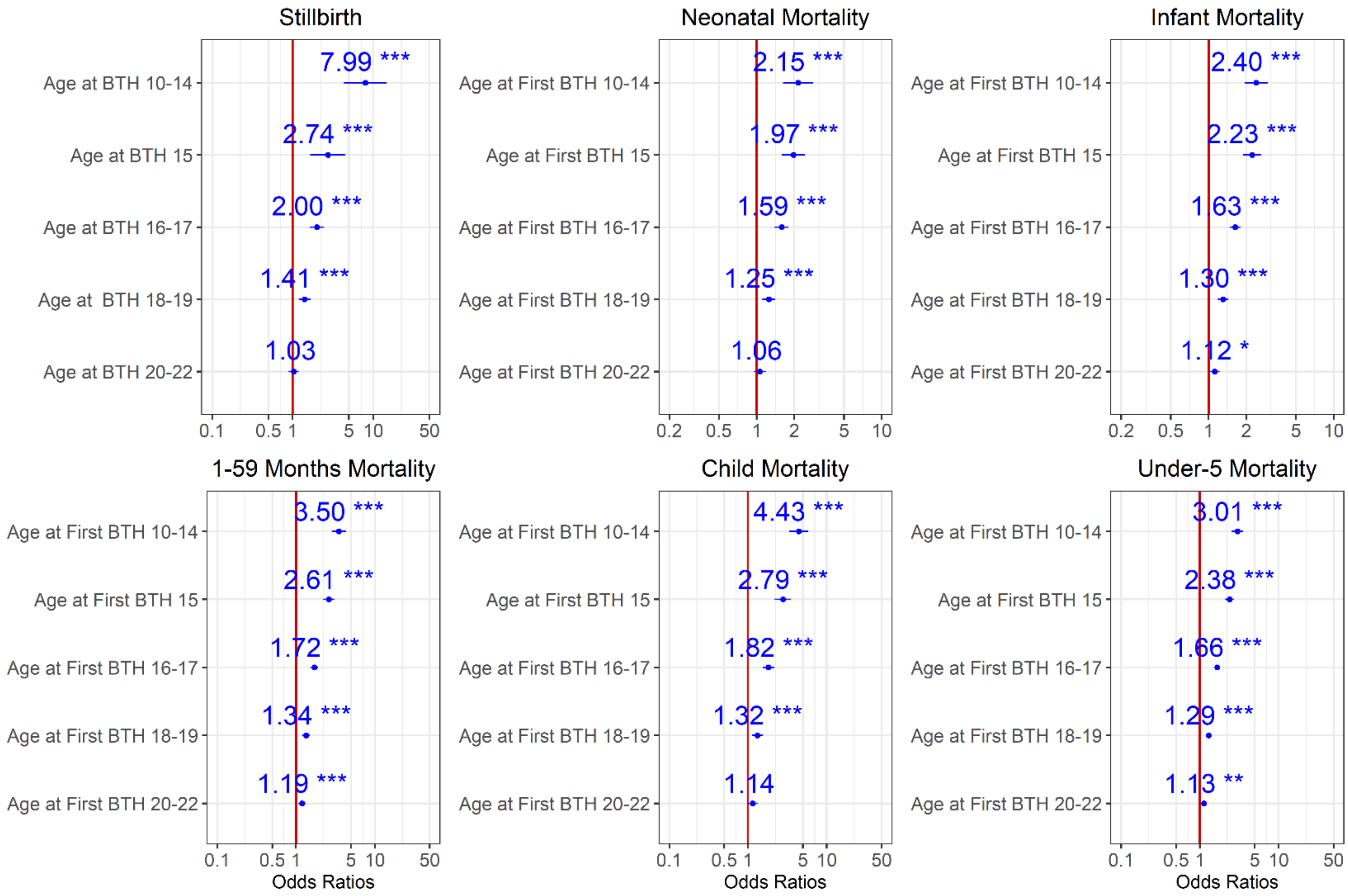


Figure 15. Ratios associated with neonate, infant, child, 1-59 months, under-5 years, and stillbirth in SSA for the 2014-2018 survey period. Risk factors reducing the probability of death have odds ratios lower than 1 to the left of the vertical red line. Odds ratios (blue points), 95% confidence intervals (horizontal blue lines) are given. P-values are shown with asterisk signs (‘***’ 0.001 ‘**’ 0.01 ‘*’ 0.05 ‘.’ 0.1 ‘ ’ 1). Reference group is mothers aged 23-25 years old.

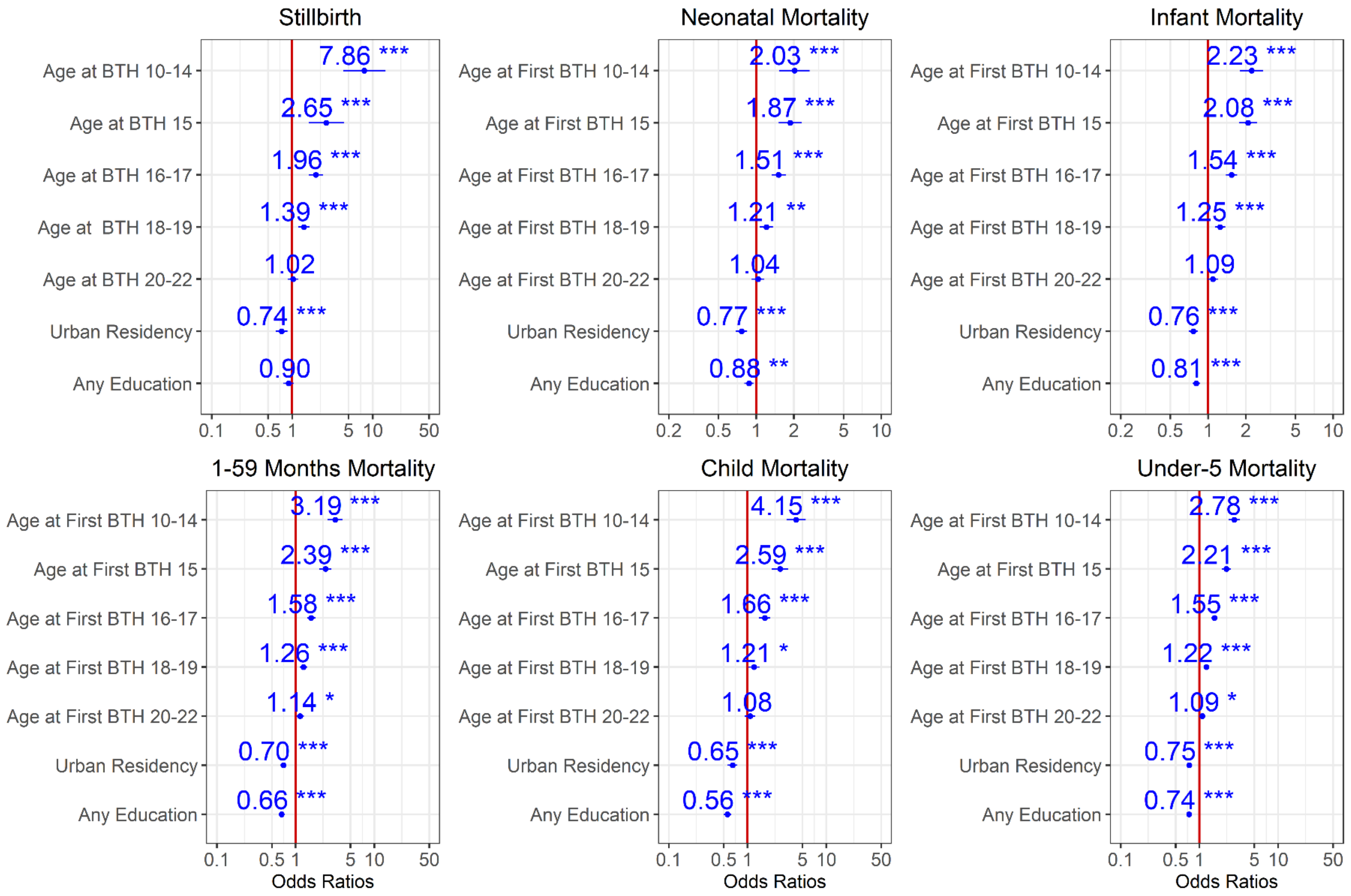


Figure 16. Ratios associated with neonate, infant, child, 1-59 months, under-5 years, and stillbirth in SSA for the 2014-2018 survey period. Risk factors reducing the probability of death have odds ratios lower than 1 to the left of the vertical red line. Odds ratios (blue points), 95% confidence intervals (horizontal blue lines) are given. P-values are shown with asterisk signs (‘***’ 0.001 ‘**’ 0.01 ‘*’ 0.05 ‘.’ 0.1 ‘ ’ 1). Reference group is mothers aged 23-25 years old who live in rural areas, and have no formal education.
